# Mucosal IgA to pre-fusion F protein predicts protection from RSV infection in a high burden setting

**DOI:** 10.64898/2026.03.16.26348479

**Authors:** David Hodgson, Sheikh Jarju, Laura Gatcombe, Tom Coleman, Giulia Dowgier, Rhys D Wenlock, Benjamin B Lindsey, Madikoi Danso, Natalie Barratt, Marie Gomes, Irina Grouneva, Ya Jankey Jagne, Beate Kampmann, Mary Y. Wu, Ashley Otter, Stefan Flasche, Adam Kucharski, Thushan I. de Silva

## Abstract

Respiratory syncytial virus (RSV) causes over 100,000 annual deaths, predominantly in low-and middle-income countries, yet immunological correlates of protection remain poorly defined in high-burden settings. We conducted an intensively sampled prospective household cohort study in The Gambia, following 342 participants over 12 months with weekly PCR testing and longitudinal serum and nasal sampling. Using a Bayesian hierarchical framework jointly modelling antibody kinetics, household transmission dynamics, and imperfect PCR ascertainment, we characterised humoral immunity to RSV-A antigens across systemic and mucosal compartments. Mucosal IgA to the pre-fusion F (pre-F) protein was the strongest predictor of protection against infection (AUC = 0.72), outperforming serum anti-Pre-F IgG (AUC = 0.66). Serum and mucosal responses were largely compartment-independent (r = 0.02–0.28), with a combined serum and mucosal anti-Pre-F model outperforming either alone, and both biomarkers correlating with RSV-A neutralising titres. These findings have direct implications for designing RSV infection preventing vaccine strategies.

## INTRODUCTION

Respiratory syncytial virus (RSV) causes approximately 33 million acute lower respiratory tract infection episodes annually in children under five years, resulting in over 100,000 deaths, with more than 97% occurring in low- and middle-income countries (LMICs).^1^ Infants under six months bear a disproportionate burden, accounting for one in five RSV episodes but nearly half of all RSV-attributable deaths.^1^ In fact, within community settings in LMICs, RSV is responsible for 5% of all infants deaths.^2^ The landscape of RSV prevention has recently changed with the regulatory approval of nirsevimab, a long-acting monoclonal antibody for infant immunisation, Abrysvo, a maternal RSV vaccine, and Arexy, a vaccine for adults aged 60 years and older.^3–6^ Early effectiveness data demonstrate that nirsevimab provides 90% protection against RSV-associated hospitalisation in infants,^7^ while maternal vaccination reduces RSV hospitalisation risk for infants in the months following birth by up to 72%.^8^ In older adults, RSV vaccines show 75% effectiveness against severe lower respiratory tract disease.^9,10^

Despite these advances, fundamental questions about RSV immunity remain unanswered. The mechanisms underlying incomplete protection following infection, permitting repeated RSV infections throughout life, are poorly understood.^11^ A critical unresolved question is whether and how asymptomatic RSV infections, which likely constitute most exposures, boost and maintain protective immunity in healthy individuals. Key knowledge gaps also persist regarding the kinetics and durability of antibody responses to different RSV antigens, the relative contributions of serum versus mucosal immunity in conferring protection, and age-dependent differences in immune responses, which may inform optimal vaccination strategies.^11^ Moreover, the majority of existing immunological data derive from high-income settings, despite LMIC bearing over 95% of the RSV disease burden.^12^ This geographical imbalance causes issues because populations in high-transmission resource-limited settings experience high levels of RSV exposure throughout life, potentially leading to distinct patterns of immune boosting, antibody persistence and protective immunity compared to low-transmission settings.^13–15^ Understanding immune dynamics in high-burden contexts is therefore important for understanding how repeated natural exposures shape population-level immunity, and for developing vaccination strategies designed for the populations at greatest risk.

Establishing immunological correlates of protection can accelerate vaccine development, help optimise immunisation schedules, and evaluate vaccine effectiveness in diverse populations. Recent successes using maternal RSV vaccines and passive infusion of long-acting monoclonal antibodies have demonstrated the clinical efficacy of serum antibody-based interventions targeting the pre-F protein. These studies confirm that serum IgG responses can provide substantial protection against medically attended and hospitalised severe RSV outcomes. However, the immunological determinants of protection against infection and subclinical disease, which drive community transmission and may be relevant for paediatric vaccine development, remain poorly defined. Recent evidence from studies in infected children and community-dwelling in older adults suggests that the mucosal immune response may play a distinct and potentially superior role in preventing RSV infection compared to serum antibodies.^16,17^ Furthermore, the relationship between antibody levels and protection may vary across age groups, RSV subtypes, and antigenic targets, necessitating comprehensive longitudinal studies that capture the full spectrum of humoral responses across systemic and mucosal compartments and linking these to asymptomatic and subclinical disease outcomes.^18^

Here, we present a comprehensive analysis of RSV antibody kinetics and correlates of protection from a prospective household cohort study in The Gambia, where respiratory virus surveillance data and SARS-CoV-2 immune kinetics have been described extensively in prior studies.^19–22^ Using Bayesian hierarchical modelling, we jointly estimate the kinetics of antibody responses following infection, quantify age-dependent patterns in antibody boosting and maintenance, and identify immunological correlates of protection, linking pre-exposure antibody levels to infection risk while accounting for household transmission dynamics. This study addresses critical knowledge gaps by: (1) providing detailed immunological data from a high-transmission West African setting, (2) characterising how asymptomatic and mild RSV infections boost and maintain immunity across the life course; (3) simultaneously measuring systemic and mucosal antibody response within individuals and (4) identifying which immunological biomarkers best predict protection from infection across different age groups.

## RESULTS

### DESCRIPTIVE ANALYSIS OF RSV PCR-CONFIRMED AND SERO-INFERRED CASES

We followed 342 participants across 52 households over 12 months from a peri-urban region in The Gambia with weekly respiratory PCR surveillance to capture both symptomatic and asymptomatic RSV infections. Serum IgG and mucosal IgA responses to multiple RSV antigens (pre-fusion (PreF), post-fusion (PostF), G protein, and nucleoprotein (NP) for RSV-A and RSV-B) were measured at enrollment, 6 months and 12 months using a multiplex binding antibody immunoassay. We documented 81 PCR-confirmed infections in 76 individuals across 40 households, with three individuals having a second infection, and one individual with a second and third infection. All first RSV infections occurred between enrollment and the 6-month bleeds. Most infections were RSV-A serotype, with 3/81 of the PCR-positive cases having an RSV-B infection. Of these PCR-confirmed infections, only 17.3% (14/81) were associated with symptoms during the week before or after PCR positivity (**Figure 1A-B**). To account for imperfect PCR detection, we assumed that PCR captured 80% of true infections and used consistent boosting across multiple antibody biomarkers at the individual level as evidence of infection in the remaining undetected cases (**Supplementary Methods 3.1**). This approach identified 18 additional infections in the PCR-negative individuals, yielding a total attack rate of 27% (94/342) over the 12-month period. Attack rates varied substantially by age groups, with children under 5 years having the highest attack rate at 53%, decreasing with age to 11% in adults aged ≥46 years (**Supplementary Figure 1**).

**Figure 1.**
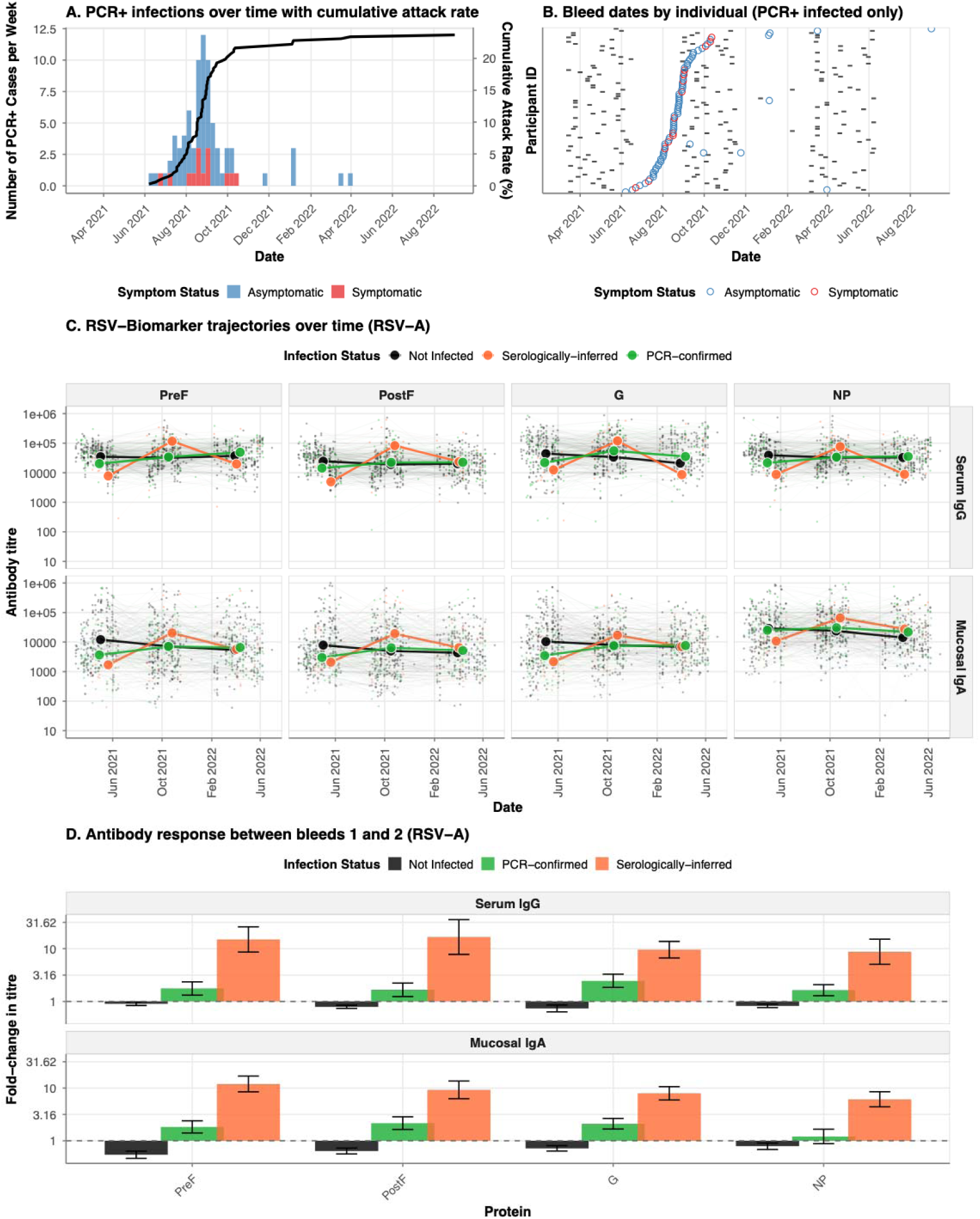
Epidemiology of RSV infections and antibody kinetics during the 2022-2023 epidemic season. **A.** Weekly incidence of PCR-confirmed RSV infections (stacked histogram) and cumulative attack rate (black line) among 342 household cohort participants from October 2022 to May 2023. Bars are colored by symptom status at detection: symptomatic (red, detected at unscheduled visits) or asymptomatic (blue, detected at scheduled home or clinic visits). B. Timeline of serological sampling for PCR-confirmed infected individuals. Each horizontal line represents one participant (ordered by date of PCR-confirmed infection), with grey tiles indicating bleed dates and colored circles marking the timing of PCR-confirmed infection (colored by symptom status as in panel a). C. Longitudinal trajectories of serum IgG (top row) and mucosal IgA (bottom row) antibody responses against four RSV-A proteins (PreF, PostF, G, NP) over time. Individual participant trajectories are shown as thin lines with low opacity, colored by infection status: not infected (black), serologically-defined infections (orange), and PCR-confirmed infections (green). D. Mean fold-change in antibody titres between the first bleed (pre-epidemic baseline) and second bleed (post-epidemic) for serum IgG and mucosal IgA responses to RSV-A proteins. Bars represent mean fold-change (log10 scale) stratified by infection status, with error bars indicating standard error.

### DESCRIPTIVE ANALYSIS OF IMMUNOLOGICAL BIOMARKERS AT MEASURED TIMEPOINTS

RSV biomarker trajectories were tracked across the 342 participants over the study for sixteen different biomarkers. Individuals with PCR-confirmed infections exhibited mean-fold increases of 1.6–2.5 in serum IgG and 1.2–2.1 in mucosal IgA across the four RSV-A viral proteins (**Figures 1C–D)** between the enrollment and 6-month visit. PCR-negative but serologically-defined infected individuals also showed substantial antibody boosting across the same biomarkers, with mean-fold increases of 8.7-16.7 in serum IgG and 6.1-12.0 in mucosal IgA between the same two visits. In contrast, uninfected participants showed decay during the same period, with mean serum IgG fold-changes of 0.7–0.9 and mucosal IgA fold-changes of 0.5–0.8. The responses to RSV-B viral proteins following RSV-A infections were similar, demonstrating the significant cross-reactivity that exists in binding antibody titres (**Supplementary Figure 2)**.

### RECONSTRUCTING ANTIBODY KINETICS FOLLOWING RSV INFECTION

We characterised the magnitude and durability of antibody responses following RSV-A infection (including both PCR-positive and serologically defined cases) by analysing fitted serum IgG and mucosal IgA kinetic curves against the four viral proteins PreF, PostF, G, and NP (**Figure 2A**). From these fitted kinetic curves, we found that RSV infection induced sustained antibody boosting across all viral proteins, with mean peak fold-rises (i.e. relative change between baseline and peak of antibody kinetic curves) of 3.0–8.0-fold above baseline titres for serum IgG and 2.7–6.3-fold for mucosal IgA. Mucosal IgA responses to surface glycoproteins PreF and PostF exhibited stronger boosting (6.3- and 4.7-fold rise) than serum IgG to the same proteins (3.9- and 3.5-fold rise). Mucosal IgA to PreF and PostF also showed more sustained persistence (220 and 235 days above 2-fold of baseline level) than serum IgG to the same proteins (175 and 132 days). In contrast, serum IgG to G protein showed exceptional boosting (8.0-fold rise) compared to mucosal IgA (5.55-fold rise), but faster waning kinetics (177 days above 2-fold of baseline) than anti-G mucosal IgA (235 days above 2-fold of baseline; **Figure 2B**). Consistent with internal protein localisation, antibodies to NP showed modest responses in both compartments (3.0-fold serum IgG, 2.7-fold mucosal IgA) with rapid waning (109 and 85 days above 2-fold baseline). Similar patterns were observed for RSV-B, with surface proteins generally showing better boosting and longer persistence, and mucosal IgA responses proving more durable than serum IgG (**Supplementary Figure 3**).

**Figure 2.**
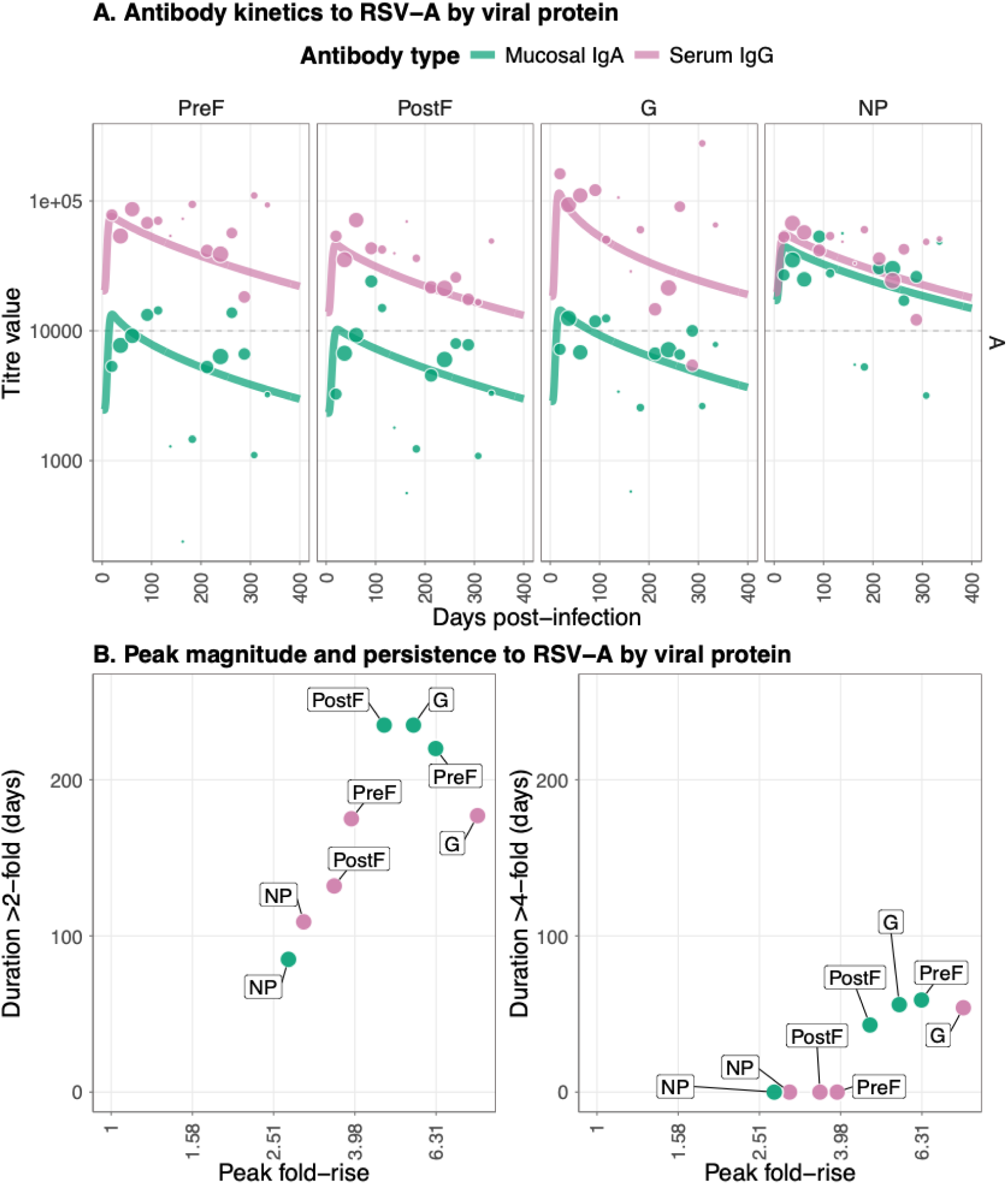
Comparison of single and dual biomarker models for predicting protection against RSV-A infection. A. Post-infection longitudinal antibody titers for four viral RSV-A proteins (PreF, PostF, G, and NP) across different antibody types; serum IgG and mucosal IgA. Lines show the median posterior predictive fit from the fitted Bayesian model, and the points show the observational titre data, with the size correlating with the sample size for that bin. B Peak antibody (x axis) and persistence measured as duration above a 2-fold (left panel) and 4-fold titre rise (right panel) in days. Data points show median posterior values for measurements of four viral RSV-A proteins (PreF, PostF, G, and NP) across different antibody types: serum IgG and mucosal IgA.

### IDENTIFICATION AND VALIDATION OF CORRELATES OF PROTECTION AGAINST RSV INFECTION

To identify immune biomarkers predictive of protection against RSV infection, we evaluated the predictive performance of IgG and IgA to RSV viral proteins after fitting logistic models (**Supplementary Figure 4**). Among single biomarker models, mucosal IgA to RSV-A PreF was the best-performing predictor of protection against infection (**Figure 3A**), achieving the highest out-of-sample predictive accuracy (LOO-EPLD) and discrimination; AUC = 0.72 (95% CrI 0.66–0.78). Notably, this was a better predictor than the target of current preventive strategies, serum IgG to RSV PreF (AUC = 0.66, 95% Crl 0.59 – 0.73). To validate the relevance of PreF as an antigenic target, we measured live virus neutralisation in a subset of 97 samples and calculated Pearson correlation coefficients with antibody levels to different RSV proteins. This analysis confirmed that PreF antibodies demonstrated the strongest correlation with neutralising activity in both compartments (serum IgG and mucosal IgA), though serum IgG exhibited stronger correlations than mucosal IgA (Pearson’s R 0.743 vs. 0.428). (**Figure 3B** and **Supplementary Figure 5**).

**Figure 3.**
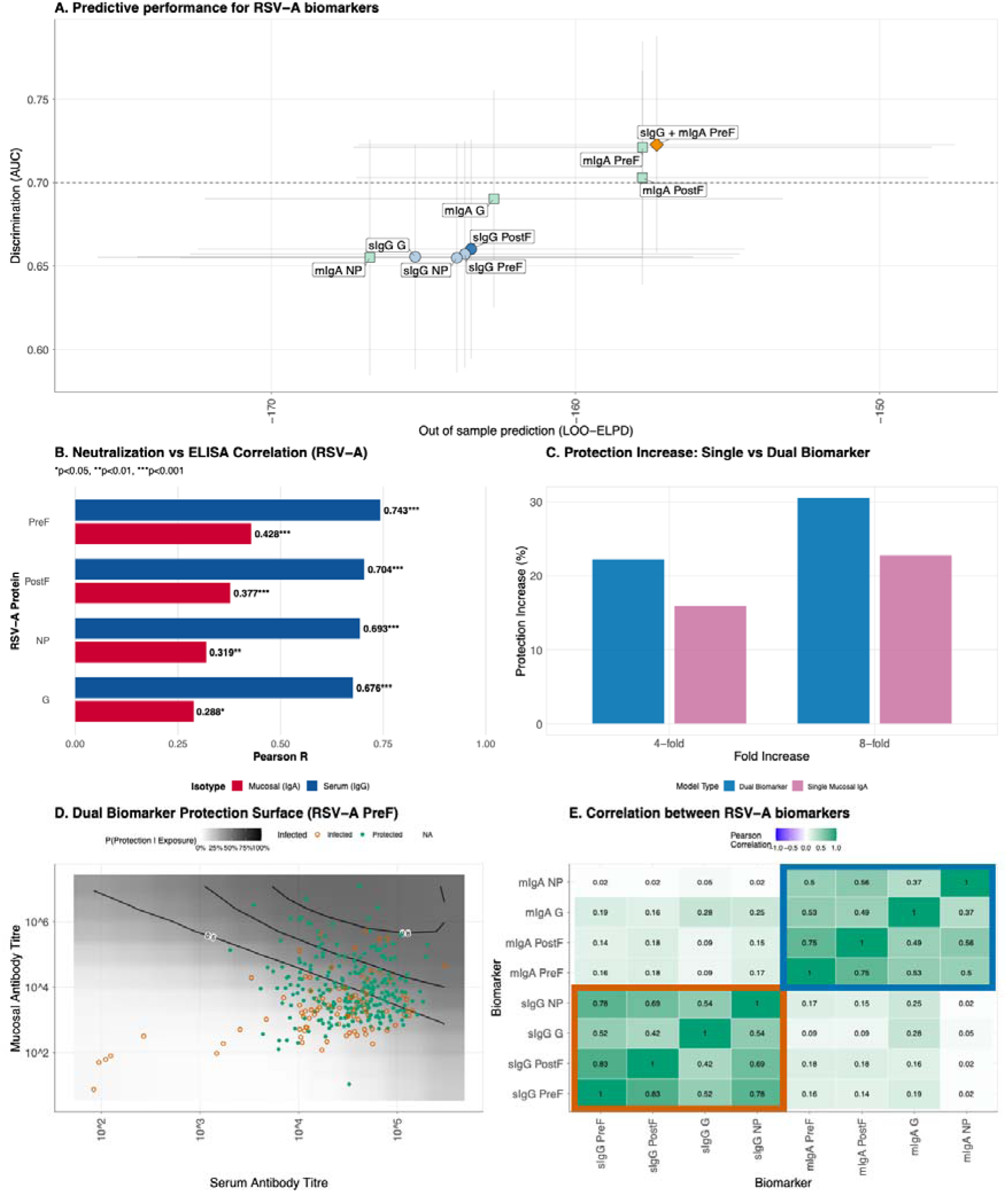
Comparison of single and dual biomarker models for predicting protection against RSV-A infection. A. Model performance comparison across single biomarker models and the dual biomarker model, defined by out-of-sample predictive accuracy (LOO-ELPD, x-axis) and discrimination ability (area under the ROC curve, AUC, y-axis). Circles indicate serum IgG models, squares represent mucosal IgA models, and the triangle denotes the dual biomarker model combining serum IgG and mucosal IgA to RSV-A PreF. The best-performing model within each biomarker class is highlighted with darker shading. Error bars show the standard error of LOO-ELPD (horizontal) and 95% confidence intervals for AUC (vertical). The dashed horizontal line indicates an AUC of 0.7. B. Summary of Pearson correlation coefficients for RSV-A protein antigens, comparing serum IgG (blue) and mucosal IgA (red) binding titres with neutralisation IC50 values. C. Counterfactual analysis comparing the increase in protection from boosting pre-infection antibody titres by 4-fold or 8-fold using either a single mucosal IgA biomarker model (purple) or a dual biomarker model combining mucosal IgA and serum IgG (blue). D. Two-dimensional protection surface for the dual biomarker model using serum IgG and mucosal IgA antibodies to RSV-A PreF. The continuous surface represents the predicted probability of protection given exposure as a function of both serum (x-axis) and mucosal (y-axis) antibody titres. Black contour lines delineate regions of constant protection probability at 50%, 70%, 80%, and 90%. Individual data points overlay the surface, with filled circles representing participants who were protected despite exposure and open circles representing those who became infected. Antibody titres are displayed on a log10 scale. E. Pearson correlation coefficients between RSV-A variant biomarkers at the time of infection. Color intensity represents correlation strength, ranging from blue (weak correlation) through white (no correlation) to green (strong positive correlation).

**Figure 4.**
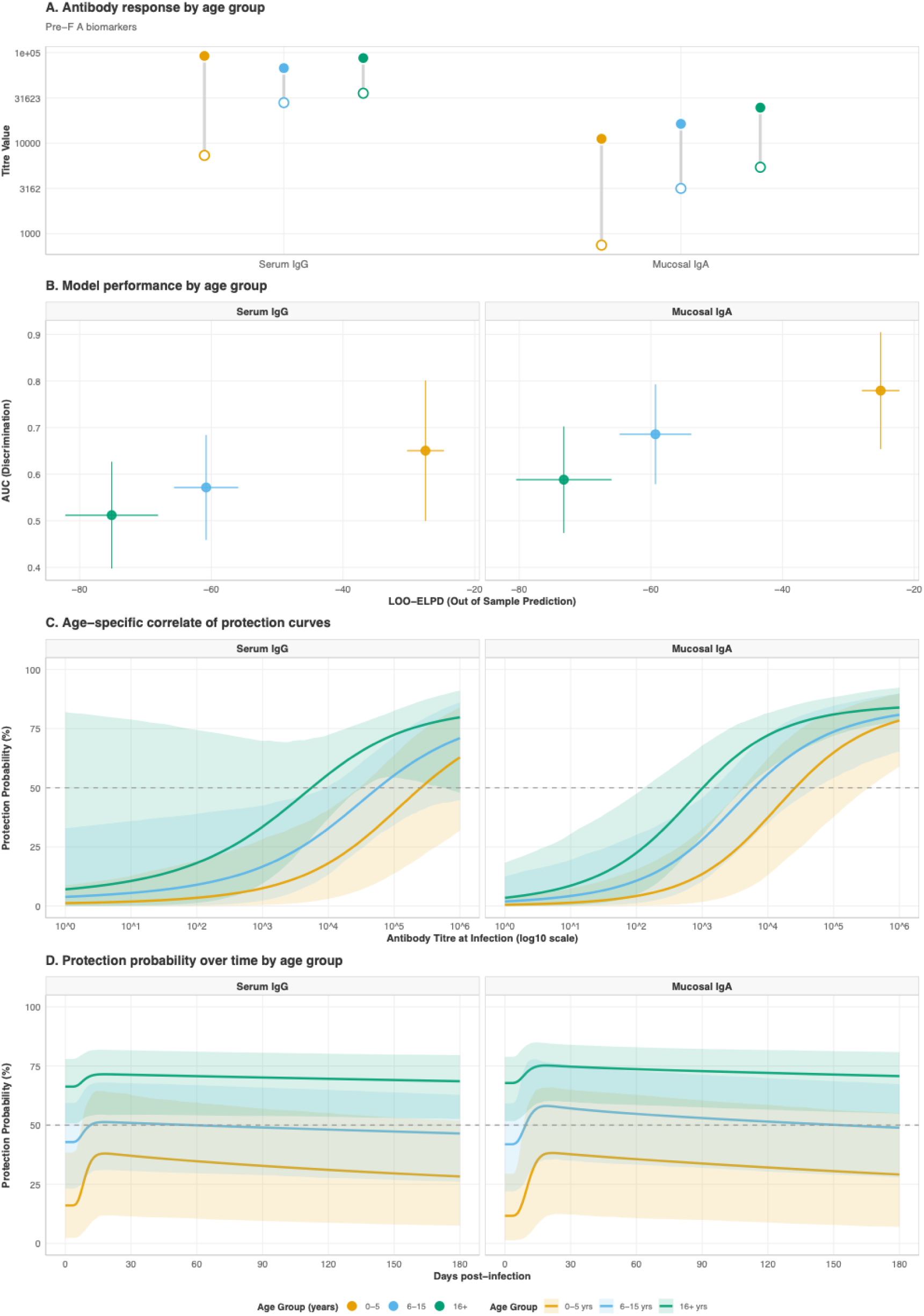
Age-stratified analysis of antibody responses and protective immunity for RSV-A PreF biomarkers. A. Antibody response magnitudes by age group, with baseline antibody titres at time of infection (hollow circles) and peak titres following infection (filled circles) for serum IgG and mucosal IgA antibodies to RSV-A PreF protein, stratified by age group (0-5 years in orange, 6-15 years in light blue, 16+ years in green). B. Model performance by age group. The scatter plot compares out-of-sample predictive accuracy (LOO-ELPD, x-axis) and discrimination ability (AUC, y-axis) for correlate of protection models fitted separately within each age stratum.. Vertical error bars show 95% confidence intervals for AUC; horizontal error bars indicate standard error of LOO-ELPD. C. Age-specific correlate of protection curves demonstrating the relationship between antibody titre at infection and probability of protection. D. Protection probability trajectories by age group following infection. The lines show the predicted probability of protection against reinfection over time (0-180 days post-infection) for serum IgG (left panel) and mucosal IgA (right panel) antibodies to RSV-A PreF. Solid lines represent the median posterior prediction for each age group, with shaded ribbons indicating 95% credible intervals.

Combining serum IgG and mucosal IgA antibodies to RSV-A PreF in a dual biomarker model yielded small improvements in predictive performance compared to either single biomarker alone (**Figure 3A**) with an increase in LOO-EPLD and discrimination; AUC = 0.73 (95% CrI 0.67–0.79). The corresponding plot for RSV-B antigens is given in **Supplementary Figure 5**, in which mucosal IgA to PreF remains the strongest correlate. In an alternative approach to evaluating the additive protective effect from two biomarkers, we estimated the average marginal gains from a 4- and 8-fold increase in the titre value at infection of either anti-PreF mucosal IgA alone *or* in both mucosal PreF IgA and serum PreF IgG. This demonstrated a greater increase in protection if both biomarkers increased compared to PreF IgA alone (e.g. 16% vs 22% increase in protection with a 4-fold increase in preF mucosal IgA vs both biomarkers respectively; **Figure 3C)**. Visualising both PreF IgG and IgA levels on a two-dimensional protection probability landscape further demonstrated the potential additive protective effect from the two biomarkers combined (**Figure 3D).** Correlation within all RSV-A IgG and IgA levels demonstrated a highly compartmentalised immune response, with correlation coefficients of 0.42-0.83 between serum IgG to the four RSV-A proteins and 0.37-0.75 between the four IgA biomarkers, but limited correlation between serum IgG and mucosal IgA levels to the same protein (0.02-0.28; **Figure 3E**). This further emphasises that the serum and mucosal antibody compartments provide largely independent information about immune protection (see **Supplementary Figure 7** for correlations for RSV-B).

### INFLUENCE OF AGE ON ANTIBODY KINETICS AND CORRELATES OF PROTECTION

Age influenced both antibody kinetics following infection and the strength of correlates of protection for RSV-A PreF IgA and IgG. Younger children (0-5 years) exhibited lower mean baseline antibody titres at the time of infection compared to older age groups (serum IgG: 7,323 AU/mL vs 35,598 AU/mL in 16+ years; mucosal IgA: 747 AU/mL vs 5,418 AU/mL), consistent with reduced cumulative exposure (**Figure 5A)**. Younger children also demonstrated the largest relative antibody boosting following infection, with mean fold-rises of 12.5-fold for serum IgG and 14.9-fold for mucosal IgA, compared to 2.4-fold and 4.5-fold, respectively, in adults (16+ years). However, we observed an antibody ceiling effect whereby boosting capacity declined with increasing baseline titres, even after accounting for age (**Supplementary Figure 8**). For serum IgG, the correlation between baseline titres and fold-rise varied by age group (slope: −1.42, −0.19, -0.09 for <5, 5–15, and ≥16 years, respectively), while mucosal IgA demonstrated more consistent ceiling effects across ages (slope: −0.27, −0.38, −0.28).

**Figure 5.**
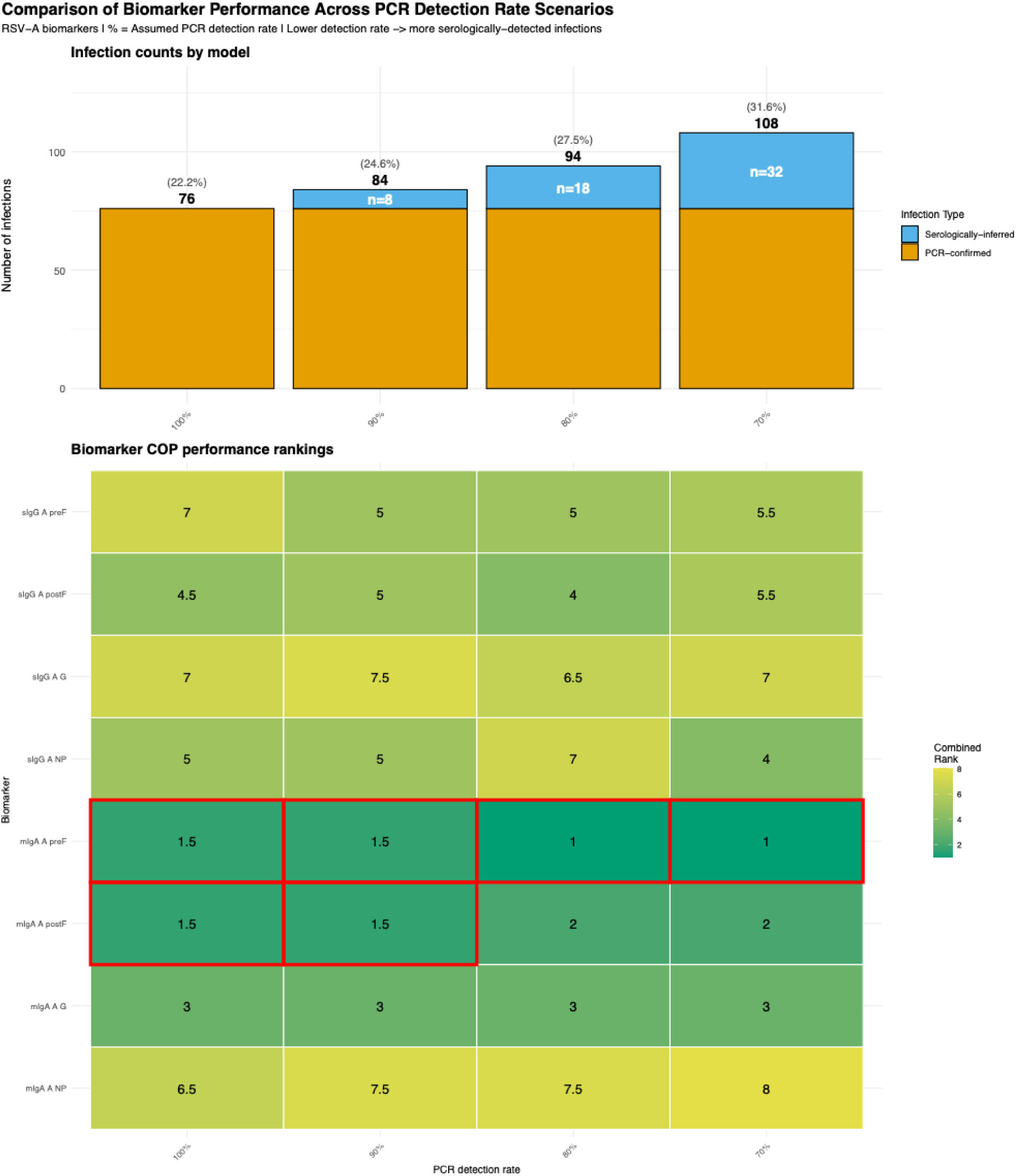
Comparison of biomarker performance across PCR detection rate scenarios for RSV-A. **Top.** shows the total number of inferred infections and infection type composition (PCR-confirmed in orange vs serologically-inferred in blue) for each model. Attack rates are shown in parentheses. **Bottom.** shows the combined performance ranking (average of AUC and LOO-ELPD ranks) for each RSV-A biomarker as a correlate of protection across the four models. Lower ranks (green) indicate better predictive performance, while higher ranks (red/orange) indicate poorer performance. Red boxes denote the top-ranked biomarker in each scenario.

Despite robust acute post-infection responses in young children, antibody titre-protection relationships showed marked age-dependent patterns. Mucosal IgA maintained superior discriminatory performance across all age groups (AUC: 0.78, 0.69, 0.58 for 0–5, 6–15, ≥16 years) compared to serum IgG (AUC: 0.66, 0.57, 0.51). The relatively greater predictive capacity of PreF mucosal IgA over serum PreF IgG in protection against RSV-A infection was most apparent in children <5 years old (AUC 0.78 vs 0.66, **Figure 5B**). In contrast, the predictive capacity of PreF IgA vs IgG in adults was more equivalent (AUC 0.58 vs 0.51). Intriguingly, younger children appeared to require higher titres to achieve equivalent protection: 50% protection corresponded to mucosal PreF IgA titres of 247,708 AU/mL in children <5 years versus 5,722 AU/mL in individuals ≥16 years. Converting post-infection antibody trajectories to protection probabilities via fitted correlate-of-protection curves revealed age-stratified dynamics, with younger children achieving 11–37% protection (depending on time post-infection) following an RSV-A infection episode, compared with 66–75% in older adults. These findings indicate that protection accumulates with age-related immune maturation but plateaus at approximately 75% at the population level, even in frequently exposed adults.

### ROBUSTNESS OF FINDINGS ACROSS PCR DETECTION RATES

To assess the robustness of our findings, we fitted models under four different assumptions about PCR detection rate (ρ): 100% (PCR-only, no missed infections), 90%, 80%. (base case), and 70% (**Figure 5A**). The superior predictive performance of mucosal IgA to PreF as a correlate of protection was robust to PCR detection rate assumptions (**Figure 5B**). PreF mucosal IgA ranked as the top-performing biomarker (combining AUC and LOO-ELPD rankings) in all of the four scenarios. Its discrimination performance (AUC) remained high across all models: 0.65 (ρ=100%), 0.70 (ρ=90%), 0.72 (ρ=80%), and 0.72 (ρ=70%). In contrast, serum IgG to PreF showed consistently lower discriminatory capacity (AUC: 0.60–0.65) while maintaining relatively stable rankings. Therefore, despite substantial differences in infection classification, our key findings remained consistent across all four detection rate scenarios.

## DISCUSSION

Using longitudinal samples, we measured serum IgG and mucosal IgA responses to multiple RSV antigens (PreF, PostF, G, and NP) and applied a Bayesian hierarchical modelling framework to jointly estimate antibody dynamics, exposure risk, and correlation of antibody titres at exposure with protection. Integration of PCR and serological surveillance suggested that 88% of RSV infections were asymptomatic in this high-endemicity setting, with attack rates declining from 53% in young children to 11% in older adults. We identified mucosal IgA to PreF viral protein as the strongest correlate of protection, with consistently higher predictive performance than serum IgG across all age groups. Younger children required a 43-fold higher mucosal IgA titre than adults to achieve equivalent protection (247,708 vs. 5,722 AU/mL for 50% protection), suggesting that immune maturation fundamentally alters antibody functionality beyond quantity alone. Furthermore, the greater predictive capacity of anti-PreF mucosal IgA over anti-PreF serum IgG was more marked in young children than in adults, where these two biomarkers were more equivalent correlates of protection. Despite robust antibody boosting following natural infection, population-level protection plateaued at approximately 79% even among frequently exposed adults, explaining RSV’s capacity for lifelong reinfection.

Our finding that 88% of RSV infections were asymptomatic aligns with observations from other African community settings, including reported asymptomatic infection percentages of 67% in South Africa^23^ and ∼57% in Kenya^24^. This substantial undetected transmission suggests RSV circulates far more widely than symptomatic surveillance indicates. Consequently, efficacy trials using symptomatic endpoints cannot accurately estimate protection against infection, potentially missing transmission-blocking effects entirely. For vaccines intended to reduce community transmission, comprehensive surveillance, including serology, will be essential for assessing their true impact.^25,26^

Mucosal IgA to PreF as the strongest predictor of protection (AUC: 0.72, 95% CrI 0.66–0.78) against RSV infection, outperforming serum pre-F IgG (AUC 0.65, 95% CrI 0.59–0.73). To contextualise this finding, AUC > 0.70 indicates acceptable discriminatory performance^27^, approaching previously reported correlate strengths against infection observed for serum IgG PreF (AUC 0.76–0.77)^28^ and for influenza HAI (AUC of 0.76).^29^ This finding aligns with previous evidence that nasal IgA provides more effective protection than serum neutralisation^30^, likely reflecting the importance of antibody responses at the site of viral entry in the respiratory mucosa. The modest improvement when combining PreF mucosal IgA with serum IgG in a dual biomarker model (AUC 0.73, 95% CrI 0.67–0.79), suggests that mucosal immunity provides substantial additional predictive information. Weak correlations between serum IgG and mucosal IgA to the same protein (r = 0.02–0.28), indicate largely independent compartment responses, consistent with previous studies.^31,32^ This contrasts with strong correlations between different antigens within the same compartment (r = 0.42–0.83). Together, these findings also suggest that protection derived from serum IgG and mucosal IgA may be relatively independent from each other. Critically, the importance of anti-PreF mucosal IgA as a correlate of protection remained consistent when the analysis was restricted to PCR-confirmed infections, demonstrating robustness across detection methods. We emphasise that these findings demonstrate an associative correlation with protection, not a causal one, as our observational design cannot definitively establish that mucosal IgA directly causes protection; it may instead serve as a biomarker of other immune mechanisms.^33^

A notable observation from our study is the discordance between functional antibody measurements and protective associations. While serum IgG to PreF correlated more strongly with live virus neutralisation than mucosal IgA (Pearson’s r = 0.74 vs. 0.43), mucosal IgA better predicted protection. This discrepancy may partly reflect technical factors, including greater variability in nasal sampling and quantification of mucosal antibodies, or the limited ability of neutralisation assays to recapitulate the physiochemical and cellular conditions of the respiratory mucosa. Mucosal IgA may also mediate protection through mechanisms not captured by standard neutralisation assays, including immune exclusion at the epithelial surface, viral aggregation, and intracellular neutralisation.^34,35^ In addition, secretory IgA may have enhanced avidity, multivalency and resistance to proteolysis within the mucosal environment, enabling effective antiviral activity even in the absence of high measured neutralisation titres. Future studies should test these hypotheses directly using functional assays that better replicate mucosal conditions and effector functions. Collectively, these findings suggest that traditional serum neutralising antibody titres, while good correlates of disease severity, may be insufficient to define protection against infection at mucosal entry sites. Moreover, as population immunity becomes increasingly vaccine-derived in certain groups, the relative contributions and correlates of mucosal versus systemic immunity may diverge from those defined in natural infection.

Our analysis of the antibody kinetics revealed that mucosal IgA responses to surface glycoproteins (PreF, PostF, and G) demonstrated stronger boosting (4.7–6.3-fold) and more sustained persistence (220–235 days above a 2-fold rise) than serum IgG responses. This superior durability of mucosal responses to surface glycoproteins contrasts with prior assumptions that mucosal antibodies are inherently short-lived, owing to the transient nature of mucosal plasma cells.^30,36–38^ The exception was G protein, which showed exceptional serum IgG boosting (8-fold) with typical waning kinetics (177 days). In contrast, the internal NP protein showed modest responses in both compartments, consistent with limited exposure at mucosal surfaces.

We found that younger children (0–5 years) demonstrated robust antibody boosting (12.5-fold serum IgG, 14.9-fold mucosal IgA) but required substantially higher titres for equivalent protection - a 43-fold increase for 50% protection compared to individuals 16 years and older. This suggests that antibody quantity alone does not determine protective capacity, with qualitative differences fundamentally altering the titre-protection relationship across ages. Several mechanisms could explain this age-dependent protective capacity. Children may have lower antibody affinity due to fewer rounds of affinity maturation;^39,40^ or they may target less protective epitopes than adults, as has been observed for influenza.^41,42^ Age-related differences in mucosal architecture and cellular immunity may also modulate effective antibody concentrations.^43^ These findings challenge immunobridging approaches: a titre correlating with 65% protection in adults might provide only 25% protection in young children. Vaccines already licensed for use in younger infants may not provide the same protection if used on other age groups (e.g. school-age children) when relying solely on infant-derived protection correlations; age-specific correlates should be established rather than extrapolated. While our study focused on humoral immunity, these age-dependent differences likely also reflect contributions from T cell-mediated responses, which mature alongside mucosal antibodies over repeated exposure.

We also observed an antibody ceiling effect whereby boosting capacity declined with increasing baseline titres. For serum IgG, baseline-fold-rise correlations varied by age (slope: −1.42, −0.19, -0.09) while mucosal IgA showed consistent ceiling effects across ages (slope: −0.27, −0.38, −0.28).). This likely reflects antibody-mediated feedback inhibition, memory B cell exhaustion, and altered germinal center dynamics.^44^ Practical implications of this include: (1) boosting timing should be optimised to the period of antibody waning; (2) high-dose strategies may have limited benefits if reflecting B cell activation limits and (3) sequential vaccination schedules may require strategic spacing.

We also found that population-level protection plateaued at 75% even in highly immune adults, while younger children achieved only 11–37% protection depending on time post-infection. Interestingly, this plateau falls around the herd immunity threshold for RSV, which requires (1 – 1/R0) of the population to be fully immune (100% protection). With an estimated R0 of 2–3 for RSV^45^, achieving herd immunity would require 50–67% of the population to have full protection. Given the study results, which indicate that most adults achieved only 70% individual protection at peak antibody levels, achieving persistent population-level herd immunity through natural infection would require 77% or more of the population to be similarly immune. This may explain RSV’s ability to cause lifelong reinfections in a non-vaccinated setting.

These findings have implications for vaccine development. Currently licensed RSV vaccines primarily induce systemic IgG responses and demonstrate 70–90% efficacy against clinical disease^3,4,6^. While these vaccines effectively prevent severe disease, the stronger association between mucosal IgA and protection against infection raises the question of whether mucosal immunity may confer additional benefits in preventing infection and reducing transmission. Vaccines optimised for mucosal delivery could theoretically provide superior protection against infection by directly eliciting mucosal IgA.^46,47^ However, vaccine-induced immunity may differ fundamentally from natural immunity^48,49^ and mucosal vaccines have historically faced significant challenges in gaining licensure.^50,51^ Therefore, we suggest that mucosal approaches could initially serve as complementary strategies to existing vaccination programmes. Vaccine trials should incorporate mucosal antibody measurements alongside serum assays, and transmission should assess whether current vaccines provide indirect protection. The practical challenges of measuring mucosal IgA require nasal sampling and a lack of standard assays^52^, currently limit its utility as a routine correlate.

Several limitations warrant consideration. First, this study captured one RSV season in rural Gambia with RSV-A predominance (96%), limiting generalisability to other populations, season and serotype-specific assessment of immunity. Findings from our high-transmission setting may not apply to lower-transmission high-income countries where exposure patterns, nutritional status, co-infections and many other factors differ. Second, antibody measurements in arbitrary units limit cross-study comparability as the titre value we report cannot be directly compared to other studies without further assay standardisation. Third, we primarily measured antibody quantity but not functional properties such as neutralisation potency, avidity, or epitope specificity, which may partially explain why children require higher titres for equivalent protection. Fourth, our observational design cannot establish causality between mucosal IgA and protection. Finally, the limited follow-up (12 months) constrains the characterisation of long-term antibody durability.

Nevertheless, this household cohort study in a sub-Saharan African population provides comprehensive insights into RSV immunity in regions with the highest disease burden. Future research should focus on mechanistic studies of mucosal IgA-mediated protection, establishing age-specific correlates against RSV infection through efficacy trials, and standardisation of mucosal antibody assays. These insights should inform vaccine development while setting realistic expectations about achievable protection given biological constraints on RSV immunity.

## METHODS

### PARTICIPANT RECRUITMENT AND DATA COLLECTION

The set up of the TRANSVIR household study has been previously described in detail.^53^ In summary, following community and household sensitisation, interested families were invited to the MRCG clinical site for consenting and screening. Recruited households ranged in size from 6 to 10 household members, including at least one adult and one child. Written consent or a thumbprint was obtained from all participants >18 years, whilst assent was obtained from participants aged 12–18 years. Consent was obtained from the parents or guardians for participants under 12 years. Participants were followed up for 12 months with weekly combined throat and nose flocked swabs (TNS) collected into RNAprotect cell reagent (Qiagen).

Participants were asked about their symptoms in the 7 days preceding each sampling time point. At enrollment, and then 6-monthly (to a total of 3 scheduled) participants attended the clinic for venous blood sampling for serum separation and collection of nasal lining fluid using a synthetic absorptive matrix (SAM) strip. If participants developed symptoms compatible with an upper respiratory tract virus infection, they were advised to contact the study team, after which an unscheduled visit would occur. At an unscheduled visit, the entire household would be swabbed, with RT-PCR testing performed contemporaneously. Participants with a positive SARS-CoV-2 RT-PCR at an unscheduled visit would then have an additional venous blood sample collected 28 days post-infection. Full details of the cohort design, sample collection, and laboratory methods have been previously published.^21,53^

### ETHICS APPROVAL

Study approval was given by the joint Gambia Government and Medical Research Council Unit, The Gambia (MRCG) Ethics committee, and the London School of Hygiene and Tropical Medicine ethics committee (LEO project ID 22556). The protocol was registered at clinicaltrials.gov (NCT05952336).^53^

### RSV LUMINEX IMMUNOASSAY PROTOCOL

A custom in-house multiplex immunoassay was developed using Luminex xMAP technology (DiaSorin). RSV antigens (RSV-A Pre-Fusion (ProteoGenix, product code: PX-P6126), RSV-B Pre-Fusion (SinoBiological, product code: 40832-V08B), RSV-A Post-Fusion (SinoBiological, product code: 11049-V08B), RSV-B Post-Fusion (SinoBiological, product code: 40999-V08H), RSV-A G protein (SinoBiological, product code: 11070-V08H2), RSV-B G protein (SinoBiological, product code: 13029-V08H, RSV-A Nucleoprotein (SinoBiological, product code: 40821-V08E and RSV-B Nucleoprotein (SinoBiological, product code: 40822-V08F)), were coupled to distinct bead regions. Pre-Fusion, Post-Fusion and Nucleoprotein antigens were coupled to specific beads using a Luminex xMAP antibody coupling kit (product code 40-50016, DiaSorin) and the G proteins were coupled to specific beads using the AnteoTech Activation Kit for Multiplex Microspheres (product code A-LMPAKMM). Coupling procedures followed the manufactures instructions. Coupled beads were stored at 4°C in PBS with 1% BSA and 0.05% sodium azide.

Microspheres coupled to each RSV antigen were diluted to 50 microspheres/ul in PBS with 0.05% Tween-20 and 1% BSA. Serum samples were diluted 1:300 and mucosal samples were diluted 1:10. Diluted samples were incubated with the microsphere antigen mix in low-binding 96-well microplates for 30 minutes. Following incubation, plates were washed three times with 300 µl/well of PBS 0.05% Tween20 using a 405 TS BioTek plate washer. Detection is performed using PE-conjugated mouse anti-human IgG antibodies (product code: ab99761, Abcam) or PE-conjugated goat anti-human IgA antibodies (product code: ab99909, Abcam). Plates read on a Luminex xMAP INTELLIFLEX system (DiaSorin). Luminex reported the signal as Medium Fluorescence Intensity (MFI). MFI values were converted to arbitrary units by interpolating sample results from a standard curve.

After excluding duplicates, missing data, and samples without paired IgG/IgA measurements, 926 serum samples from 342 individuals in 52 households remained for analysis (Supplementary Methods Section 1).

### RSV LIVE-NEUTRALISATION ASSAY

#### Sample selection criteria for live microneutrsliation assay

To validate the relevance of our biomarkers as antigenic targets, we measured live virus neutralization in representative subsets of 97 serum samples and 97 mucosal samples, which were selected to evenly span the distribution of antibody titre values.

#### Virus cultivation and titration

RSV BT2a and Hep-2 cells were kindly gifted by Lindsay Broadbent, University of Surrey. The origin and characterisation of the clinical isolate RSV BT2a has been previously described^54^. RSV BT2a was propagated in Hep-2 cells and titres were determined by immunostaining plaque assay using Vero E6 cells as by standard method^55^. Briefly, Hep-2 cells (107 cells seeded into T175 flask the day before) were infected with RSV BT2a at an MOI of approx. 0.005. Cells were washed once with DMEM (Sigma; D6429), then 5 ml virus inoculum made up in DMEM was added to each T175 flask and incubated at at 37° C, 5% CO2, rocking gently for 2h. DMEM + 1% FCS (Biosera; FB-1001/500) was added to each flask for 24h, then media was removed and cells washed with PBS. DMEM + 1% FCS was re-added and cells incubated for 3-4 days until CPE was evident. Virus was harvested by scraping attached cells into the overlay medium, then the suspension was collected into centrifuge tubes, sonicated for 10 min, and cleared by centrifugation (460 x g for 10min at 4°C). Supernatant was aliquoted, snap-frozen and store in liquid nitrogen.

#### High-throughput live virus microneutralisation assay

High-throughput live virus microneutralisation assays were performed as previously described^56^ adapting the established assay pipeline initially setup for SARS-CoV-2 for RSV. In brief, Vero E6 cells (Institute Pasteur) at 90-100% confluency were infected with RSV BT2a in 384-well format in the presence of serial dilutions of patient serum samples and Nirsevimab as positive control. The assay was further miniaturised to 1536-well format for testing nasal lining fluid samples, which are limited in volume. After infection, cells were fixed with 4% final Formaldehyde, permeabilised with 0.2% TritonX-100 and 3% BSA in PBS (v/v), before staining for RSV using the Alexa488 conjugated MAB88262 antibody at 1:3000 and cellular DNA using DAPI.

Whole-well imaging was carried out at 5x or 10x, respectively for 384 and 1536 well plate formats, using an Opera Phenix Microscope (Perkin Elmer) and fluorescent areas and intensity were calculated using the Phenix-associated software Harmony (Perkin Elmer). Inhibition was estimated from the measured area of infected cells/total area occupied by all cells for the 5x imaging, whereas counting of nuclei and infected cells was automated for the 10x imaging of 1536 plates. In both cases, viral inhibition was expressed as percentage of maximal (virus only control wells). The inhibitory profile of each serum sample was estimated by fitting a 4-parameter dose response curve executed in SciPy. Neutralising antibody titres are reported as the fold-dilution of serum required to inhibit 50% of viral replication (IC50), and are further annotated if they lie above the quantitative range, below the quantitative range but still within the qualitative range (i.e. partial inhibition is observed but a dose-response curve cannot be fitted because it does not sufficiently span 50% inhibition), or if they show no inhibition at all.

### SEROLOGICALLY-DEFINED INFECTIONS AND SENSITIVITY ANALYSIS

To identify RSV infections missed by PCR surveillance, we developed a rank-based serological method applied to PCR-negative individuals. For each of 16 biomarkers, we computed log10 fold-changes between baseline and 6-month follow-up (the interval during which all PCR-confirmed infections occurred): 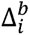 for individual i and biomarker b. Within each biomarker, we ranked all PCR-negative individuals by their titre changes (rank 1 = highest fold-rise) and then calculated their average rank across all biomarkers 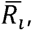 assuming equal weighting. Individuals with the lowest rank score show the broadest serological evidence of infections across all biomarkers. To determine how many PCR-negative individuals to classify as serologically defined infections, we specified *ρ*, the detection rate of the PCR sampling protocol in the study. We assumed a base-case value of *ρ* = 0.8 for the primary analysis, implying 80% of infections were detected by the PCR-sampling method. We selected the top *N_missed_* = *ρ*/(1 - *ρ*)*N_PCR_*_+_, where *N_PCR_*_+_ the number of PCR-negative individuals, with the lowest aggregated rank scores 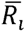 and classified these as serologically-defined infections. We conducted a sensitivity analysis *ρ* = {1.0, 0.9, 0.8, 0.7}, varying to assess robustness to uncertainty about the true burden of missed infections (Supplementary Figure S9 and Supplementary Methods Section 3.1).

### BAYESIAN HIERARCHICAL MODELING FRAMEWORK

We developed a Bayesian hierarchical model to jointly estimate antibody kinetics following RSV infections and identify correlates of protection against infection. The framework integrates three key components: (1) an antibody kinetics model describing waning and boosting dynamics following infection or vaccination, (2) a logistic correlate of protection model relating antibody levels to infection risk given exposure, and (3) an exposure model accounting for household transmission intensity. (Full details in Supplementary Methods Section 3.2)

Antibody trajectories were modelled on the log_₁₀_ scale, with linear waning prior to exposure and parametric boosting post-exposure using power-function formulations adapted from Teunis et al.^57^ Boost kinetics parameters included group-level random effects stratified by age. For each individual, we predicted titre values at observed time points and compared these to measured values using Gaussian likelihoods with left-censoring at the limit of detection (Supplementary Methods Section 3.2).

The correlate of protection, that is the relationship between pre-infection antibody titre and infection risk, was modelled using a logistic function with hierarchical effects. For infected individuals, we used model-predicted titres at infection time; for non-infected individuals, we used mid-season control titres (when 50% of surveillance infections occurred). The logistic function which characterized the correlate of protection was given by

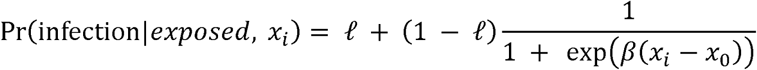

where *β* represents the protection gradient (slope), *x*_0_ is the protective titre threshold (50% protection level),) and *ℓ* is the lower asymptote representing residual infections risk at high titres. Both *β* and *ℓ* included group-level random effects stratified by age, allowing protection curves to vary across age groups.

To account for uncertain exposure status among PCR-negative individuals, we modelled individual exposure probability, as a function of household exposure level (number of household members with confirmed infections) using a logistic regression. The observed infection outcome was then modelled as a mixture: unexposed individuals contribute a negligible infection probability, while exposed individuals contribute an infection probability determined by their pre-infection titre and the protection curve (Supplementary Methods Section 3.2).

## PRIORS AND MODEL FITTING

Weakly informative prior distributions were specified for all parameters (**Supplementary Methods Section 3.2**). Models were fitted separately for each biomarker and wave combination, and we also developed dual-biomarker models with multivariate logistic protection functions including interaction terms (**Supplementary Methods Section 3.2**).

For each biomarker model, we ran 4 Markov chains for 2,000 iterations (1,000 warmup) in Stan (version 0.5.3) using Hamiltonian Monte Carlo sampling via cmdstanr in R version 4.2.3.

Convergence was assessed using the Potential Scale Reduction Factor (all parameters <1.1) and inspection of trace plots. Model comparison was performed using leave-one-out cross-validation (LOO-CV) and area under the ROC curve (AUC) for discriminating infections from non-infections. Posterior distributions were summarised using median and 95% credible intervals. Full model code is available at https://github.com/ccgh-idd/cop-transvir-rsv. (**Supplementary Methods Section 4**).

## Supporting information

Supplementary results

## Data Availability

All data produced are available online at https://github.com/ccgh-idd

## Acknowledgements

We are extremely grateful to all the TransVir participants who gave up their time and engaged so fully with the study. We also acknowledge the wider field TransVir field team members for their dedication and hard work during the field study. The study was funded by a United Kingdom Research and Innovation Grant (No. MC_PC_19084) and by the Medical Research Council (grant MR/Y004450/1). DH is funded by the Einstein Foundation Berlin (EPP-BUA-2022-697). SF is funded by the Einstein Foundation Berlin as an Einstein BUA Strategic Professor (EPP-BUA-2022-697, https://www.einsteinfoundation.de/en/). MYW and GD are supported by the Francis Crick Institute, which receives its core funding from Cancer Research UK (CC2230), the UK Medical Research Council (CC2230), and the Wellcome Trust (CC2230). LG and TC are supported by Grant-in-aid funding to the Emerging Pathogen Serology Group at Porton Down, UK Health Security Agency. BBL is supported by a Wellcome Trust Clinical Research Training Fellowship.

## SUPPLEMENTARY FIGURES

**Supplementary Figure 1.**
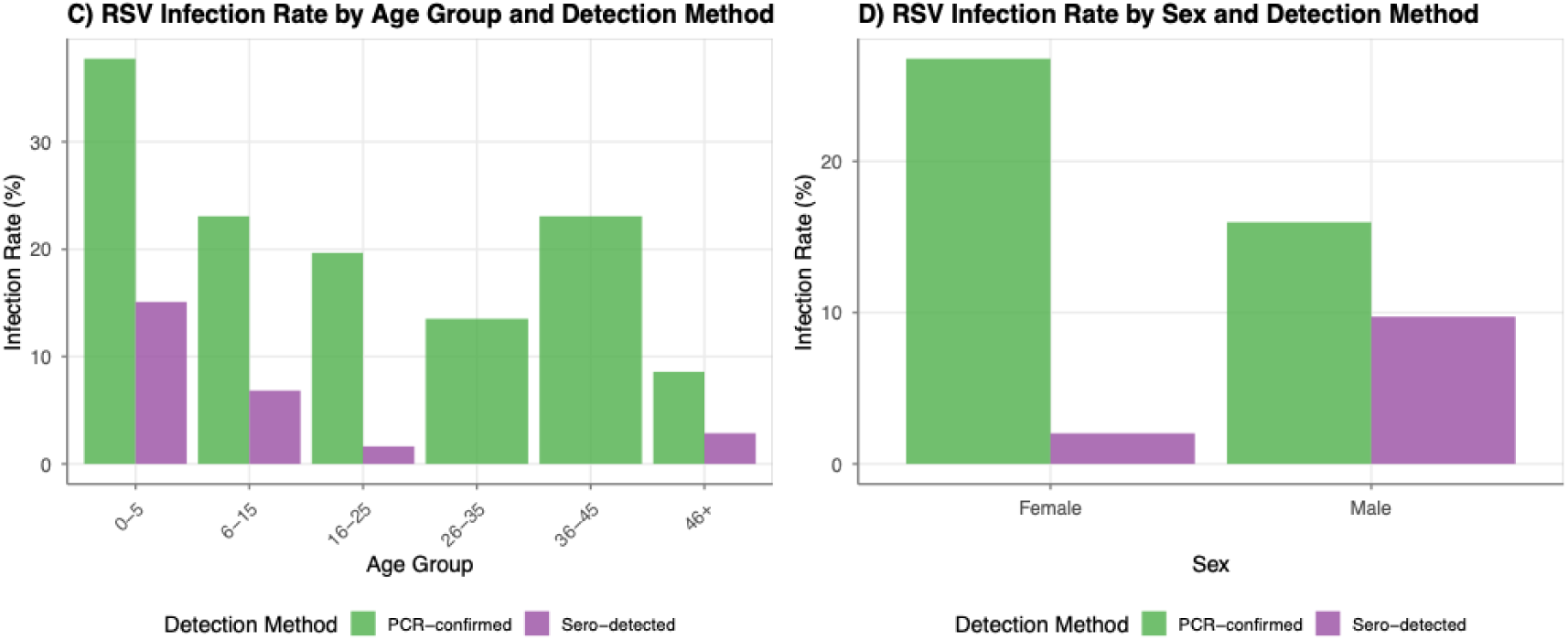
Demographic distribution of RSV infections stratified by detection method. Left panel shows infection rates across age groups, comparing PCR-confirmed infections (green) detected through active surveillance testing versus serologically-detected infections (purple) identified through antibody responses in the absence of PCR confirmation. Right panel displays infection rates stratified by sex for both detection methods

**Supplementary Figure 2.**
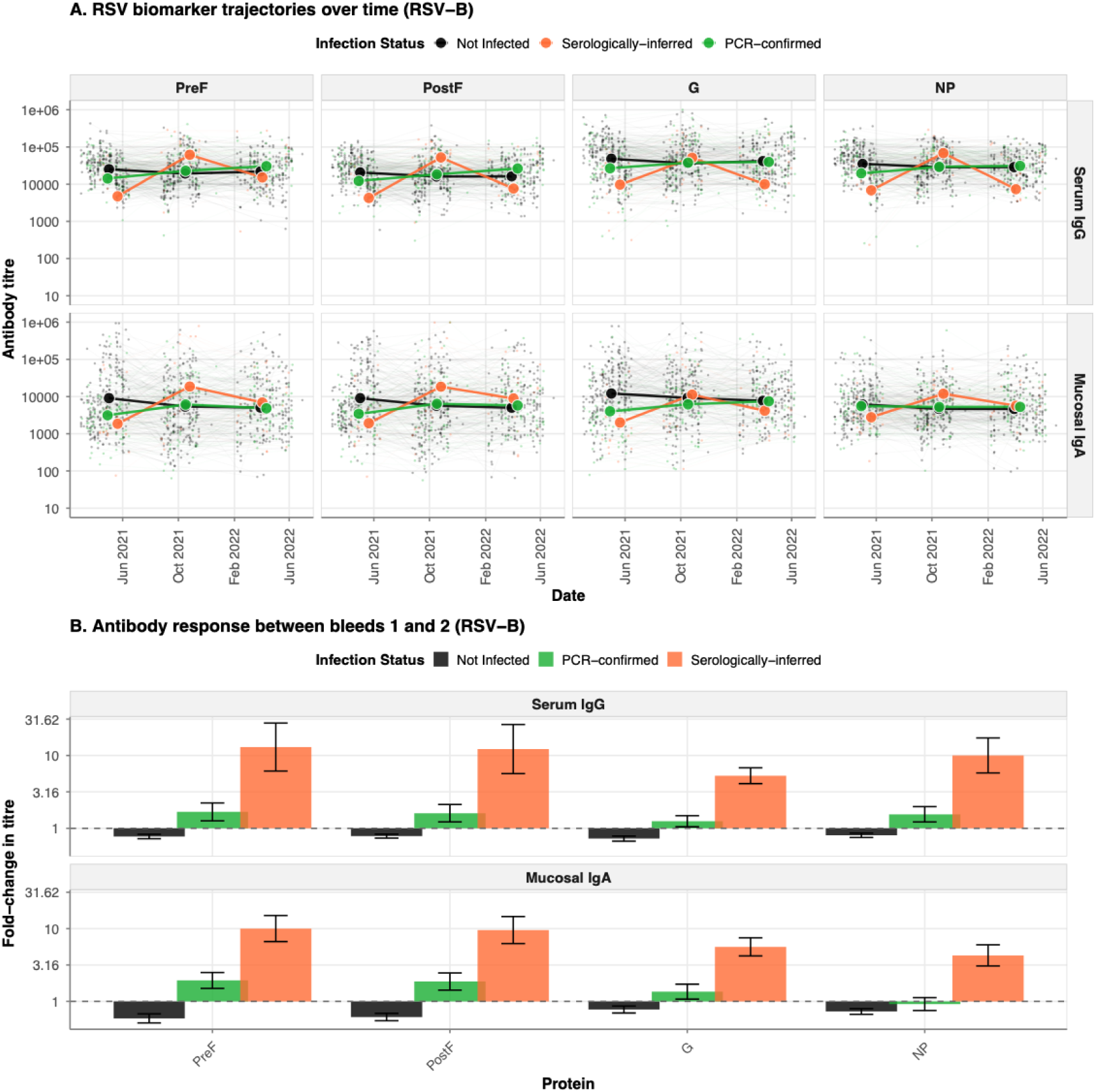
(A) Longitudinal trajectories of serum IgG (top row) and mucosal IgA (bottom row) antibody responses against four RSV-B proteins (PreF, PostF, G, NP) over time. Individual participant trajectories are shown as thin lines with low opacity, colored by infection status: not infected (black), sero-detected infections (orange), and PCR-confirmed infections (green). (B) Mean fold-change in antibody titres between the first bleed (pre-epidemic baseline) and second bleed (post-epidemic) for serum IgG and mucosal IgA responses to RSV-B proteins. Bars represent mean fold-change (log10 scale) stratified by infection status, with error bars indicating standard error.

**Supplementary Figure 3.**
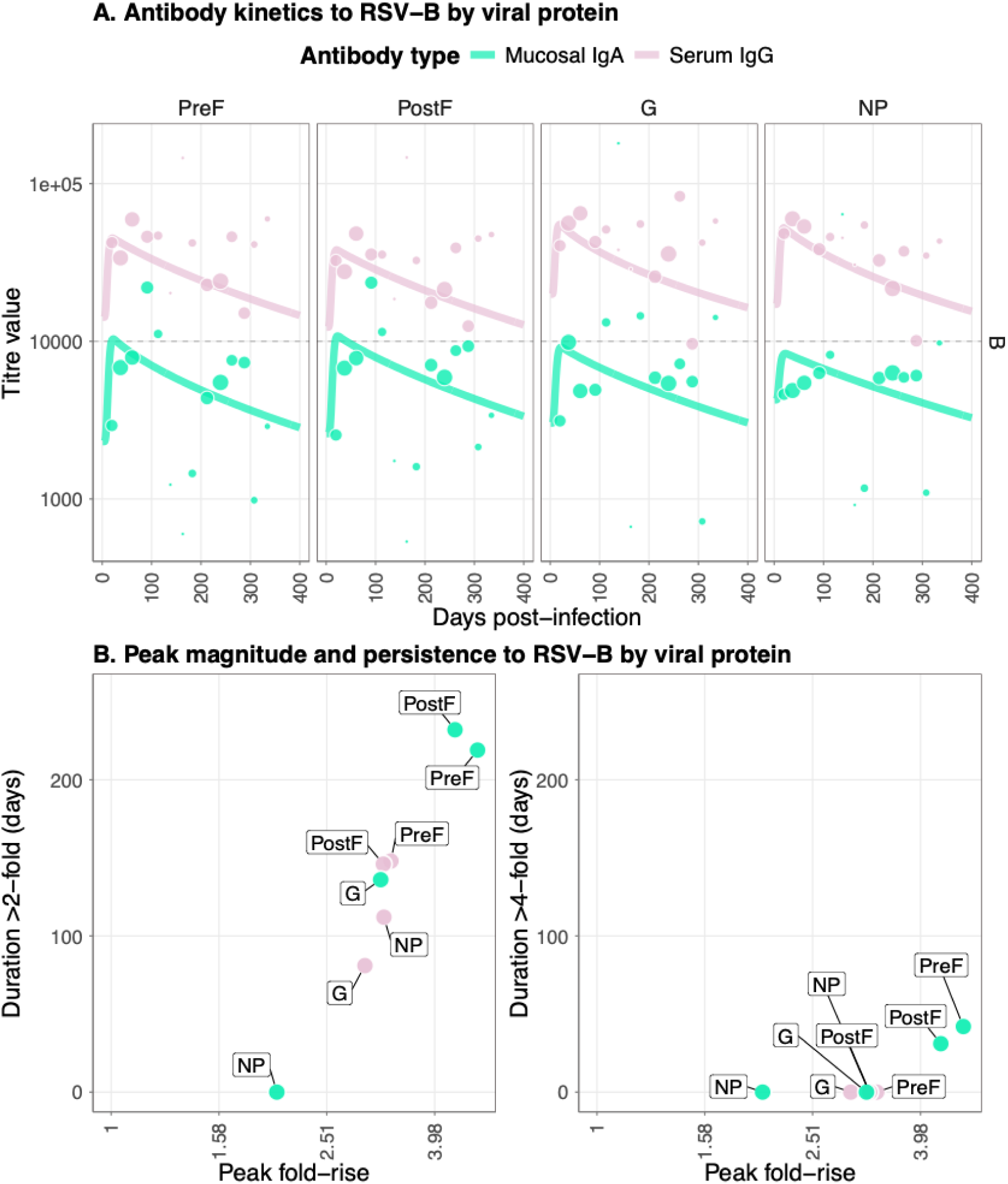
Antibody kinetics and immune responses for RSV-B following infection. (A) Post-infection longitudinal antibody titers for four viral RSV-B proteins (PreF, PostF, G, and NP) across different antibody types; serum IgG and mucosal IgA. Lines show the median posterior predictive fit from the fitted Bayesian model, and the points show the observational titre data, with the size correlating with the sample size for that bin. (B) Peak antibody (x axis) and persistence measured as duration above a 2-fold (left panel) and 4-fold titre rise (right panel) in days. Data points show median posterior values for measurements of four viral RSV-B proteins (PreF, PostF, G, and NP) across different antibody types: serum IgG and mucosal IgA.

**Supplementary Figure 4.**
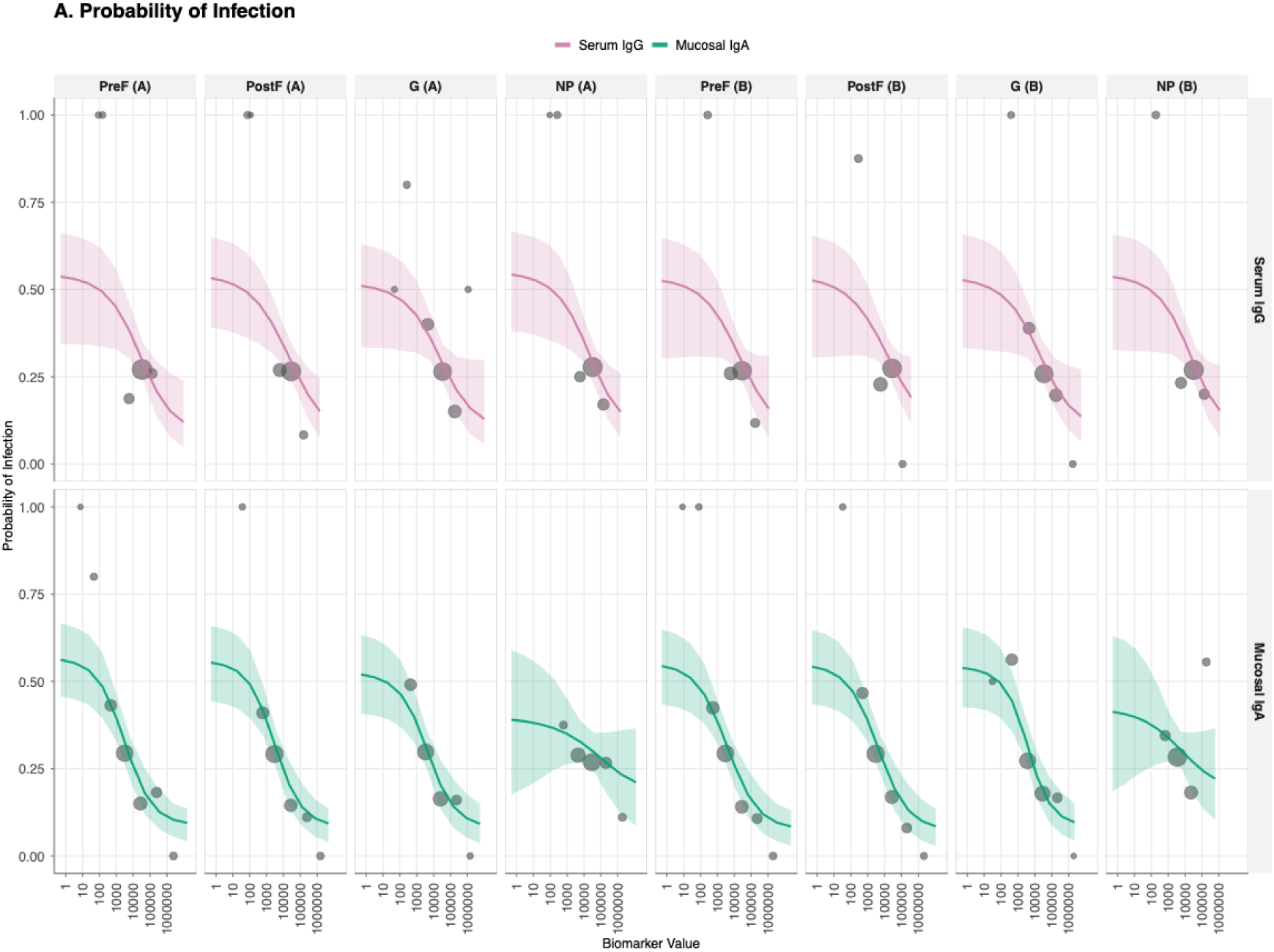
Correlate of protection (CoP) curves showing the relationship between antibody titre at infection and probability of protection from RSV infection for all 16 biomarker-antigen combinations. Serum IgG, top row; mucosal IgA, bottom row and columns are viral antigen target (PreF, PostF, G, and NP for both RSV-A and RSV-B strains). The solid green line represents the mean estimated probability of protection given exposure to infection as a function of antibody titre, with shaded ribbons indicating 95% credible intervals. Background histograms show the distribution of antibody titres at infection for infected individuals (orange) versus non-infected individuals (gray).

**Supplementary Figure 5.**
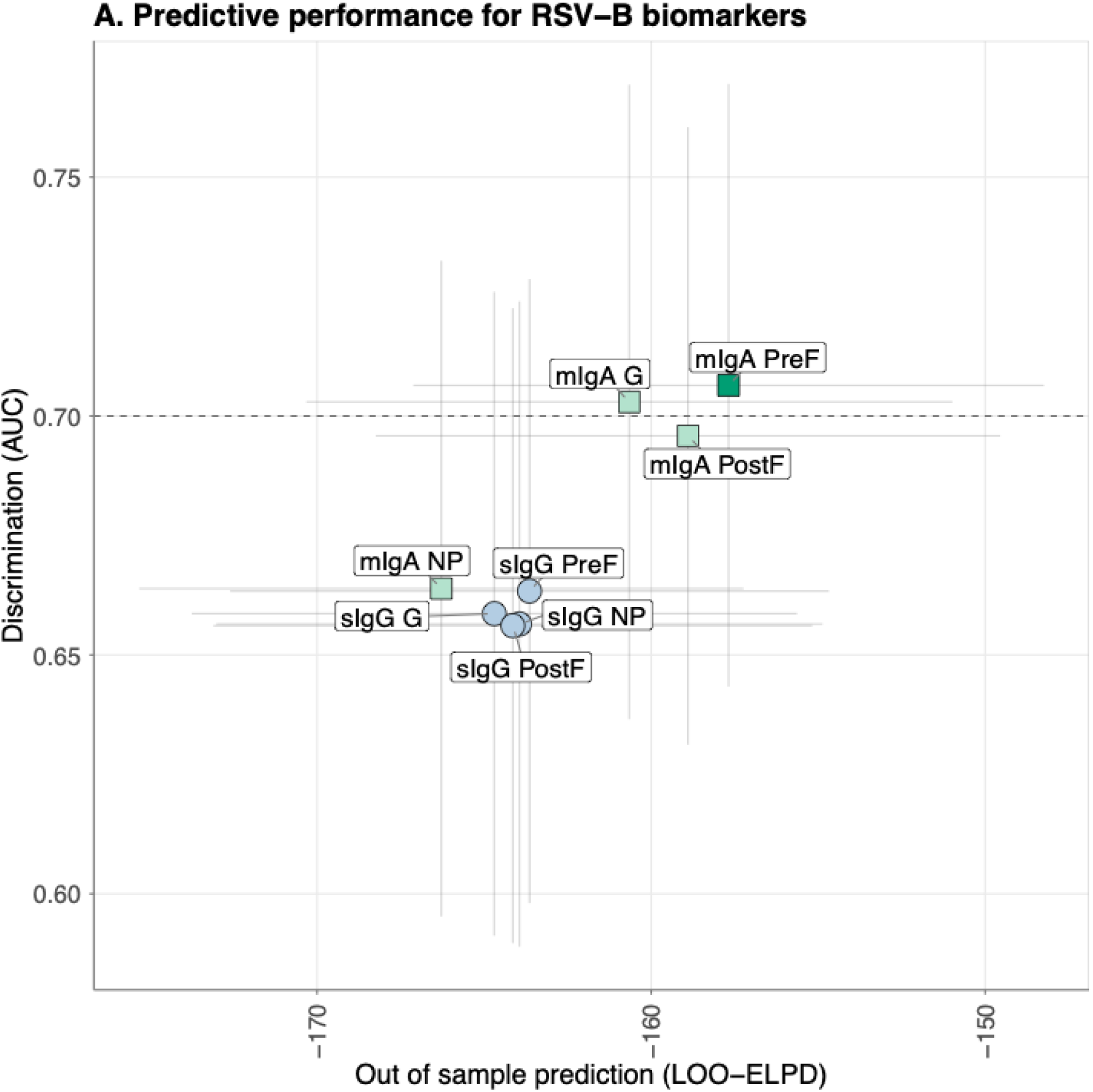
Comparison of single and dual biomarker models for predicting protection against RSV infection. Model performance comparison across single biomarker models and the dual biomarker model, defined by out-of-sample predictive accuracy (LOO-ELPD, x-axis) and discrimination ability (area under the ROC curve, AUC, y-axis). Circles indicate serum IgG models, squares represent mucosal IgA models, and the triangle denotes the dual biomarker model combining serum IgG and mucosal IgA to RSV-B PreF. The best-performing model within each biomarker class is highlighted with darker shading. Error bars show the standard error of LOO-ELPD (horizontal) and 95% confidence intervals for AUC (vertical). The dashed horizontal line indicates an AUC of 0.7.

**Supplementary Figure 6.**
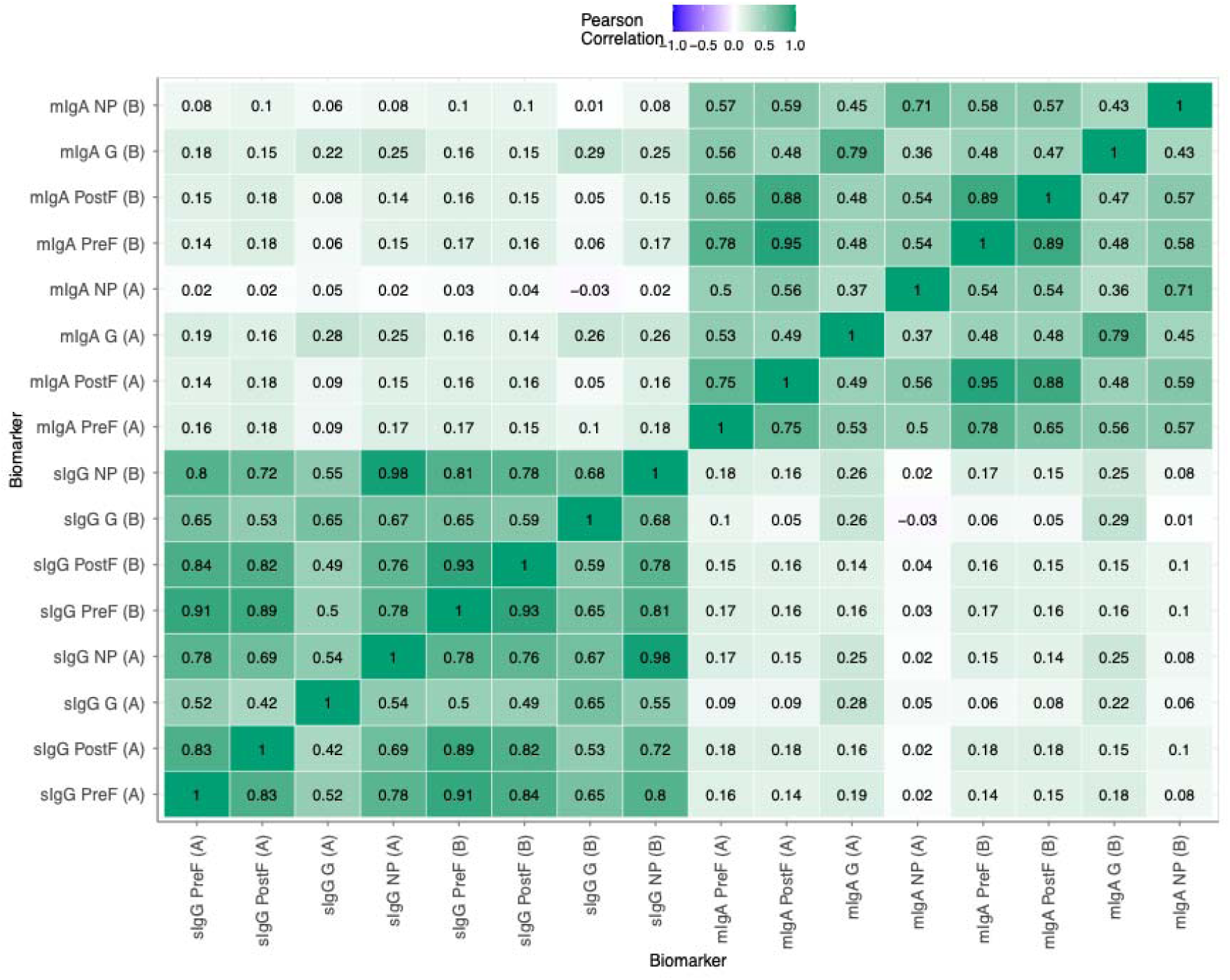
Pearson correlation coefficients between RSV-A and RSV-B variant biomarkers at the time of infection. Colour intensity represents correlation strength, ranging from blue (weak correlation) through white (no correlation) to green (strong positive correlation).

**Supplementary Figure 7.**
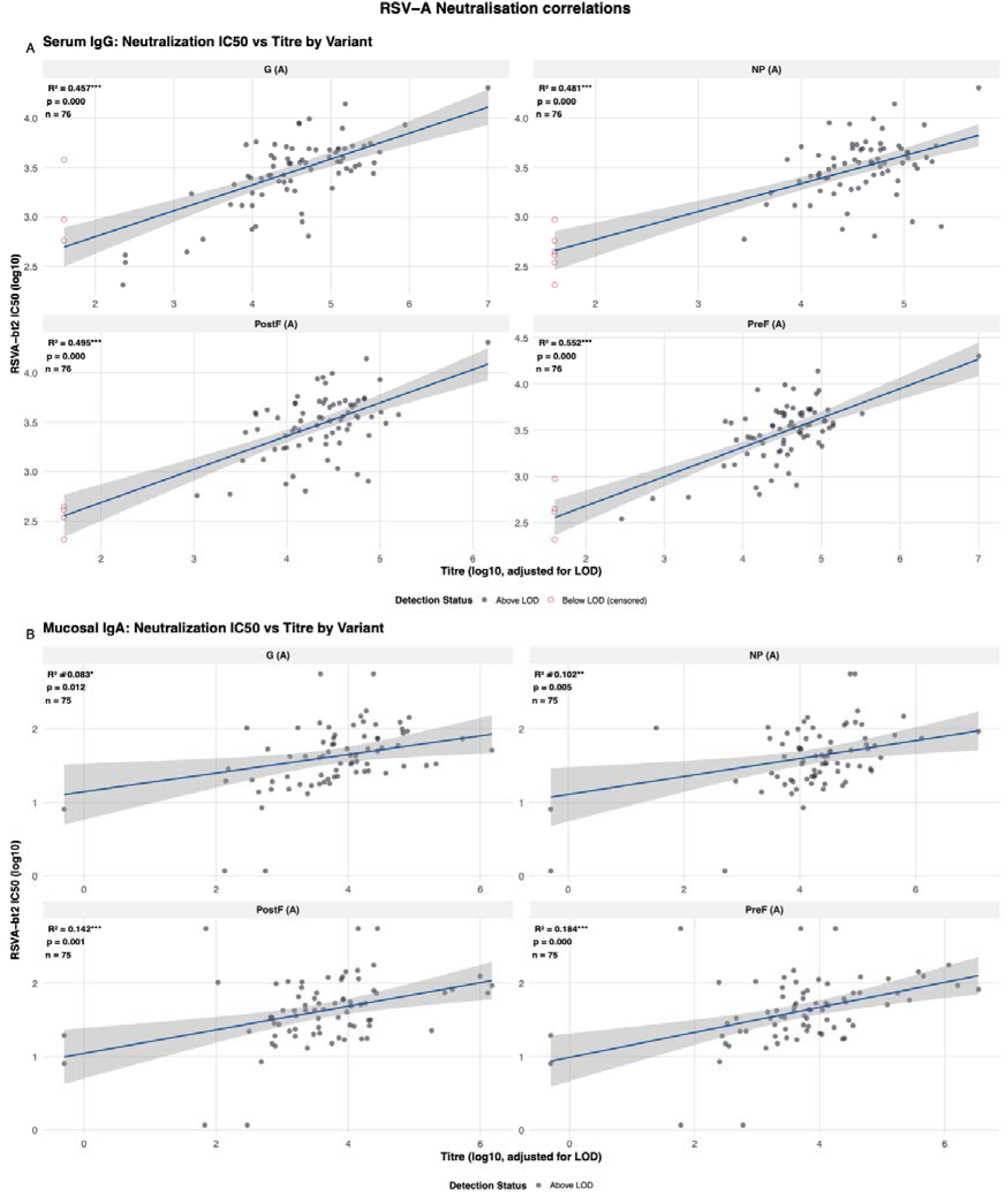
Correlation between neutralization capacity and ELISA binding antibody titres, with focus on RSV-B antigens. (A) Scatter plots showing serum IgG ELISA titres versus neutralization IC50 for all RSV-A variants. Filled circles indicate measurements above the limit of detection (LOD) and open circles indicating censored values below LOD (adjusted to LOD value). Linear regression lines with 95% confidence intervals are shown in blue. (B) Scatter plots showing mucosal IgA ELISA titres versus neutralization IC50 for all RSV-A variants following the same format as panel B.

**Supplementary Figure 8.**
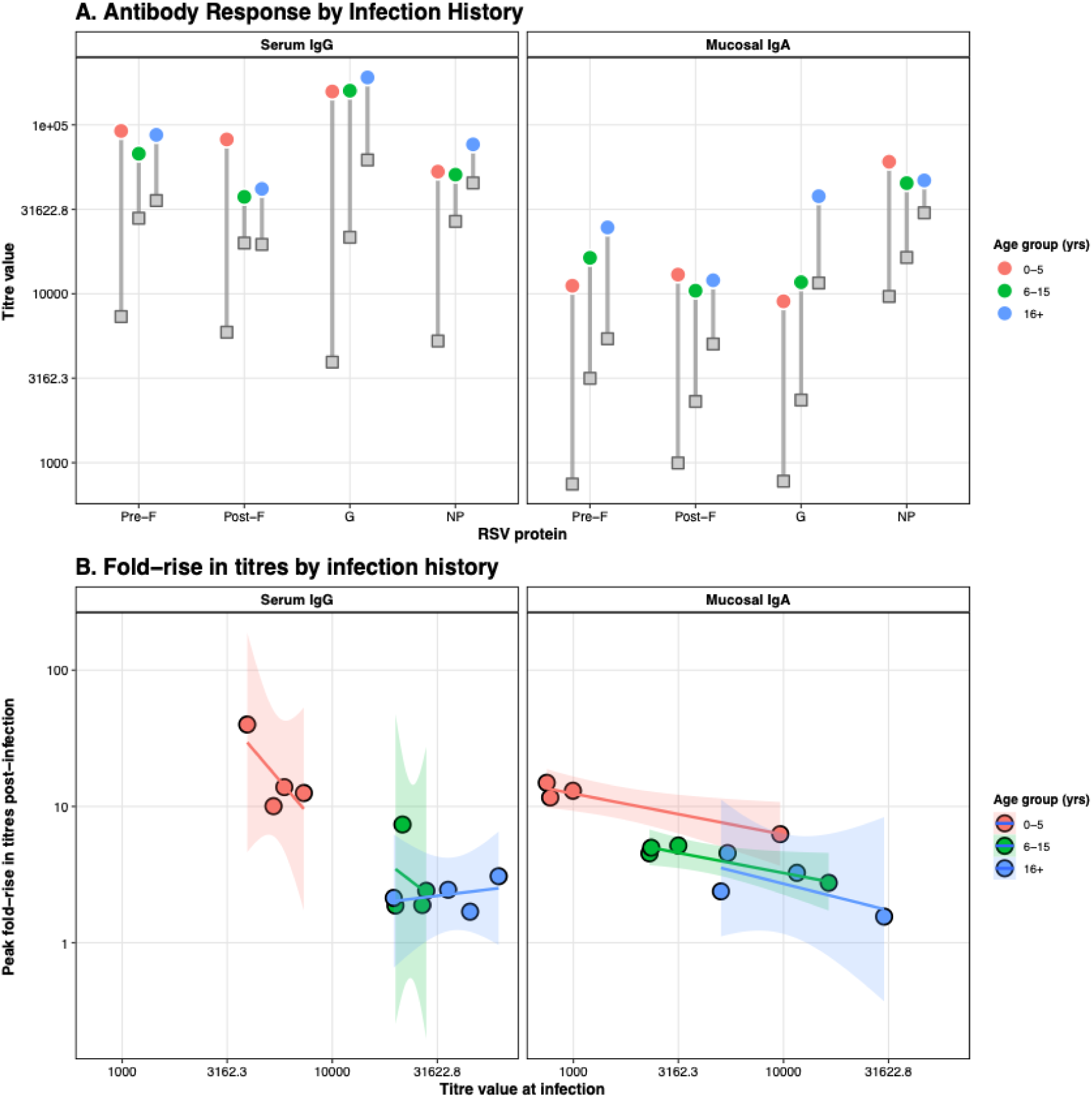
Age-dependent patterns in antibody magnitude and boosting following natural RSV infection. (A) Baseline and peak antibody titres stratified by age group for serum IgG (left panel) and mucosal IgA (right panel) against four RSV-A antigens (PreF, PostF, G, NP). Gray squares represent baseline titres at the time of infection, colored circles represent peak titres post-infection, and gray lines connecting them indicate the magnitude of antibody boosting. Point are colored according to age groups. (B) Relationship between baseline antibody titre at infection (x-axis) and fold-rise in titre post-infection (y-axis), illustrating the antibody ceiling effect where individuals with higher pre-existing titres exhibit smaller relative boosts. Linear regression lines (solid) with 95% confidence intervals (shaded regions) demonstrate the negative relationship

## Notes

### Competing Interest Statement

The authors have declared no competing interest.

### Author Declarations

Study approval was given by the joint Gambia Government and Medical Research Council Unit, The Gambia (MRCG) Ethics committee, and the London School of Hygiene and Tropical Medicine ethics committee (LEO project ID 22556). The protocol was registered at clinicaltrials.gov (NCT05952336).

## REFERENCES

1. Li, Y. et al. Global, regional, and national disease burden estimates of acute lower respiratory infections due to respiratory syncytial virus in children younger than 5 years in 2019: a systematic analysis. Lancet 399, 2047–2064 (2022).

2. Gill, C. J. et al. Infant deaths from respiratory syncytial virus in Lusaka, Zambia from the ZPRIME study: a 3-year, systematic, post-mortem surveillance project. Lancet Glob Health 10, e269–e277 (2022).

3. Papi, A. et al. Respiratory Syncytial Virus Prefusion F Protein Vaccine in Older Adults. N. Engl. J. Med. 388, 595–608 (2023).

4. Walsh, E. E. et al. Efficacy and Safety of a Bivalent RSV Prefusion F Vaccine in Older Adults. N. Engl. J. Med. 388, 1465–1477 (2023).

5. Hammitt, L. L. et al. Nirsevimab for Prevention of RSV in Healthy Late-Preterm and Term Infants. N. Engl. J. Med. 386, 837–846 (2022).

6. Kampmann, B. et al. Bivalent Prefusion F Vaccine in Pregnancy to Prevent RSV Illness in Infants. N. Engl. J. Med. 388, 1451–1464 (2023).

7. Moline, H. L. Early Estimate of Nirsevimab Effectiveness for Prevention of Respiratory Syncytial Virus–Associated Hospitalization Among Infants Entering Their First Respiratory Syncytial Virus Season — New Vaccine Surveillance Network, October 2023–February 2024. MMWR Morb Mortal Wkly Rep 73, (2024).

8. Williams, T. C. et al. Bivalent prefusion F vaccination in pregnancy and respiratory syncytial virus hospitalisation in infants in the UK: results of a multicentre, test-negative, case-control study. The Lancet Child & Adolescent Health 9, 655–662 (2025).

9. Se, F., P, T., Dc, K., R, X. & Pb, D. Effectiveness and Safety of Respiratory Syncytial Virus Vaccine for US Adults Aged 60 Years or Older. JAMA network open 8, (2025).

10. CDC. RSV Vaccine Guidance for Adults. Respiratory Syncytial Virus Infection (RSV) https://www.cdc.gov/rsv/hcp/vaccine-clinical-guidance/adults.html (2025).

11. Chaumont, A. et al. Host immune response to respiratory syncytial virus infection and its contribution to protection and susceptibility in adults: a systematic literature review. Expert Review of Clinical Immunology 21, 745–760 (2025).

12. Mazur, N. I., Caballero, M. T. & Nunes, M. C. Severe respiratory syncytial virus infection in children: burden, management, and emerging therapies. The Lancet 404, 1143–1156 (2024).

13. Tang, J. W. & Loh, T. P. Correlations between climate factors and incidence—a contributor to RSV seasonality. Reviews in Medical Virology 24, 15–34 (2014).

14. Reicherz, F. et al. Recovery of Antibody Immunity After a Resurgence of Respiratory Syncytial Virus Infections. J Infect Dis 231, e840–e845 (2025).

15. Asseri, A. A. Respiratory Syncytial Virus: A Narrative Review of Updates and Recent Advances in Epidemiology, Pathogenesis, Diagnosis, Management and Prevention. Journal of Clinical Medicine 14, (2025).

16. Vissers, M., Ahout, I. M. L., de Jonge, M. I. & Ferwerda, G. Mucosal IgG Levels Correlate Better with Respiratory Syncytial Virus Load and Inflammation than Plasma IgG Levels. Clin Vaccine Immunol 23, 243–245 (2016).

17. Öner, D. et al. Serum and mucosal antibody-mediated protection and identification of asymptomatic respiratory syncytial virus infection in community-dwelling older adults in Europe. Front Immunol 15, 1448578 (2024).

18. Maltseva, M., Keeshan, A., Cooper, C. & Langlois, M.-A. Immune imprinting: The persisting influence of the first antigenic encounter with rapidly evolving viruses. Human Vaccines & Immunotherapeutics 20, 2384192 (2024).

19. Jarju, S., et al. Incidence, Shedding and Transmission Amongst Common Respiratory Viruses: Data from a Prospective, Longitudinal, Household Cohort Study in the Gambia. SSRN Scholarly Paper at 10.2139/ssrn.5516505 (2025).

20. Hodgson, D. et al. Impact of SARS-CoV-2 exposure history on antibody kinetics and correlates of protection in The Gambia. 2026.01.02.26343369 Preprint at 10.64898/2026.01.02.26343369 (2026).

21. Jarju, S. et al. High SARS-CoV-2 incidence and asymptomatic fraction during Delta and Omicron BA.1 waves in The Gambia. Nat Commun 15, 3814 (2024).

22. Jagne, Y. J. et al. Compartmentalised mucosal and blood immunity to SARS-CoV-2 is associated with high seroprevalence before the Delta wave in Africa. Commun Med 5, 178 (2025).

23. Cohen, C. et al. Incidence and transmission of respiratory syncytial virus in urban and rural South Africa, 2017-2018. Nat Commun 15, 116 (2024).

24. Munywoki, P. K. et al. Continuous Invasion by Respiratory Viruses Observed in Rural Households During a Respiratory Syncytial Virus Seasonal Outbreak in Coastal Kenya. Clin. Infect. Dis. 67, 1559–1567 (2018).

25. White, E. B. et al. Influenza Vaccine Effectiveness Against Illness and Asymptomatic Infection in 2022–2023: A Prospective Cohort Study. Clin Infect Dis 80, 893–900 (2025).

26. Arnold, B. F., Scobie, H. M., Priest, J. W. & Lammie, P. J. Integrated Serologic Surveillance of Population Immunity and Disease Transmission. Emerg Infect Dis 24, 1188–1194 (2018).

27. Mandrekar, J. N. Receiver operating characteristic curve in diagnostic test assessment. J Thorac Oncol 5, 1315–1316 (2010).

28. Frivold, C. et al. Correlates of risk of respiratory syncytial virus disease: a prospective cohort study. Nat Commun 16, 8490 (2025).

29. McIlwain, D. R. et al. Human influenza virus challenge identifies cellular correlates of protection for oral vaccination. Cell Host Microbe 29, 1828–1837.e5 (2021).

30. Habibi, M. S. et al. Impaired Antibody-mediated Protection and Defective IgA B-Cell Memory in Experimental Infection of Adults with Respiratory Syncytial Virus. Am J Respir Crit Care Med 191, 1040–1049 (2015).

31. Öner, D. et al. Serum and mucosal antibody-mediated protection and identification of asymptomatic respiratory syncytial virus infection in community-dwelling older adults in Europe. Front. Immunol. 15, (2024).

32. Bagga, B. et al. Effect of Preexisting Serum and Mucosal Antibody on Experimental Respiratory Syncytial Virus (RSV) Challenge and Infection of Adults. J Infect Dis 212, 1719–1725 (2015).

33. Lim, W. W., Leung, N. H. L., Sullivan, S. G., Tchetgen Tchetgen, E. J. & Cowling, B. J. Distinguishing Causation from Correlation in the Use of Correlates of Protection to Evaluate and Develop Influenza Vaccines. Am J Epidemiol 189, 185–192 (2020).

34. Yan, H., Lamm, M. E., Björling, E. & Huang, Y. T. Multiple Functions of Immunoglobulin A in Mucosal Defense against Viruses: an In Vitro Measles Virus Model. J Virol 76, 10972–10979 (2002).

35. Corthésy, B. Multi-Faceted Functions of Secretory IgA at Mucosal Surfaces. Front Immunol 4, 185 (2013).

36. Stensballe, L. G. et al. Duration of secretory IgM and IgA antibodies to respiratory syncytial virus in a community study in Guinea-Bissau. Acta Paediatr 89, 421–426 (2000).

37. Long-lived plasma cells are generated in mucosal immune responses and contribute to the bone marrow plasma cell pool in mice | Mucosal Immunology. https://www.nature.com/articles/mi201538.

38. Radbruch, A. et al. Competence and competition: the challenge of becoming a long-lived plasma cell. Nat Rev Immunol 6, 741–750 (2006).

39. F, W. & M, S. Memory B Cells of Mice and Humans. Annual review of immunology 35, (2017).

40. Gd, V. & Mc, N. Germinal Centers. Annual review of immunology 40, (2022).

41. Angeletti, D. et al. Defining B cell immunodominance to viruses. Nat Immunol 18, 456–463 (2017).

42. Linderman, S. L. et al. Potential antigenic explanation for atypical H1N1 infections among middle-aged adults during the 2013–2014 influenza season. Proceedings of the National Academy of Sciences 111, 15798–15803 (2014).

43. Simon, A. K., Hollander, G. A. & McMichael, A. Evolution of the immune system in humans from infancy to old age. Proceedings of the Royal Society B: Biological Sciences 282, 20143085 (2015).

44. Maltseva, M., Keeshan, A., Cooper, C. & Langlois, M.-A. Immune imprinting: The persisting influence of the first antigenic encounter with rapidly evolving viruses. Human Vaccines & Immunotherapeutics 20, 2384192 (2024).

45. Reis, J. & Shaman, J. Simulation of four respiratory viruses and inference of epidemiological parameters. Infectious Disease Modelling 3, 23–34 (2018).

46. Jang, E., Lee, S., Han, J. & Chang, J. Single-dose intranasal adenovirus-based RSV vaccines targeting G and M2. npj Vaccines 10, 139 (2025).

47. Lei, H., et al. Intranasal Inoculation of Cationic Crosslinked Carbon Dots-Adjuvanted Respiratory Syncytial Virus F Subunit Vaccine Elicits Mucosal and Systemic Humoral and Cellular Immunity. MedComm (2020) 6, e70146 (2025).

48. Walsh, E. E., Peasley, M., Branche, A. R. & Falsey, A. R. Respiratory Syncytial Virus Humoral Antibody Responses in Older Adults After Vaccination or Infection. J Infect Dis 231, e1146–e1150 (2025).

49. Maier, C. et al. Mucosal immunization with an adenoviral vector vaccine confers superior protection against RSV compared to natural immunity. Front Immunol 13, 920256 (2022).

50. Sinha, D., Gopalakrishna, P. K., Paul, S. & Longet, S. A user’s guide to designing efficient and safe mucosal vaccines: Challenges & potentials. Oxf Open Immunol 6, iqaf007 (2025).

51. Neutra, M. R. & Kozlowski, P. A. Mucosal vaccines: the promise and the challenge. Nat Rev Immunol 6, 148–158 (2006).

52. Zhang, X. et al. Progress and challenges in the clinical evaluation of immune responses to respiratory mucosal vaccines. Expert Rev Vaccines 23, 362–370 (2024).

53. Transmission of Respiratory Viruses in Households in The Gambia: a Longitudinal Cohort Study (TransVIR) (TransVIR). ClinicalTrials.gov https://clinicaltrials.gov/study/NCT05952336 (2023).

54. Broadbent, L. et al. An endogenously activated antiviral state restricts SARS-CoV-2 infection in differentiated primary airway epithelial cells. PLOS ONE 17, e0266412 (2022).

55. Jorquera, P. A. & Tripp, R. A. Quantification of RSV Infectious Particles by Plaque Assay and Immunostaining Assay. in Human Respiratory Syncytial Virus: Methods and Protocols (eds Tripp, R. A. & Jorquera, P. A.) 33–40 (Springer, New York, NY, 2016). doi:10.1007/978-1-4939-3687-8_3.

56. Three-dose vaccination elicits neutralising antibodies against omicron - The Lancet. https://www.thelancet.com/journals/lancet/article/PIIS0140-6736(22)00092-7/fulltext.

57. Teunis, P. F. M., van Eijkeren, J. C. H., de Graaf, W. F., Marinović, A. B. & Kretzschmar, M. E. E. Linking the seroresponse to infection to within-host heterogeneity in antibody production. Epidemics 16, 33–39 (2016).

