## Supplementary results for "Mucosal IgA to pre-fusion F protein predicts protection from RSV infection in a high burden setting"

Overview

This document presents a Bayesian hierarchical model that jointly estimates antibody kinetics and a titre-based correlate of protection (CoP) for immunological biomarkers measured in a longitudinal surveillance cohort. The model captures the dynamic interplay between individual-level antibody waning prior to infection, post-infection boosting responses across multiple biomarkers, and exposure intensity within households, to link pre-exposure antibody titres to the probability of RSV infection.

1. OUTLINE OF THE DATA

Our data consists of individual-level longitudinal serological samples taken from a cohort study in The Gambia described previously.^1–4^

**Data cleaning and serological sample exclusion process**

The initial dataset comprised 956 serum IgG samples and 958 mucosal IgA samples tested against 16 biomarkers. From the IgG dataset, we excluded five duplicate samples and four samples with missing titre data. From the IgA dataset, we removed four samples with insufficient volume, one sample that could not be linked to participant metadata, and three samples lacking metadata and IgA testing records. After merging datasets and retaining only samples with both IgG and IgA measurements, and removing one participant with discordant bleed dates (6 samples) and one with no mucosal IgA data at recruitment (3 samples), the cleaned dataset comprised 926 serological samples from 342 individuals in 52 households (**Supplementary Methods Figure 1**).

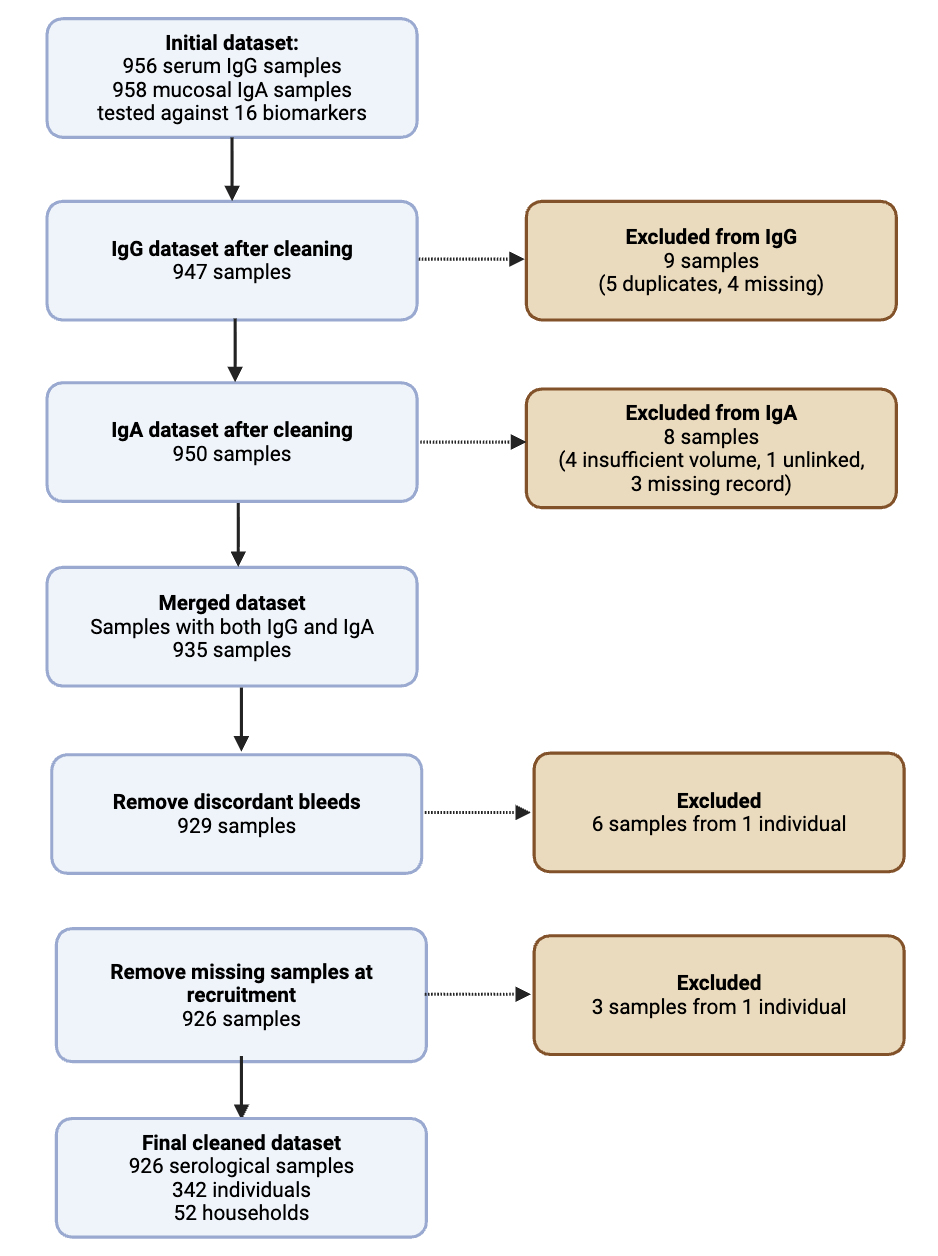

**Supplementary Methods Figure 1.** Data cleaning and serological sample exclusion process.

2. GOALS OF THE ANALYSIS

We quantify how RSV antibody titres evolve over time and how they relate to infection risk, by combining PCR outcomes with longitudinal serology from a household cohort study in The Gambia. A key challenge in this setting is that PCR surveillance does not capture all infections, as the window of PCR detectability may not align with the sampling schedule. We therefore (i) use multi-biomarker serological rises to identify infections missed by weekly PCR, and (ii) fit a joint Bayesian model that links individual kinetics (waning and post-infection boosting), to household exposure intensity, and a titre-based correlate of protection (CoP). This framework enables us to estimate infection timing, protection curves, and group-level differences with principled uncertainty.

**Major objectives:**

1. **Detect missed infections in the PCR-negative cohort.**
   Develop and calibrate a multi-biomarker rank-aggregation rule that flags likely infections using upper-tail fold-rises in biomarkers; quantify its operating characteristics as a function of the assumed PCR detection rate $\rho$.
2. **Estimate antibody kinetics and a titre-based CoP in a unified model.**
   Fit a Bayesian hierarchical model that captures individual pre-infection waning and post-infection boosting, links titres to infection risk via a logistic protection curve (given exposure), and marginalizes over exposure using household infection counts and a small background risk.
3. **Characterize heterogeneity and identify informative biomarkers.**
   Compare biomarkers and estimate hierarchical effects of age) on kinetics and protection, yielding interpretable summaries (e.g., titre levels associated with specified protection) and guiding selection of biomarkers that best predict infection risk.
3. DESCRIPTION OF THE MODEL

3.1 OVERVIEW OF THE MATHEMATICAL STRUCTURE

We developed a Bayesian hierarchical model to describe longitudinal antibody titres in individuals with known or inferred infection and vaccination events. The model captures individual-level waning, functional boosting responses to exposures (infection or vaccination), and uses these dynamics to estimate a biomarker-based correlate of protection (CoP) via a logistic function.

3.1. Inferring missed RSV infections in the PCR-negative cohort using serology

3.1.1. Overview

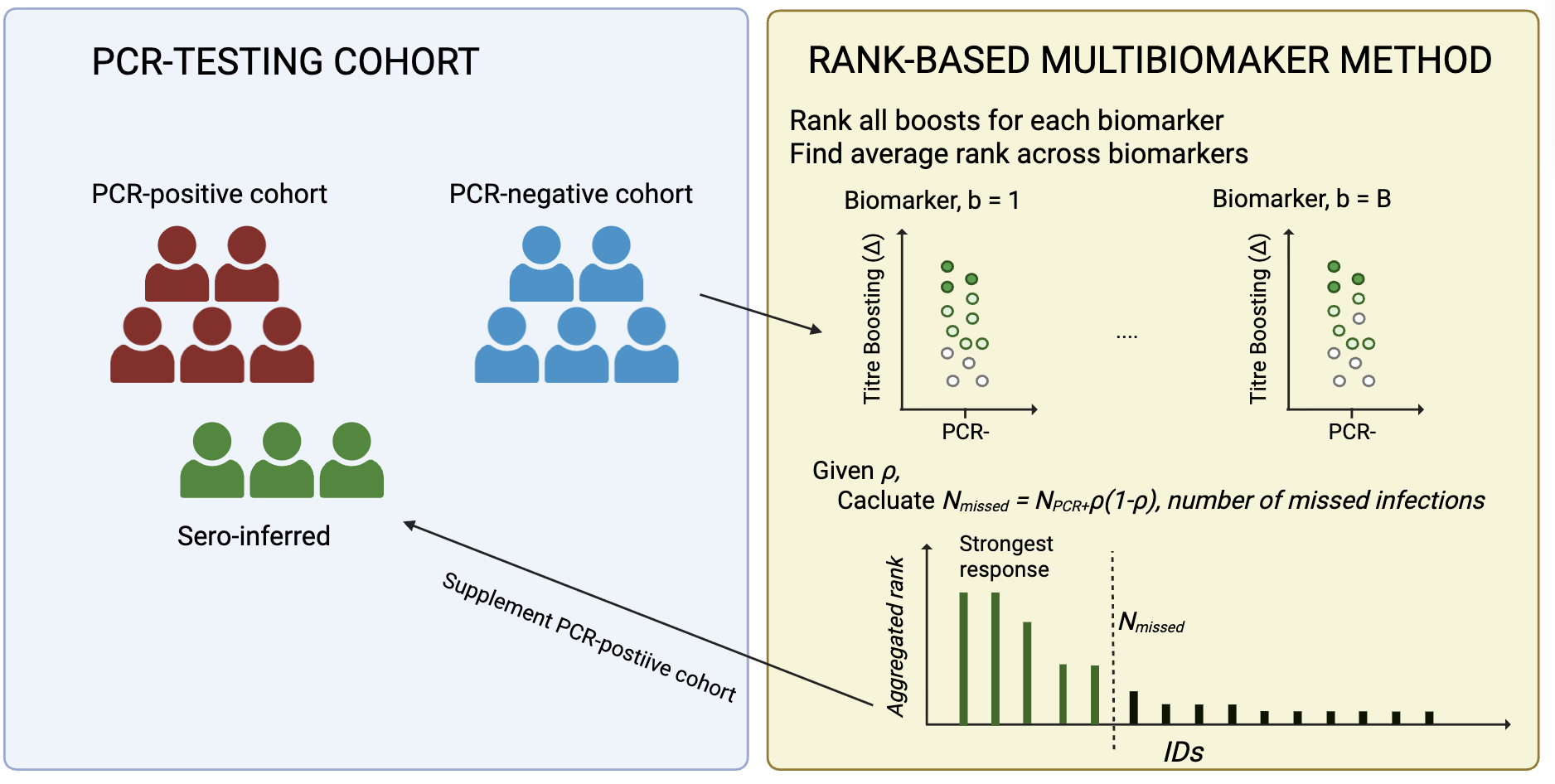

**Supplementary Methods Figure 2**. Schematic showing the methods for which serologically missed infection from the PCR- group are identified.

A schematic of the ensemble classification rule; used to identify the RSV infections in the PCR-negative cohort is given in **Supplementary Methods Figure 2**. We developed a rank-based multi-biomarker detection method to identify RSV infections issued by PCR testing. This method ranks PCR-negative individuals by their aggregated serological response across all biomarkers and selects those with the strongest responses as missed infections. The selection is determine by $\rho$, the estimated PCR detection rate (proportion of all true infections detected by PCR), which translates to the expected number of missed infections $N_{missed}= N_{PCR+}\rho/(1-\rho)$. Because serologically detected cases supplement an existing cohort of PCR-positive infections, the method is designed to identify individuals with the most consistent and substantial antibody responses.

3.1.2. Mathematical framework

Define individuals, $i \in\left\{ 1, 2, \ldots, N \right\}$, and biomarkers, $b \in\left\{ 1, 2, \ldots, B=16 \right\}$, and log_10_ titre values for individual *i* and biomarker *b* at time points 1 and 2 as $T_{i,1}^{b}$ and $T_{i,2}^{b}$ respectively. Finally, let $Y_{i}=\left\{ 0, 1 \right\}$, where 1 denotes PCR-positive in the study and 0 denotes PCR-negative (or “unlabeled”).

First, for each individual we determined the change in titre for each biomarker between time points 1 and 2,

$\Delta_{i}^{b}=T_{i,2}^{b}-T_{i,1}^{b}$,

and denoted the “PCR-positive” individuals as $I = \left\{ i : Y_{i}=1 \right\}$, and the “unlabeled” individuals as $U = \left\{ i : Y_{i}=0 \right\}$.

3.1.2.1 Aggregated serological response

For each individual, *b,* rank all PCR-negative individual by their titre change $\Delta_{i}^{b}$:

$$R_{i}^{b}=\mathrm{rank}\left( -\Delta_{i}^{b} \mathrm{among} \left\{ i: Y_{i}=0 \right\} \right), \mathrm{where} \mathrm{rank}\left( 1 \right)=\mathrm{highest}\Delta_{i}^{b}$$

Individuals with bigger boosts receive lower (better) rank numbers, e.g. $R_{i}^{b}=1$is the individual in $U$ with the highest boost to biomarker *b*.

3.1.2.2 Calculate average rank across biomarkers

For each individual, *i*, compute the average rank across all available biomarkers:

$$\bar{R_{i}}=\frac{1}{M}\sum_{b=1}^{B} R_{i}^{b}$$

This aggregation identifies individuals who consistently rank highly across multiple biomarkers, treats all biomarkers equally, and provides robustness against extreme responses.

3.1.2.3 Calculate average rank across biomarkers

Flag the top $N_{flagged}$ , individuals with the lowest $\bar{R_{i}}$, as missed infections:

$$I_{i}^{sero}=\left\{ \begin{aligned} 1 \mathrm{if}Y_{i}=0 \mathrm{and}\bar{R_{i}}\leq\bar{R}^{N_{flagged}} \\ 0 \mathrm{otherwise} \end{aligned} \right.$$

3.1.2.4 Determining the detection rate of PCR surveillance, $\rho$

The PCR detection rate $\rho$ reflects the proportion of all RSV-negative which were missed by weekly PCR surveillance. This could be influenced by the viral shedding in RSV relative to the sampling schedule and PCR sensitivity. We considered PCR detection rates of $\rho=\{1.0, 0.9, 0.8, 0.7\}$ the base case is $0.8$ in the manuscript. This value is chosen to reflect a mean PCR-detectable RSV shedding of 5-6 days. This was informed by community PCR household studies in sub-Saharan Africa reporting all-age mean shedding of 6.5–11.3 days^5,6^, adjusted downward based on consistent evidence that shedding is shorter in older age groups and asymptomatic infections^6,7^. Assuming testing occurred at ~7-day intervals, we estimate $\rho$ = (5.5)/7 ~ 0.8.

3.1.3 Algorithm implementation

The algorithm to detect missed infectoions as follows:

1. **Data preparation:**

- Extract measurements at timepoints 1 and 2 for all individuals and biomarkers
- Compute log10 fold-changes: $\Delta_{i}^{b}=T_{i,2}^{b}-T_{i,1}^{b}$,
- Identify PCR-positive individuals as $I = \left\{ i : Y_{i}=1 \right\}$, and the “unlabeled” individuals as $U = \left\{ i : Y_{i}=0 \right\}$

2. **Biomarker informativeness:**

- For each individual *i*:
  - Within each biomarker *b*, rank by $\Delta_{i}^{b}: \mathrm{rank}\left( -\Delta_{i}^{b} \mathrm{among} \left\{ i: Y_{i}=0 \right\} \right)$
  - Calculate average rank across biomarkers:

$$\bar{R_{i}}=\frac{1}{M}\sum_{b=1}^{B} R_{i}^{b}$$

3. **Rank and select for a** $\boldsymbol{\rho}$

- Among PCR-negative individuals
  - Sort by ascending $\bar{R_{i}}$ (lowest average rank = best)
  - Calculate $N_{flagged}=\rho{/(1- \rho)N}_{PCR+}$
  - Flag top $N_{flagged}$ individuals (those with lowest $R_{i}$) as missed infections

4. **Update infection status:**

- Combine PCR-confirmed and serologically-detected infections:
  - $\bar{I_{i}}=max(I_{i}, I_{i}^{sero})$

3.1.4. Graphical overview of ranks for different $\rho$

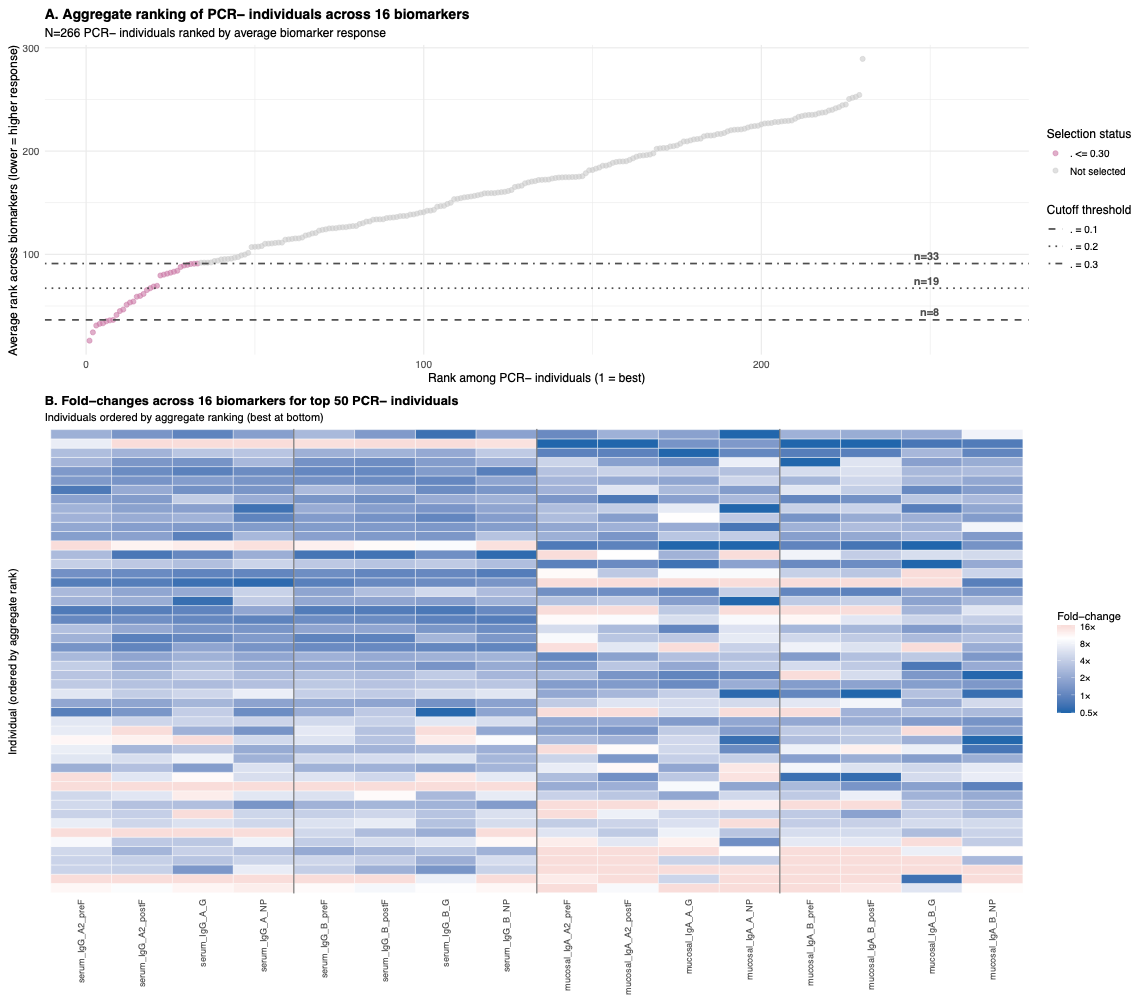

**Supplementary Methods Figure 3. Serological detection strategy across different ρ thresholds. (A)** shows the aggregate ranking of all PCR-negative individuals (N=266) based on their antibody responses across 16 biomarkers (8 RSV-A and 8 RSV-B proteins measured for both serum IgG and mucosal IgA). Horizontal lines indicate cutoff thresholds for different values of ρ (the proportion of PCR-confirmed infections missed by PCR testing): ρ=0.9 (n=8 individuals selected, dashed line), ρ=0.8 (n=19, dotted line), and ρ=0.7 (n=33, dot-dash line). **(B)** A heatmap of fold-changes across all 16 biomarkers for the top 50 PCR-negative individuals ranked by aggregate score. Each row represents one individual (ordered from best rank at bottom to 50th best at top), and each column represents a biomarker. Fold-changes are calculated as 10^(titre_2 - titre_1) and displayed on a log10 scale, with red indicating increases, blue indicating decreases, and white indicating no change.

3.2. Bayesian model to infer infection kinetics and COP

3.2.1. Overview

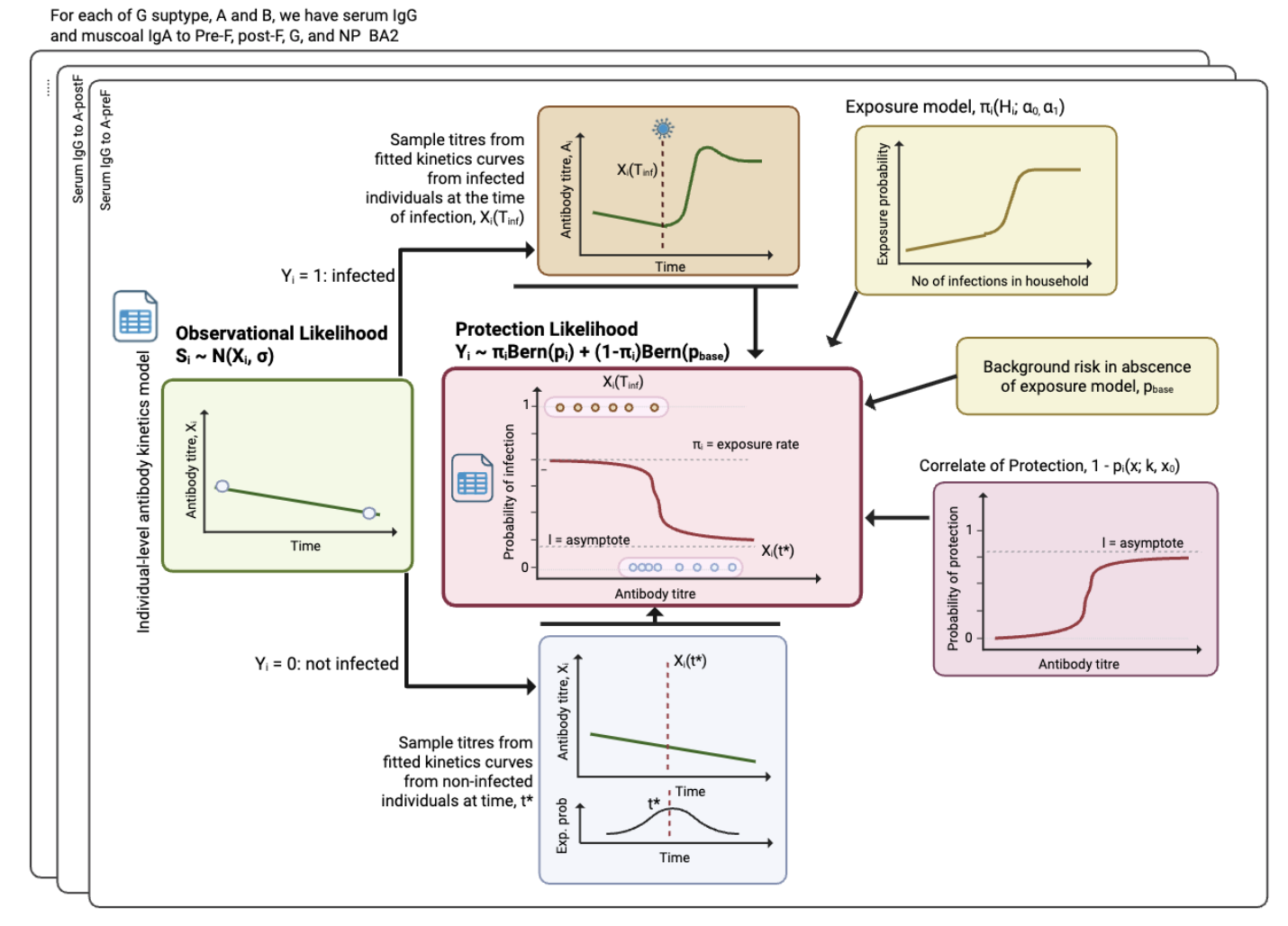

**Supplementary Methods Figure 4**. Full schematic of the Bayesian model.

We developed a Bayesian hierarchical model to jointly estimate antibody kinetics following RSV infection and identify correlates of protection against infection. The model accounts for

- Longitudinal antibody titre measurements at up to four time points per individual
- Left-censored observations below the limit of detection (LOD)
- Individual-level variation in antibody waning rates and infection-induced boosting
- Uncertainty in exposure status
- Logistic protection curves relative pre-exposure antibody titre to infection risk

3.2.2. Setup and notation

Define individuals $i \in\left\{ 1, 2, \ldots, N \right\}$, and choose a biomarker b (note we fit each biomarker separately, so we drop the *b*). Let log_10_ titre values at timepoint $j =\{1,2 , 3, 4\}$ denoted by, $S_{i, j},$ occuring at times $T_{i, j}$days and allow time to run between $T_{i, 1}\leq t_{i} \leq T_{i, 4}$. We have an infection indicator $Y_{i} = \left\{ 0, 1 \right\}$, which denotes infection if 1, no infection otherwise, and in the case of infection, the time is given by $T_{inf}$. The lower limit of detection is defined as a value, LOD.

3.2.3. Likelihood function for the antibody kinetics model

*3.2.3.1 Antibody kinetics functions*

The log_10_ titre trajectory value, $X_{i}\left( t \right)$, at time, *t,* for each individual, *i*, is modelled using a biphasic kinetics models adapted from Teunis et al. (2016)^8^. This model consists of:

1. **Pre-infection wane/continuous wane:** Over the course of the study we assume that titre values wane linearly until the end of the study (if there is no infection), or until infection occurs. The equation the give the log_10_ titre trajectory value is:

$X_{i}\left( t \right) = \max\left( 0, S_{i, 1} - \omega\cdot\left( t -T_{i, 1} \right) \right)$,

Where the rate of waning is defined by the parameter $\omega$, and we truncate the waning at a log_10_ value of 0.

2. **Post-infection boost:** Each infection leads to a functional antibody increase modelled by a power-function formulation of Teunis et al. (2016).^8^ Specifically, the boost (change relative to the log_10_ titre trajectory value at infection, $X_{i}\left( T_{inf} \right)$ on the log_10_) following the time after infection is give by the equation:

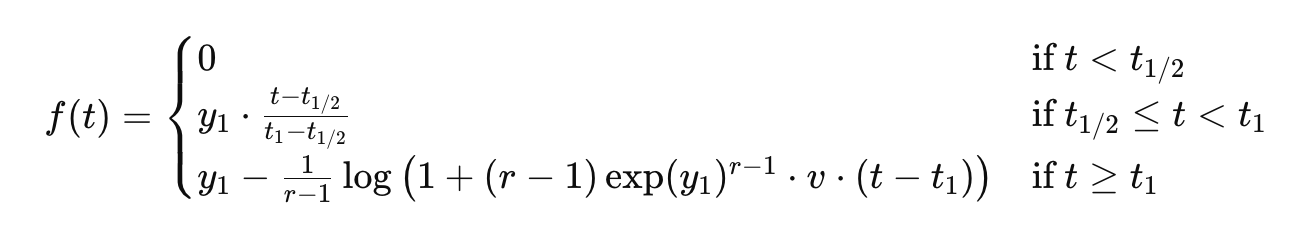
where $y_{1}$ controls the peak amplitude of the boost, $t_{1}$ is the time of the peak, *r* controls the curvature, and v is a small constant fixed to 0.001 to stabilize waning. Thus, the value of the antibody kinetics time $t_{inf}$ after infection is given by

$$X_{i}\left( t \right) = X_{i}\left( T_{inf} \right) + f\left( t_{inf} \right)$$

From the log_10_ titre trajectory values, we can estimate the model-predicted titre values at any time *t*, for all individuals *i.*

*3.2.3.2. Observational likelihood*

Using these model-predicted titre values, we determine the titre value times after *j = 1*, $X_{i}\left( T_{i, j} \right)$ and define an observational model likelihood (not censored)

$$S_{i, j}\sim N\left( X_{i}\left( T_{i, j} \right), \sigma\right),$$

With PDF *P*. If the titre is left-censored (below the LOD):

$$P\left( S_{i, j}<LOD | X_{i}\left( T_{i, j} \right), \sigma\right)= \Phi\left( \frac{LOD-X_{i}\left( T_{i, j} \right)}{\sigma} \right)$$

Where $\sigma$, is a standard deviation of the normal distributions, and $\Phi(\cdot)$ is the standard normal CDF. We represent the likelihoods for individual, *i* at timepoint *j* $L_{i}^{j}\left( D|\theta\right)$. Thus, the total likelihood across all individuals and timepoints is:

$$\mathcal{L}^{1}\left( D | \theta\right)= \prod_{i=1}^{N} \prod_{j=1}^{4} L_{i}^{j}\left( D|\theta\right)=\prod_{i=1}^{N} \prod_{j=1}^{4} \left[ 1\left( \delta_{i,t}=0 \right)\cdot P(S_{i, j}|X_{i}\left( T_{i, j} \right), \sigma)+1\left( \delta_{i,t}=1 \right)\Phi\left( \frac{LOD-X_{i}\left( T_{i, j} \right)}{\sigma} \right) \right]$$

Where $1\left( \delta_{i,t} \right)$ is an indicator function for if a sample is below the limit of detection.

3.2.4. Likelihood function for the Protection model

We model infection risk as function of pre-exposure antibody titre using a logistic regression framework with mixture components.

*3.2.4.1 Defining the Protection model*

Defining t* as the calendar time when 50% of all cases have occurred. The titre values used as inputs in the exposure-conditional protection model (see below) is given by:

$$x_{i} =\left\{ \begin{aligned} X_{i}\left( T_{inf} \right), Y_{i} = 1 \\ X_{i}\left( t^{*} \right), Y_{i} = 0 \end{aligned} \right.$$

That is, if an individual is infected their titre value at infection is determine from the antibody kinetics function, and if an individual is not infected it is the estimated a mid-season control titre. Using these inputs, we define the equation which defines the probability of protection against infection given exposure.

$$p_{i}^{exp} =\Pr\left( Y_{i}=1|exposed, x_{i} \right)\mathcal{= l +}\left( 1 - \mathcal{l} \right)\frac{1}{1 + \exp\left( \beta\left( x_{i}-x_{0} \right) \right)}, \beta>0$$

It is a decreasing logistic curve with a floor value, $\mathcal{l}$(or lower bound asymptote), to allow residual risk at high titre values: Here, $\beta$, is the gradient of the slope at the midpoint, x_0_.

*3.2.4.2 Defining the Exposure model*

Let $H_{i}$ be the number of infected household members for an individual, *i*, over the season and use $H_{i}$ as proxy for the intensity of exposure for each individual. The relationship between $H_{i}$ and the individual probability of infection is assumed to be logistic:

$$\pi_{i} =\Pr\left( exposed|H_{i} \right) = \frac{1}{1 + \exp\left( \alpha_{1}\left( \alpha_{0}-H_{i} \right) \right)}$$

With $\alpha_{0}$ is the exposure level at which the gradient is steepest at $\alpha_{1}$> 0.

*3.2.4.3 Infection outcome (mixture) likelihood*

Reminding ourselves of the conditional infection probability above, we also define:

$p_{base =}\Pr\left( Y_{i}=1|not exposed \right)$,

The baseline probability infection unexposed indviiduals. Which is defined to include the possibility of infections occurring outside of the exposure model defined in this framework (e.g. infections from outside influence). It provides numerical stability during the inference process and is keep close to 0 (see priors).

With these defined, we can marganlise out the latent exposure state, $\pi_{i}$ in the likelihood through the equations:

$\Pr\left( Y_{i}=1|\left( x_{i}, H_{i} \right) = \pi_{i} p_{i}^{exp} + \left( 1-\pi_{i} \right)p_{base} \right)$,

$$\Pr\left( Y_{i}=0|\left( x_{i}, H_{i} \right)= \pi_{i}\left( 1- p_{i}^{exp} \right) + \left( 1-\pi_{i} \right)\left( 1-p_{base} \right) \right)$$

Thus the likelihood contribution is given by the equation

$$L_{i}^{mixture}\left( \theta|D \right) = \log\left( \pi_{i}\cdot Bern\left( Y_{i}|p_{i}^{exp} \right) +\left( 1-\pi_{i} \right)\cdot Bern\left( Y_{i}|p_{base} \right) \right),$$

And overall:

$$\mathcal{L}^{2}\left( D | \theta\right)=\prod_{i=1}^{N} L_{i}^{mixture}\left( D|\theta\right)$$

3.2.5 Full posterior and priors

The full likelihood is a combination of the likelihoods:

$${\mathcal{L}\left( D | \theta\right)\mathcal{=L}}^{1}\left( D | \theta\right)\mathcal{L}^{2}\left( D | \theta\right)$$

The prior distributions for the parameters are defined below:

3.2.5.1 Antibody kinetics model; priors and support

*Antibody Kinetics*

$$\omega_{0} \sim N\left( 0.001, 0.0001 \right)\left[ -0.1, 0.1 \right]$$

$$y_{0} \sim N\left( 0, 2 \right) \left[ -6, 6 \right]$$

$$t_{1} \sim N\left( 14, 4 \right) \left[ 7, 40 \right]$$

$$r_{0} \sim U\left( 1, 5 \right) \left[ 1, 5 \right]$$

*Likelihood*

$$\sigma\sim Exponential\left( 1 \right)$$

3.2.5.2 Protection model; priors and support

*Correlate of Protection model*

$$\mathcal{l\sim}U\left( 0, 0.2 \right) \left[ 0, 0.2 \right]$$

$$\beta_{0}\sim N\left( -1, 1 \right) \left[ -4, 4 \right]$$

$x_{0,0}\sim\frac{N\left( \left[ \max\left( S \right) + \min\left( S \right) \right] \right]}{2}, 1 \left[ \min\left( S \right), \max\left( S \right) \right]$, where S is all the titre values in the study.

*Exposure model*

$$\alpha_{1}\sim N\left( 1, 0.5 \right) \left[ 0, \right]$$

$$\alpha_{0}\sim N\left( 2, 2 \right) \left[ -5, 10 \right]$$

*Likelihood*

$$p_{base} \sim Beta\left( 1, 50 \right)$$

3.2.6. HIERARCHICAL EFFECTS

We want to assess the influence of vaccine type and host factors (age and time since vaccination) on the kinetics of antibody production. To do this, we add hierarchical effects to our parameters of interest:

Antibody kinetics

Formula for hierarchical effects,

$\omega= \omega_{0} + z_{\omega}\sigma_{\omega}$, $\sigma_{\omega} \sim N\left( 0, 0.005 \right) \left[ 0, \right]$

$y_{1} = y_{0} + z_{y}\sigma_{y}$, $\sigma_{y} \sim N\left( 0, 1 \right) \left[ 0, \right]$

$r = r_{0} + z_{r}\sigma_{r}$, $\sigma_{r} \sim N\left( 0, 1 \right) \left[ 0, \right]$

Protection model

$x_{0} = x_{0,0} + z_{x}\sigma_{x}$, $\sigma_{x} \sim N\left( 0, 1 \right) \left[ 0, \right]$

$\beta= \beta_{0} + z_{\beta}\sigma_{\beta}$, $\sigma_{\beta} \sim N\left( 0, 0.3 \right) \left[ 0, \right]$

For all the above equations, z ~ *N*(0, 1).

3.2.7. Dual biomarker extension

The model extends naturally to multiple biomarkers (e.g. IgG and IgA) by:

For biomarker $b\in\{1, 2\}$:

$${S_{i, j}}^{(b)}\sim N\left( {X_{i}}^{(b)}\left( {T_{i, j}}^{(b)} \right),\sigma^{(b)} \right),$$

Where each biomarkers has

- Biomarker specific waning rate: $\omega^{(b)}$
- Biomarker specific boost parameters: $y^{(b)}, t^{(b)}, r^{(b)}$
- Biomarker-specific measurment error $\sigma^{(b)}$
- Biomarker specific LOD: LOD^(b)^

And all processes for both biomarkers are fitted simultaneously.

Further, the correlation of protection model can be extended to incude both biomarkers:

1 biomarker: $\beta\left( x_{i}-x_{0} \right)$

2 biomarkers: $\beta^{1}\left( {x_{i}}^{1}-{x_{0}}^{1} \right)+\beta^{2}\left( {x_{i}}^{2}-{x_{0}}^{2} \right)+\beta^{1,2}\left( {x_{i}}^{1}-{x_{0}}^{1} \right)\left( {x_{i}}^{2}-{x_{0}}^{2} \right)$,

This alllows assessment of whether each biomarker independent predicts protection and whether biomarkers provide complementary information.

4. IMPLEMENTATION

4.1 Model implementation

All Bayesian hierarchical models were implemented in Stan (version 2.35.0) and fitted using Hamiltonian Monte Carlo (HMC) via the cmdstanr R package (version 0.5.3) in R version 4.2.3. The posterior package (version 1.4.1) was used for posterior inference and diagnostics, and the bayesplot package (version 1.10.0) for visualisation of MCMC diagnostics.

We fitted two classes of Bayesian hierarchical models:

Single biomarker models (16 models total): Each model estimated antibody kinetics (post-infection boosting and pre-infection waning) and correlates of protection for a single biomarker-protein combination. The 16 models comprised:

- 2 biomarkers (serum IgG, mucosal IgA) ×

- 8 viral proteins (RSV-A: PreF, PostF, G, NP; RSV-B: PreF, PostF, G, NP)

Dual biomarker models (6 models total): Each model simultaneously estimated kinetics and correlates of protection for both serum IgG and mucosal IgA targeting the same viral protei. The 6 models comprised:

- Serum IgG + Mucosal IgA combinations for 6 proteins (RSV-A: PreF, PostF, G; RSV-B: PreF, PostF, G)

The full code to reproduce this analysis is given at <https://github.com/ccgh-idd/cop-transvir-rsv>.

4.2. MCMC sampling procedure

For all models, we employed four independent Markov chains run in parallel, with 1,000 warmup iterations and 1,000 sampling iterations per chain (2,000 total iterations per chain), yielding 4,000 posterior draws. The target acceptance rate (adapt_delta) was set to 0.95 to minimize divergent transitions.

4.3. Convergence diagnostics

We assessed MCMC convergence using multiple diagnostic criteria for all fitted parameters:

1. **Rhat (Gelman-Rubin diagnostic):** All parameters should achieve an Rhat < 1.011, indicating excellent convergence across chains.
2. **Divergent transitions**: The proportion of divergent transitions should. Be close to 0 as possible indicating the sampler successfully explored the posterior without encountering problematic geometry.

REFERENCES

1.	Hodgson, D. *et al.* Impact of SARS-CoV-2 exposure history on antibody kinetics and correlates of protection in The Gambia. 2026.01.02.26343369 Preprint at https://doi.org/10.64898/2026.01.02.26343369 (2026).

2.	Jarju, S. *et al.* High SARS-CoV-2 incidence and asymptomatic fraction during Delta and Omicron BA.1 waves in The Gambia. *Nat Commun* **15**, 3814 (2024).

3.	Jarju, S. *et al.* Incidence, Shedding and Transmission Amongst Common Respiratory Viruses: Data from a Prospective, Longitudinal, Household Cohort Study in the Gambia. SSRN Scholarly Paper at https://doi.org/10.2139/ssrn.5516505 (2025).

4.	Jagne, Y. J. *et al.* Compartmentalised mucosal and blood immunity to SARS-CoV-2 is associated with high seroprevalence before the Delta wave in Africa. *Commun Med* **5**, 178 (2025).

5.	Cohen, C. *et al.* Incidence and transmission of respiratory syncytial virus in urban and rural South Africa, 2017-2018. *Nat Commun* **15**, 116 (2024).

6.	Munywoki, P. K. *et al.* Influence of age, severity of infection, and co-infection on the duration of respiratory syncytial virus (RSV) shedding. *Epidemiol. Infect.* **143**, 804–812 (2015).

7.	Otomaru, H. *et al.* Risk of Transmission and Viral Shedding From the Time of Infection for Respiratory Syncytial Virus in Households. *Am J Epidemiol* **190**, 2536–2543 (2021).

8.	Teunis, P. F. M., van Eijkeren, J. C. H., de Graaf, W. F., Marinović, A. B. & Kretzschmar, M. E. E. Linking the seroresponse to infection to within-host heterogeneity in antibody production. *Epidemics* **16**, 33–39 (2016).
