## Supplementary methods for "Mucosal IgA to pre-fusion F protein predicts protection from RSV infection in a high burden setting"

#### Overview

This document presents supplementary results to the main manuscript, particularly of the model fits to the four different assumptions on serologically missed infections.

### 1. PCR-ONLY MODEL $\rho = 100\%$

#### A. CONVERGENCE AND MODEL FITS

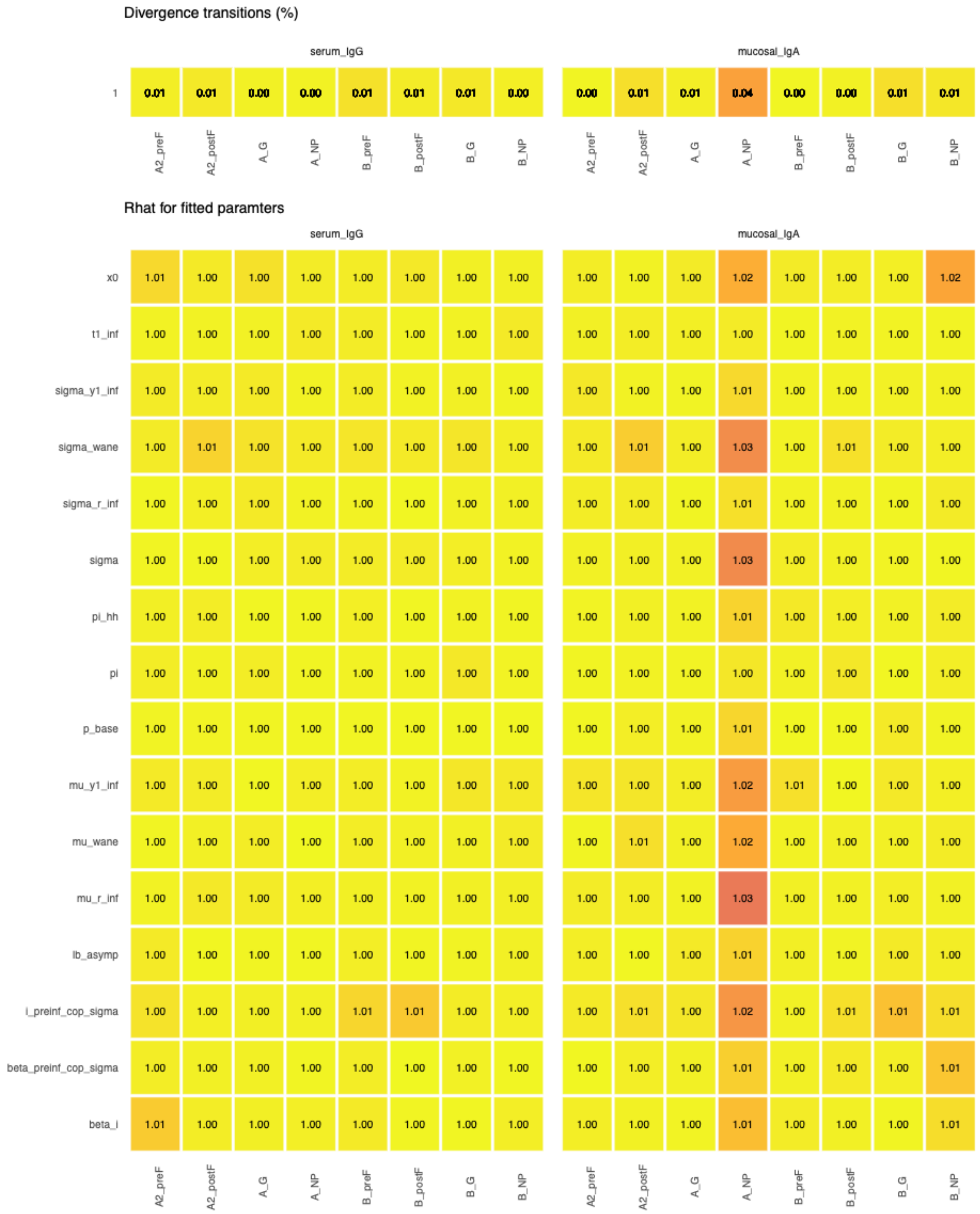


**Supplementary Results Figure 1.1. MCMC convergence diagnostics for 16 single biomarker antibody kinetics and correlates of protection models. PCR-only.** Convergence diagnostics for Bayesian hierarchical models estimating antibody kinetics and correlates of protection. The top panel shows the percentage of divergent transitions

(out of 4,000 total iterations across 4 chains) for each biomarker-protein combination. All models achieved divergent transition rates < 2%, indicating successful exploration of the posterior distribution. Parameters shown include: mu_wane and sigma_wane (waning kinetics), mu_y1_inf and sigma_y1_inf (peak fold-rise), mu_r_inf and sigma_r_inf (boosting rate), t1_inf (time to peak), sigma (measurement error), lb_asymp (asymptotic lower bound), x0 (protection threshold), i_preinf_cop_sigma (pre-infection titre variability effect), beta_i (infection effect on susceptibility), beta_preinf_cop_sigma (pre-infection titre effect on protection), pi (within-household transmission probability), pi_hh (baseline attack rate), and p_base (baseline infection probability).

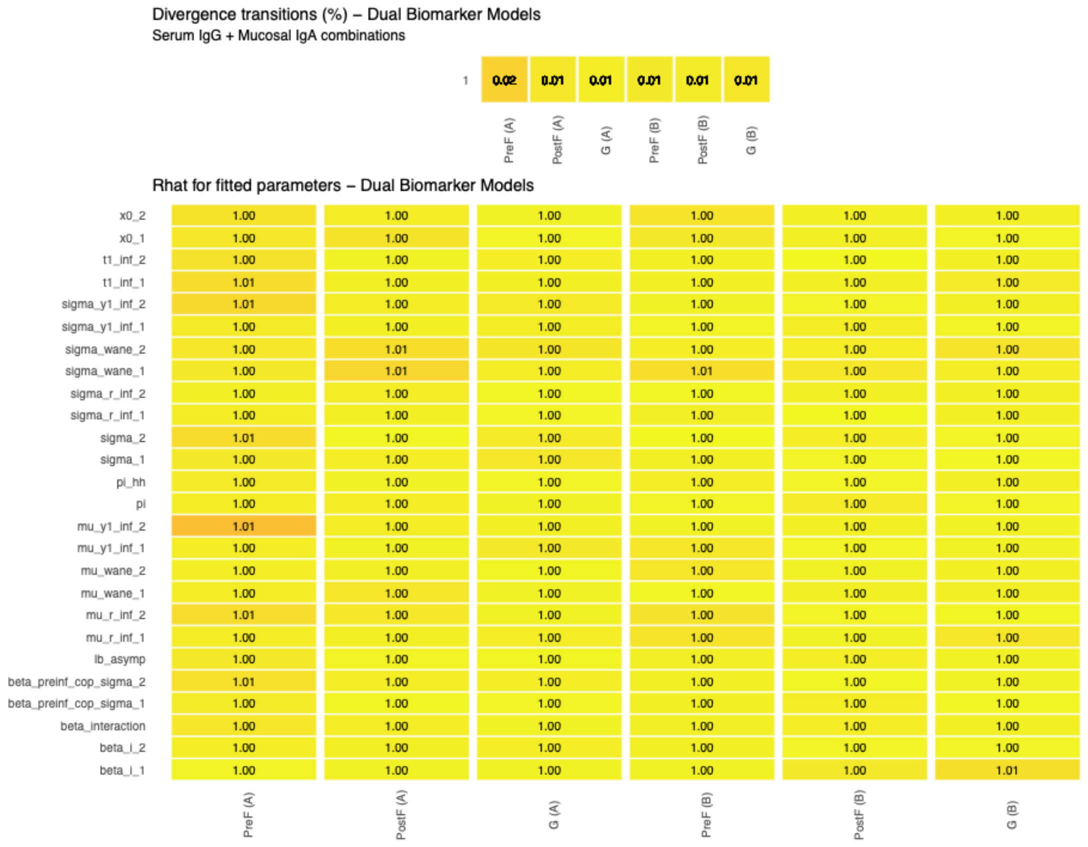


**Supplementary Results Figure 1.2. MCMC convergence diagnostics for 6 dual biomarker antibody kinetics and correlates of protection models. PCR-only.** Convergence diagnostics for dual biomarker Bayesian hierarchical models simultaneously estimating antibody kinetics and correlates of protection for serum IgG and mucosal IgA targeting the same viral protein. The top panel shows the percentage of divergent transitions (out of 4,000 total iterations across 4 chains) for each dual biomarker model. All models achieved divergent transition rates < 1%, with most < 0.5%. Parameters shown include biomarker-specific parameters (with subscripts _1 and _2 denoting serum IgG and mucosal IgA, respectively): waning kinetics (mu_wane, sigma_wane), boosting kinetics (mu_y1_inf, sigma_y1_inf, mu_r_inf, sigma_r_inf, t1_inf), measurement error (sigma), protection thresholds (x0), and correlates of protection effects (beta_i,

beta_preinf_cop_sigma), as well as the interaction term (beta_interaction) capturing synergistic or antagonistic effects between the two biomarkers. Shared parameters include lb_asymp (asymptotic lower bound), pi (within-household transmission), and pi_hh (baseline attack rate).

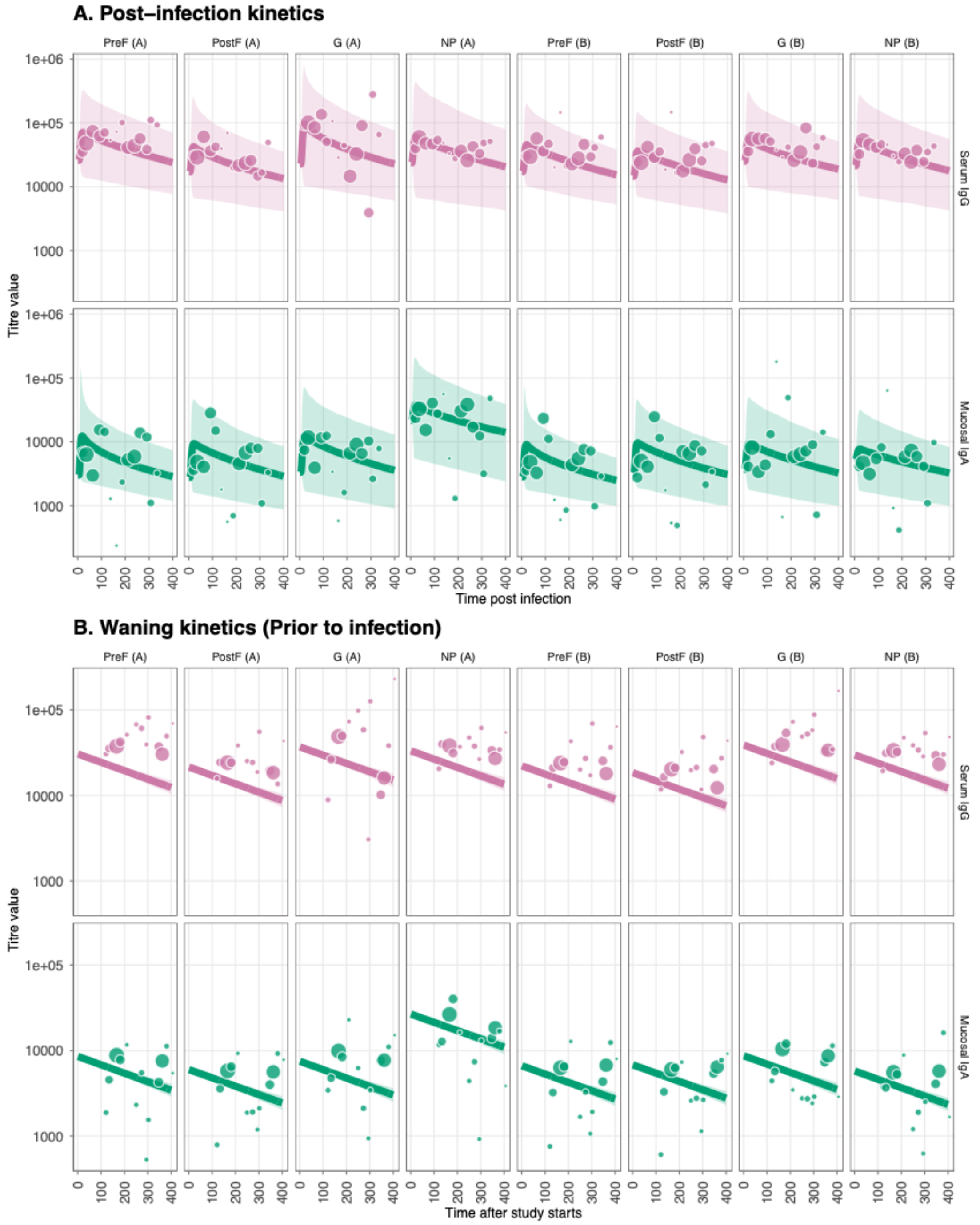


**Supplementary Results Figure 1.3. Antibody kinetics fits and immune responses for RSV-A and RSV-B post and prior to infection. PCR-only.** (A) Post-infection longitudinal antibody titers for eight viral RSV-A/B proteins (PreF, PostF, G, and NP) across different

antibody types; serum IgG and mucosal IgA. Lines show the median posterior predictive fit from the fitted Bayesian model, and the points show the observational titre data, with the size correlating with the sample size for that bin. (B) Prior-infection antibody waning for eight viral RSV-A/B proteins (PreF, PostF, G, and NP) across different antibody types; serum IgG and mucosal IgA.

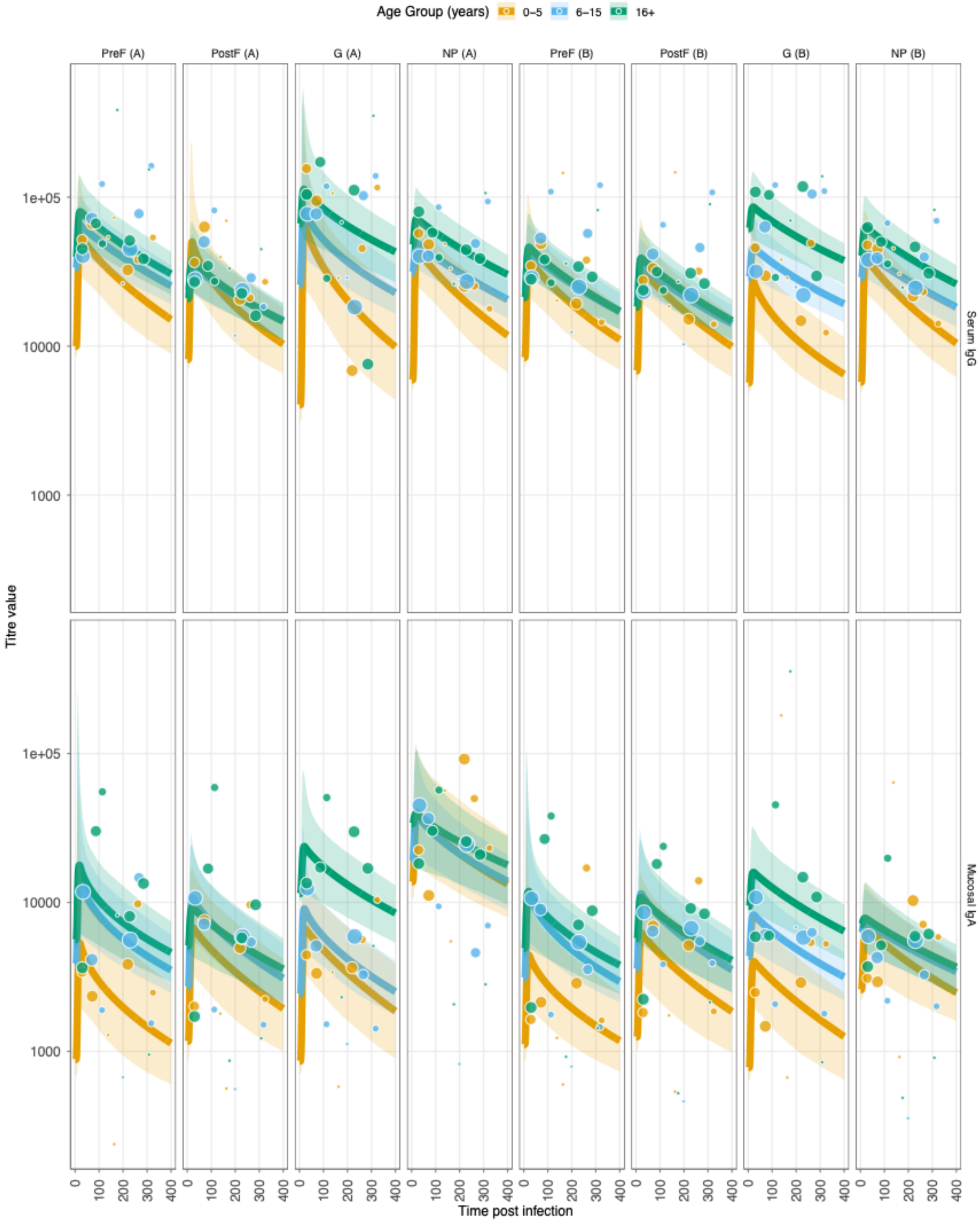

**Supplementary Results Figure 1.4. Age-stratified antibody kinetics fits and immune responses for RSV-A and RSV-B post and prior to infection. PCR-only.** Layout as **Supplementary Figure 1.3.**

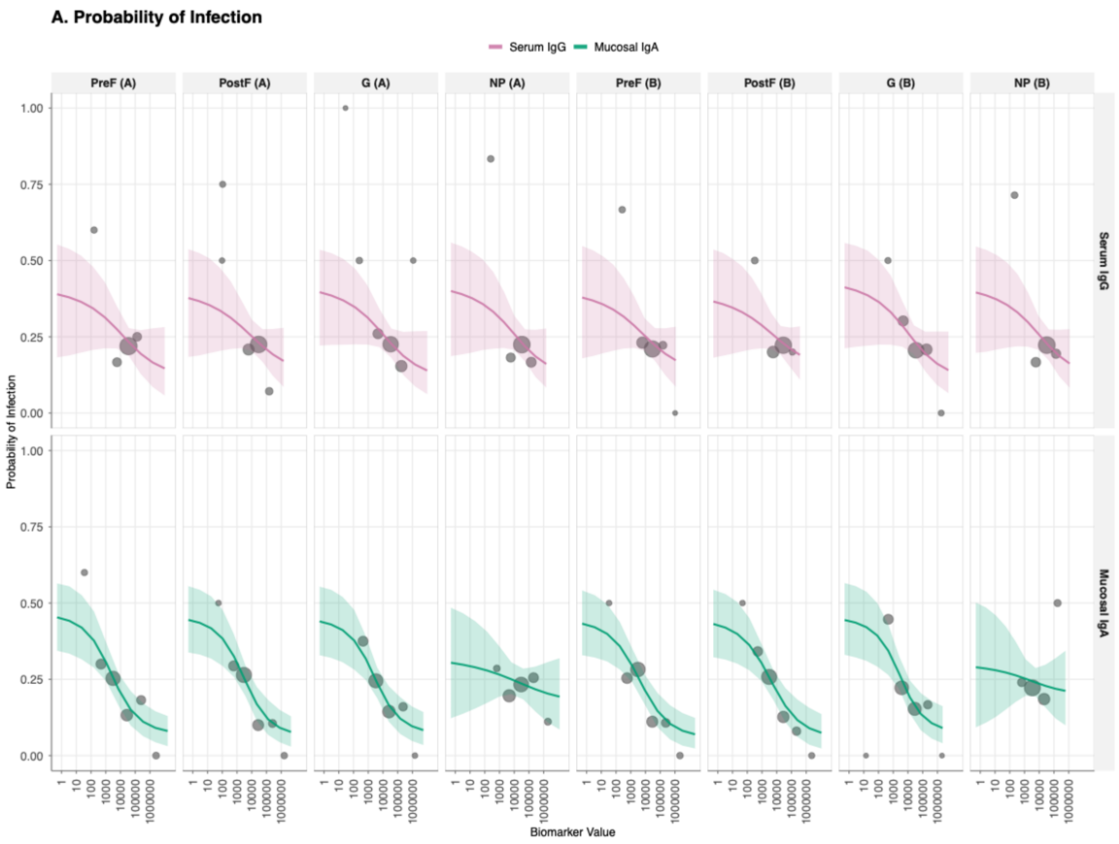


**Supplementary Results Figure 1.5. Correlate of risk (CoR) curves showing the relationship between antibody titre at infection and probability of protection from RSV infection for all 16 biomarker-antigen combinations. PCR-only.** Serum IgG, top row; mucosal IgA, bottom row and columns are viral antigen target (PreF, PostF, G, and NP for both RSV-A and RSV-B strains). The solid lines represents the mean estimated probability of protection given exposure to infection as a function of antibody titre, with shaded ribbons indicating 95% credible intervals, and the points show the observational titre at infection, with the size correlating with the sample size for that bin.

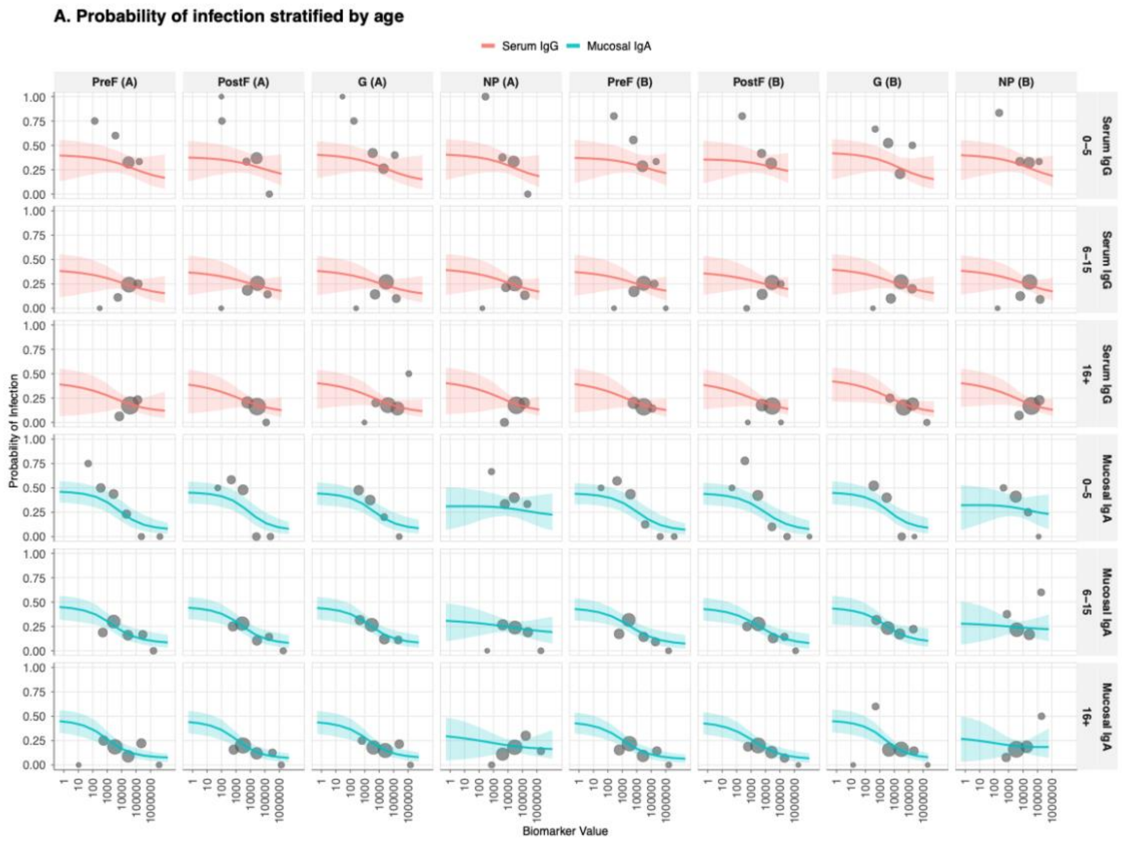


**Supplementary Results Figure 1.6. Age-stratified correlate of risk (CoR) curves showing the relationship between antibody titre at infection and probability of protection from RSV infection for all 16 biomarker-antigen combinations. PCR-only.** Layout as **Supplementary Figure 1.5.**

### B. RESULTS AND INFERENCE

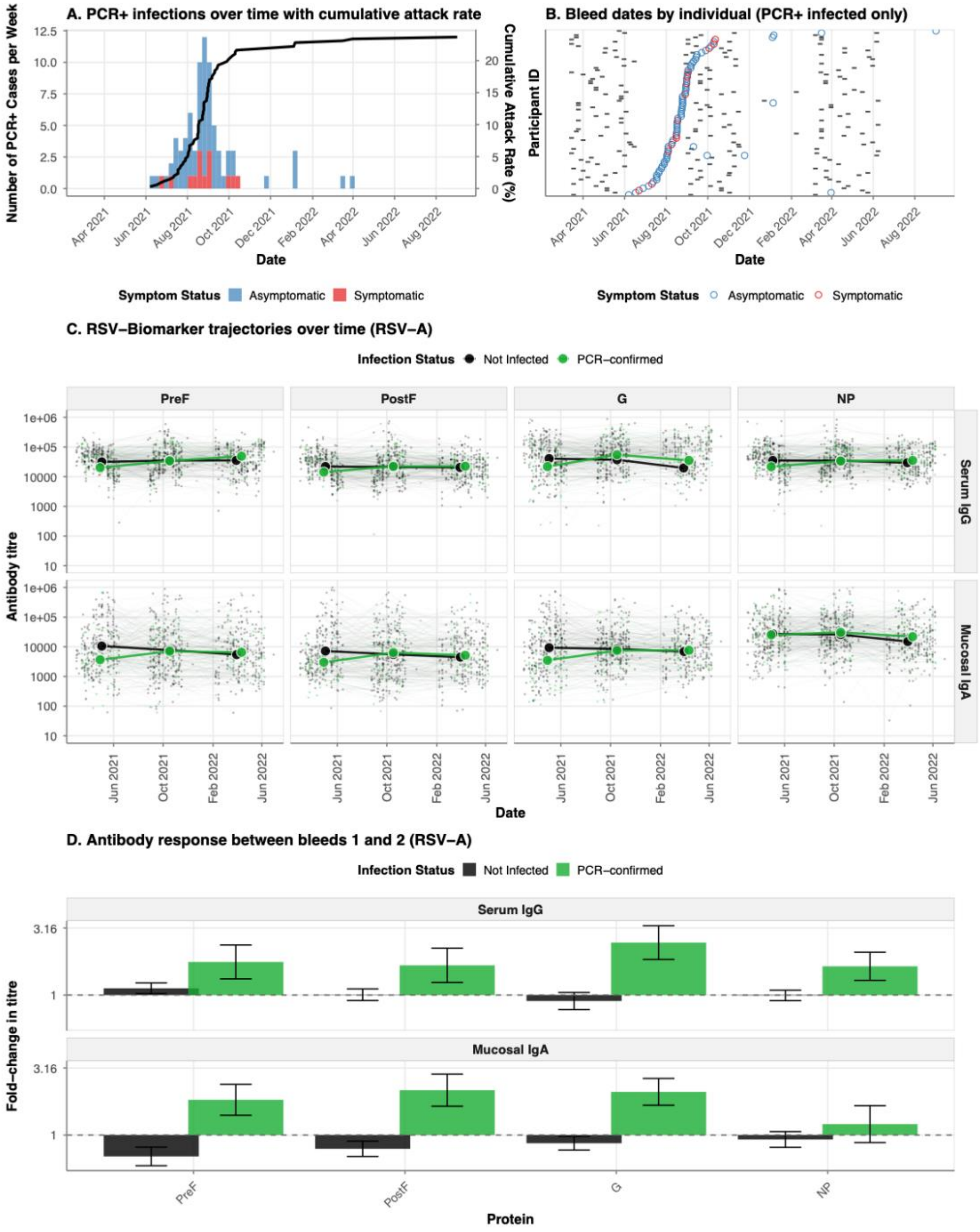


**Supplementary Results Figure 1.7. Epidemiology of RSV infections and antibody kinetics during the 2022-2023 epidemic season. PCR-only. A.** Weekly incidence of PCR-confirmed RSV infections (stacked histogram) and cumulative attack rate (black line) among 343 household cohort participants from October 2022 to May 2023. Bars are colored by symptom status at detection: symptomatic (red, detected at unscheduled visits) or asymptomatic (blue, detected at scheduled home or clinic visits). **B.** Timeline of serological sampling for PCR-confirmed infected individuals. Each horizontal line represents one participant (ordered by date of PCR-confirmed infection), with grey tiles indicating bleed dates and colored circles marking the timing of PCR-confirmed infection (colored by symptom status as in panel a). **C.** Longitudinal trajectories of serum IgG (top row) and mucosal IgA (bottom row) antibody responses against four RSV-A proteins (PreF, PostF, G, NP) over time. Individual participant trajectories are shown as thin lines with low opacity, colored by infection status: not infected (black), serologically-defined infections (orange), and PCR-confirmed infections (green). **D.** Mean fold-change in antibody titres between the first bleed (pre-epidemic baseline) and second bleed (post-epidemic) for serum IgG and mucosal IgA responses to RSV-A proteins. Bars represent mean fold-change (log10 scale) stratified by infection status, with error bars indicating standard error.

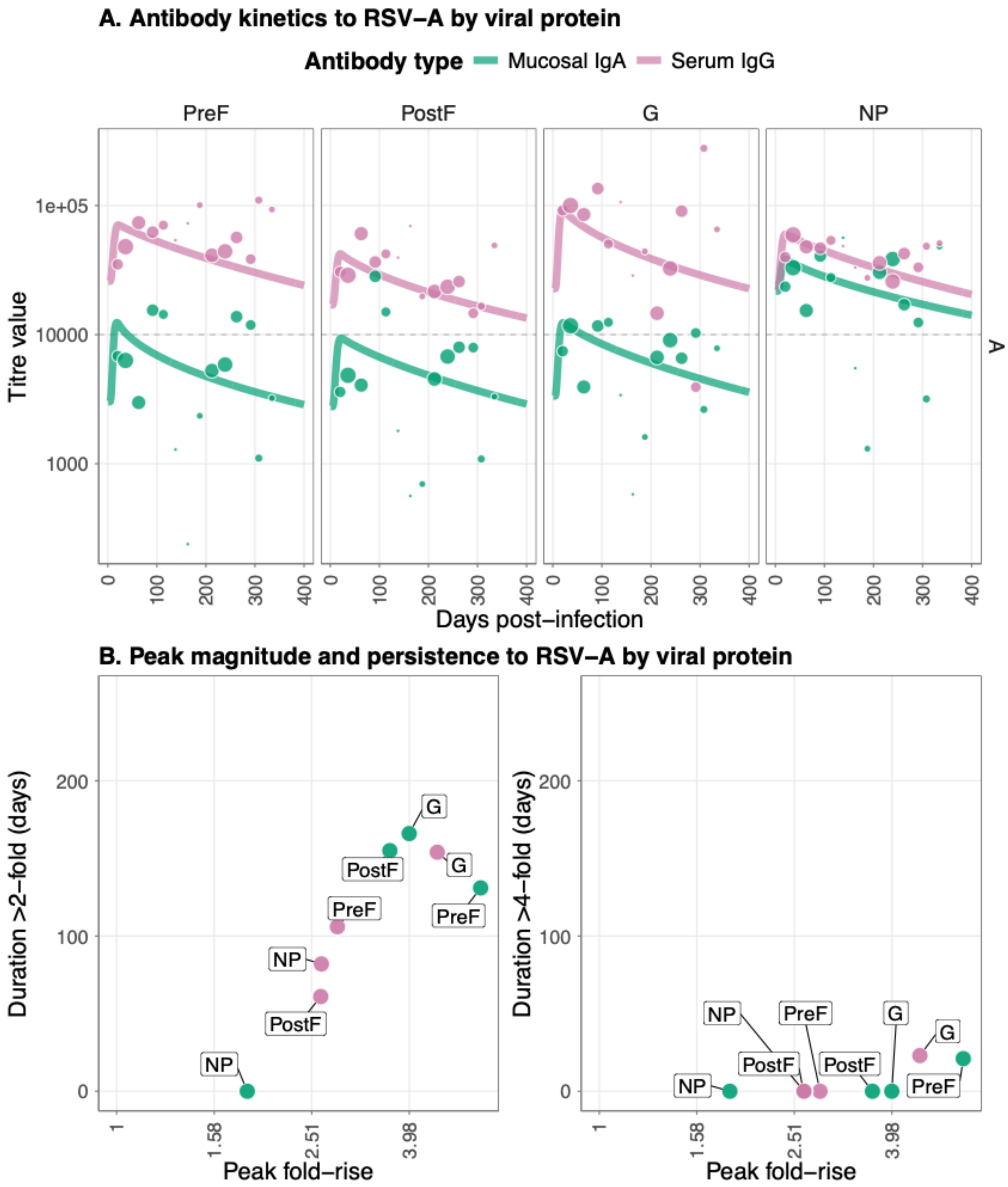


**Supplementary Results Figure 1.8. Antibody kinetics and immune responses for PCR-confirmed infections to RSV-A.** (A) Post-infection longitudinal antibody titers for four viral RSV-A proteins (PreF, PostF, G, and NP) across different antibody types; serum IgG and mucosal IgA. Lines show the median posterior predictive fit from the fitted Bayesian model, and the points show the observational titre data, with the size correlating with the sample size for that bin. (B) Peak antibody (x axis) and persistence measured as duration above a 2-fold (left panel) and 4-fold titre rise (right panel) in days. Data points show median posterior values for measurements of four viral RSV-A proteins (PreF, PostF, G, and NP) across different antibody types: serum IgG and mucosal IgA.

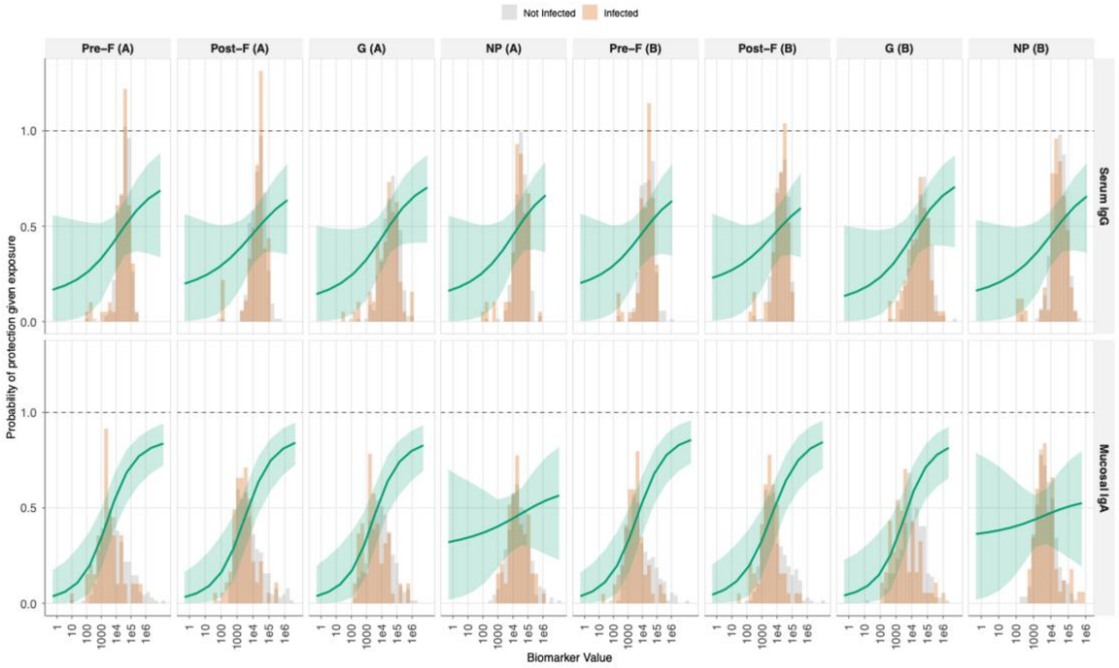


**Supplementary Results Figure 1.9. Correlate of protection (CoP) curves showing the relationship between antibody titre at infection and probability of protection from RSV infection for all 16 biomarker-antigen combinations.** Serum IgG, top row; mucosal IgA, bottom row and columns are viral antigen target (PreF, PostF, G, and NP for both RSV-A and RSV-B strains). The solid green line represents the mean estimated probability of protection given exposure to infection as a function of antibody titre, with shaded ribbons indicating 95% credible intervals. Background histograms show the distribution of antibody titres at infection for infected individuals (orange) versus non-infected individuals (gray).

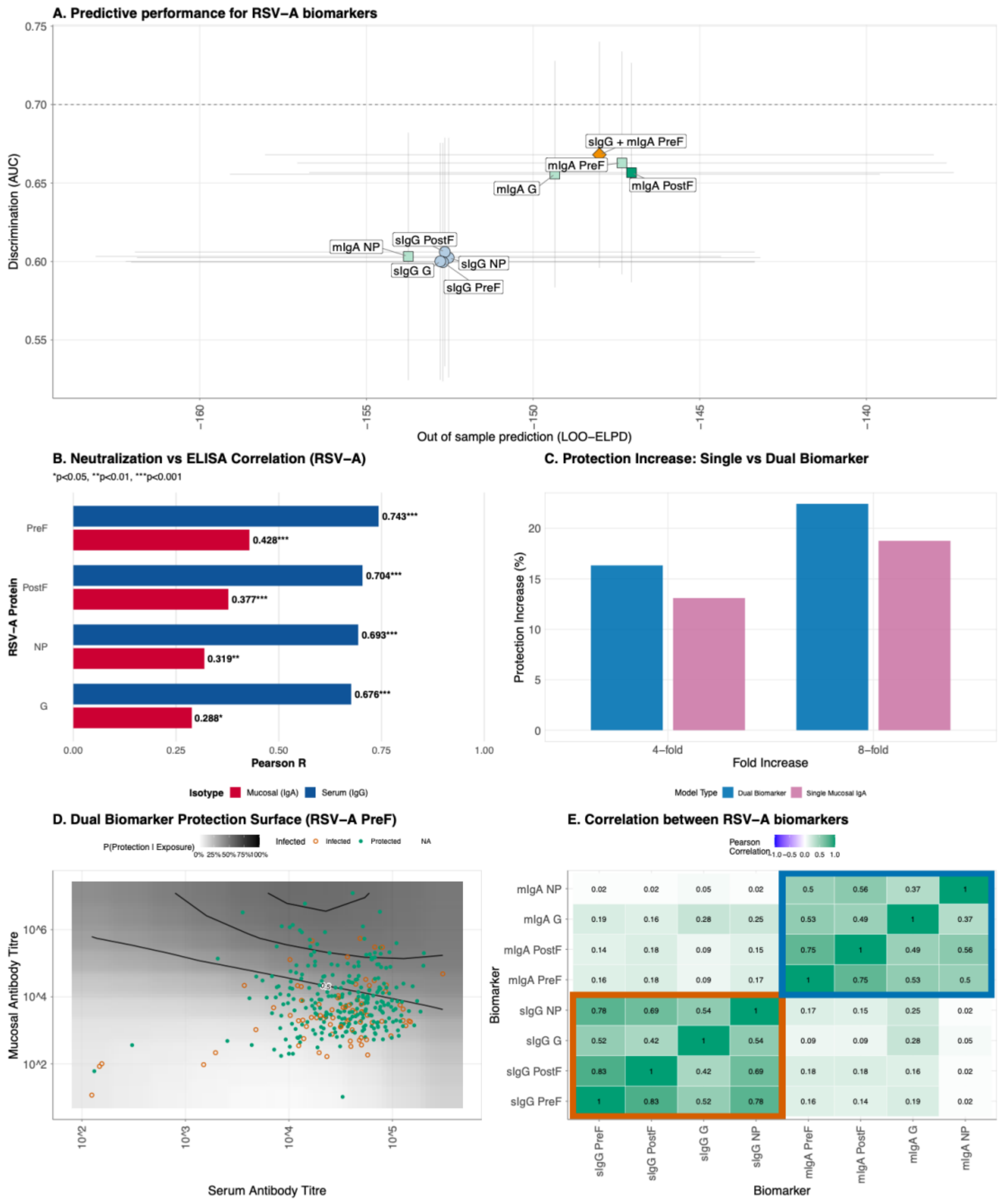


**Supplementary Results Figure 1.10. Comparison of single and dual biomarker models for predicting protection against RSV-A PCR-confirmed infection.** (A) Model performance comparison across single biomarker models and the dual biomarker model, defined by out-of-sample predictive accuracy (LOO-ELPD, x-axis) and discrimination ability (area under the ROC curve, AUC, y-axis). Circles indicate serum IgG models, squares represent mucosal IgA models, and the triangle denotes the dual biomarker model combining serum IgG and mucosal IgA to RSV-A PreF. The best-performing model within each biomarker class is highlighted with darker shading. Error bars show the standard error of LOO-ELPD (horizontal) and 95% confidence intervals for AUC (vertical).

The dashed horizontal line indicates an AUC of 0.7. (B) Summary of Pearson correlation coefficients for RSV-A protein antigens, comparing serum IgG (blue) and mucosal IgA (red) ELISA titres with neutralisation IC50 values. (C) Counterfactual analysis comparing the increase in protection from boosting pre-infection antibody titres by 4-fold or 8-fold using either a single mucosal IgA biomarker model (purple) or a dual biomarker model combining mucosal IgA and serum IgG (blue). (D) Two-dimensional protection surface for the dual biomarker model using serum IgG and mucosal IgA antibodies to RSV-A PreF. The continuous surface represents the predicted probability of protection given exposure as a function of both serum (x-axis) and mucosal (y-axis) antibody titres. Black contour lines delineate regions of constant protection probability at 50%, 70%, 80%, and 90%. Individual data points overlay the surface, with filled circles representing participants who were protected despite exposure and open circles representing those who became infected. Antibody titres are displayed on a log10 scale. (E) Pearson correlation coefficients between RSV-A variant biomarkers at the time of infection. Color intensity represents correlation strength, ranging from blue (weak correlation) through white (no correlation) to green (strong positive correlation).

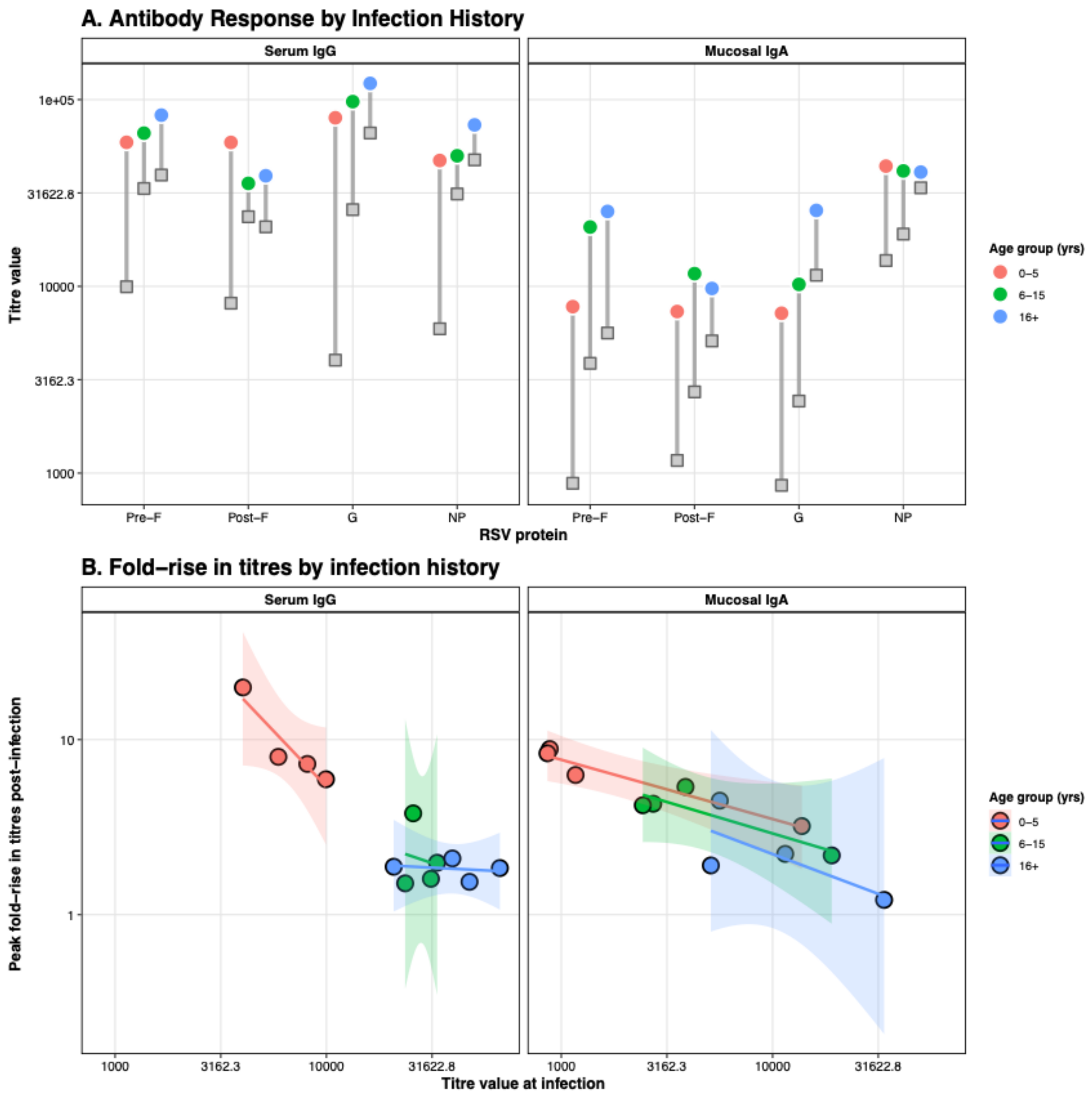


**Supplementary Results Figure 1.11. Age-dependent patterns in antibody magnitude and boosting following natural RSV infection. PCR-only** (A) Baseline and peak antibody titres stratified by age group for serum IgG (left panel) and mucosal IgA (right panel) against four RSV-A antigens (PreF, PostF, G, NP). Gray squares represent baseline titres at the time of infection, colored circles represent peak titres post-infection, and gray lines connecting them indicate the magnitude of antibody boosting. Point are colored according to age groups. (B) Relationship between baseline antibody titre at infection (x-axis) and fold-rise in titre post-infection (y-axis), illustrating the antibody ceiling effect where individuals with higher pre-existing titres exhibit smaller relative boosts. Linear regression lines (solid) with 95% confidence intervals (shaded regions) demonstrate the negative relationship.

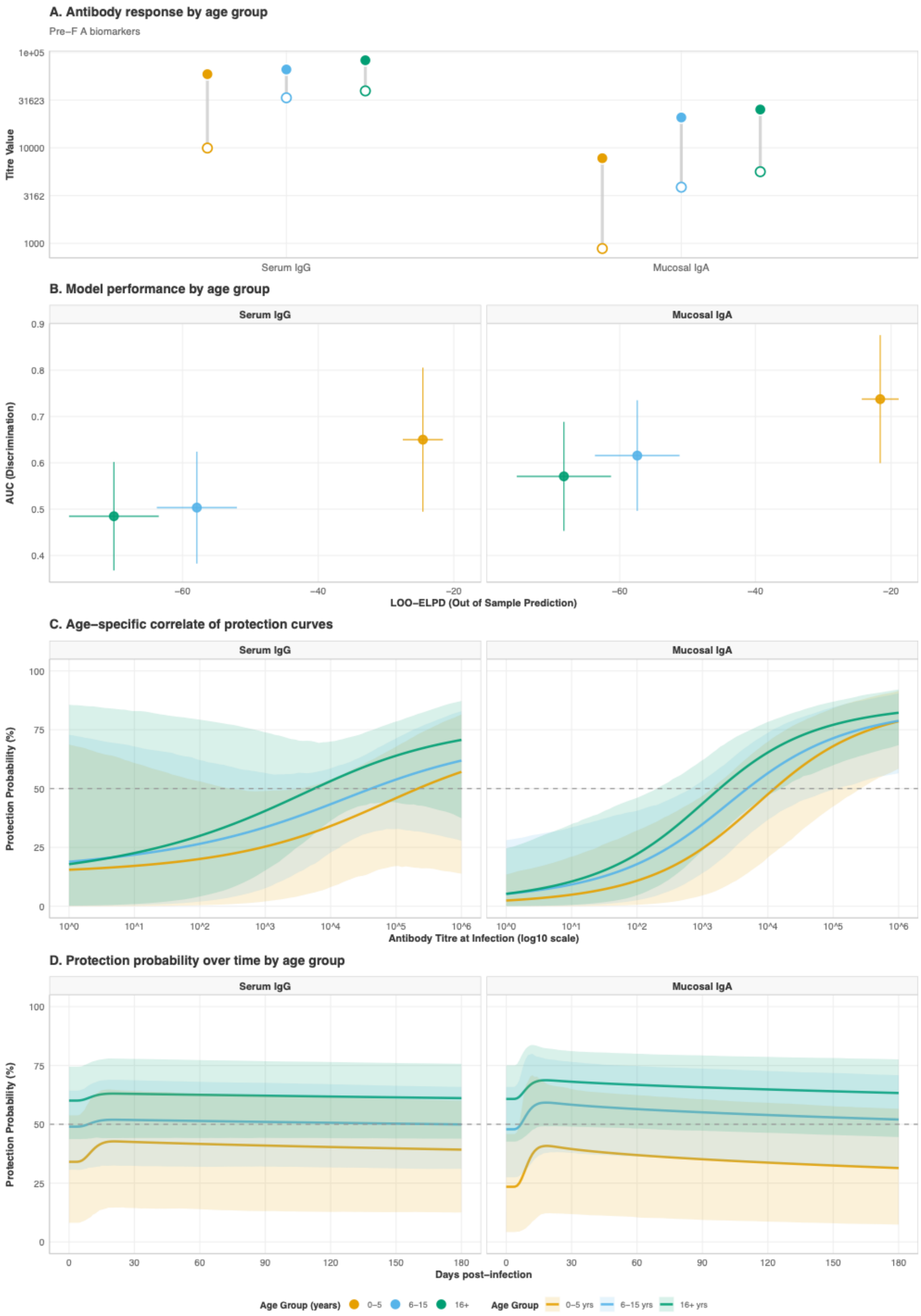


**Supplementary Results Figure 1.12. Age-stratified analysis of antibody responses and protective PCR-confirmed immunity for RSV-A PreF biomarkers.** (A) Antibody response

magnitudes by age group, with baseline antibody titres at time of infection (hollow circles) and peak titres following infection (filled circles) for serum IgG and mucosal IgA antibodies to RSV-A PreF protein, stratified by age group (0-5 years in orange, 6-15 years in light blue, 16+ years in green). (B) Model performance by age group. The scatter plot compares out-of-sample predictive accuracy (LOO-ELPD, x-axis) and discrimination ability (AUC, y-axis) for correlate of protection models fitted separately within each age stratum.. Vertical error bars show 95% confidence intervals for AUC; horizontal error bars indicate standard error of LOO-ELPD. (C). Age-specific correlate of protection curves demonstrating the relationship between antibody titre at infection and probability of protection. (D) Protection probability trajectories by age group following infection. The lines show the predicted probability of protection against reinfection over time (0-180 days post-infection) for serum IgG (left panel) and mucosal IgA (right panel) antibodies to RSV-A PreF. Solid lines represent the median posterior prediction for each age group, with shaded ribbons indicating 95% credible intervals.

---

### 2. PCR + SERO MODEL $\rho = 0.9$

#### A. CONVERGENCE AND MODEL FITS

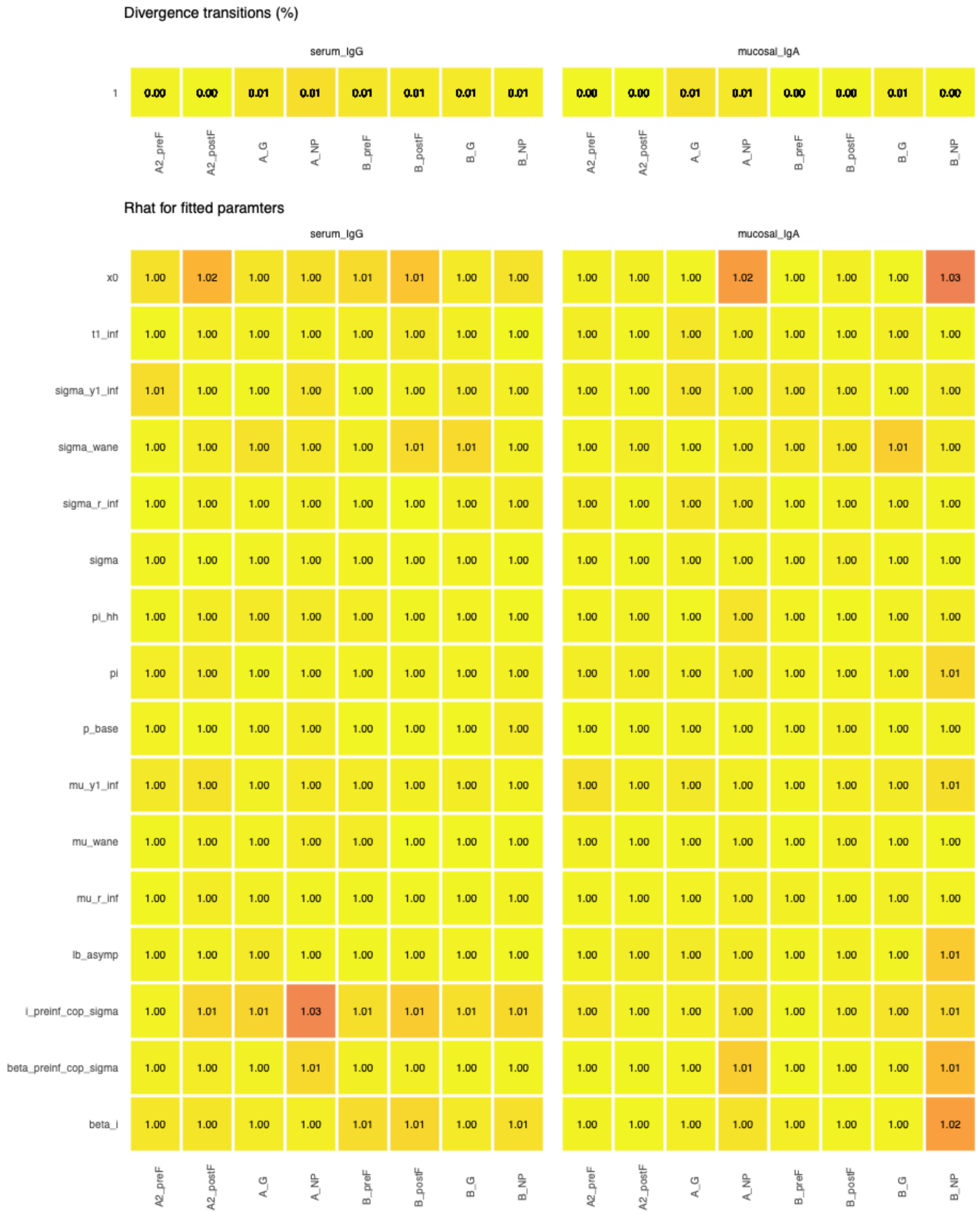


**Supplementary Results Figure 2.1. MCMC convergence diagnostics for 16 single biomarker antibody kinetics and correlates of protection models. $\rho = 0.9$.** Convergence diagnostics for Bayesian hierarchical models estimating antibody kinetics and correlates of protection. The top panel shows the percentage of divergent transitions

(out of 4,000 total iterations across 4 chains) for each biomarker-protein combination. All models achieved divergent transition rates < 2%, indicating successful exploration of the posterior distribution. Parameters shown include: mu_wane and sigma_wane (waning kinetics), mu_y1_inf and sigma_y1_inf (peak fold-rise), mu_r_inf and sigma_r_inf (boosting rate), t1_inf (time to peak), sigma (measurement error), lb_asymp (asymptotic lower bound), x0 (protection threshold), i_preinf_cop_sigma (pre-infection titre variability effect), beta_i (infection effect on susceptibility), beta_preinf_cop_sigma (pre-infection titre effect on protection), pi (within-household transmission probability), pi_hh (baseline attack rate), and p_base (baseline infection probability).

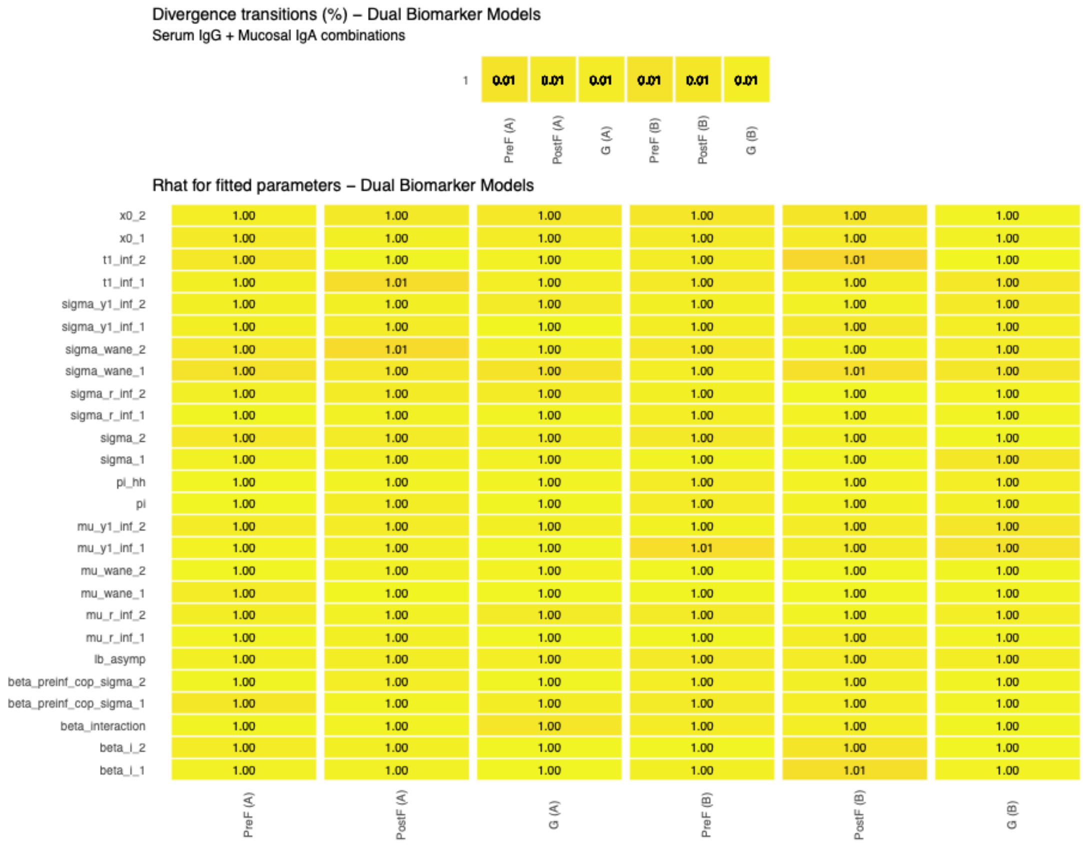


**Supplementary Results Figure 2.2. MCMC convergence diagnostics for 6 dual biomarker antibody kinetics and correlates of protection models. $\rho = 0.9$.** Convergence diagnostics for dual biomarker Bayesian hierarchical models simultaneously estimating antibody kinetics and correlates of protection for serum IgG and mucosal IgA targeting the same viral protein. The top panel shows the percentage of divergent transitions (out of 4,000 total iterations across 4 chains) for each dual biomarker model. All models achieved divergent transition rates < 1%, with most < 0.5%. Parameters shown include biomarker-specific parameters (with subscripts _1 and _2 denoting serum IgG and mucosal IgA, respectively): waning kinetics (mu_wane, sigma_wane), boosting kinetics (mu_y1_inf, sigma_y1_inf, mu_r_inf, sigma_r_inf, t1_inf), measurement error (sigma), protection thresholds (x0), and correlates of protection effects (beta_i,

beta_preinf_cop_sigma), as well as the interaction term (beta_interaction) capturing synergistic or antagonistic effects between the two biomarkers. Shared parameters include lb_asymp (asymptotic lower bound), pi (within-household transmission), and pi_hh (baseline attack rate).

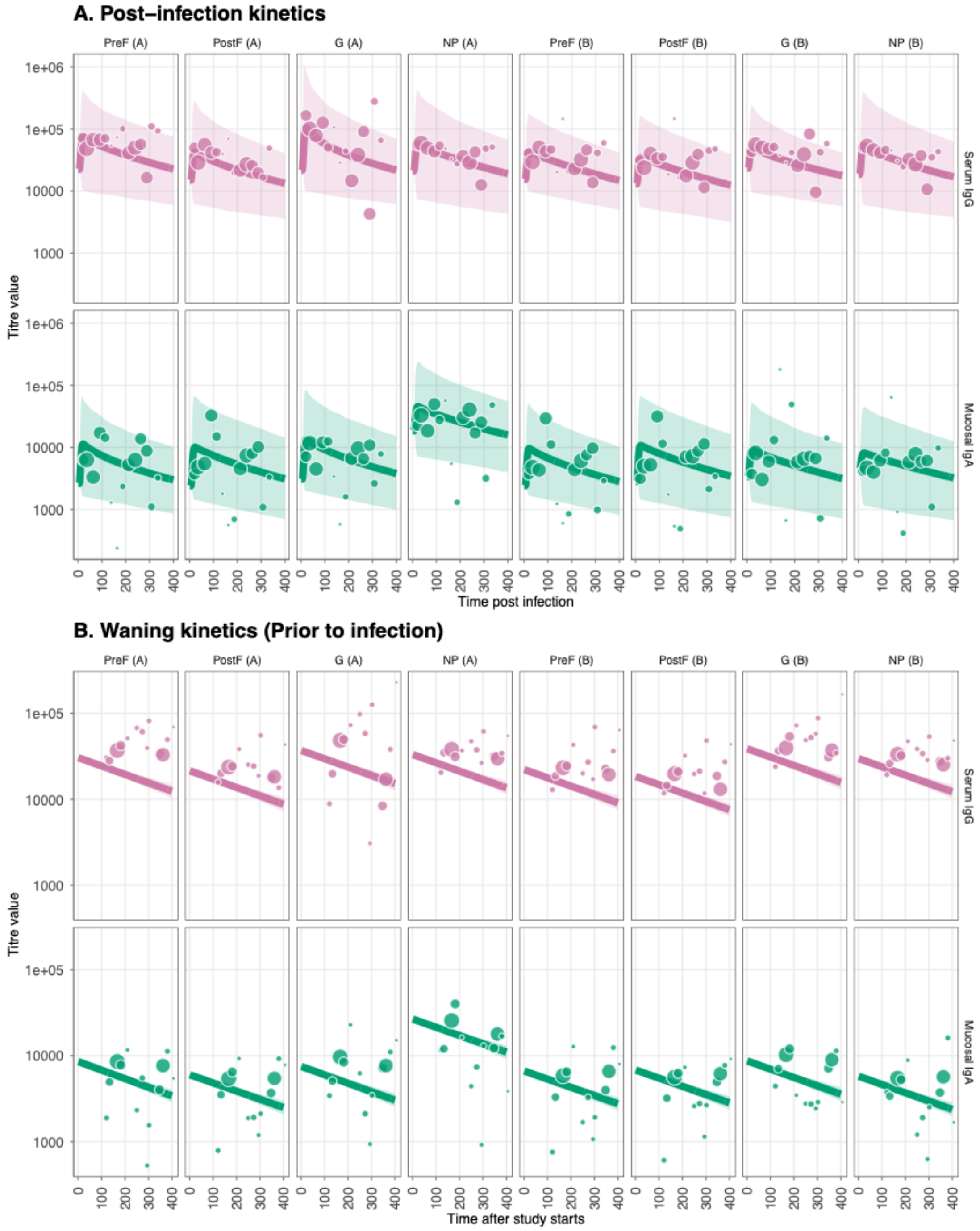


**Supplementary Results Figure 2.3. Antibody kinetics fits and immune responses for RSV-A and RSV-B post and prior to infection.** $\rho = 0.9$ (A) Post-infection longitudinal antibody titers for eight viral RSV-A/B proteins (PreF, PostF, G, and NP) across different

antibody types; serum IgG and mucosal IgA. Lines show the median posterior predictive fit from the fitted Bayesian model, and the points show the observational titre data, with the size correlating with the sample size for that bin. (B) Prior-infection antibody waning for eight viral RSV-A/B proteins (PreF, PostF, G, and NP) across different antibody types; serum IgG and mucosal IgA.

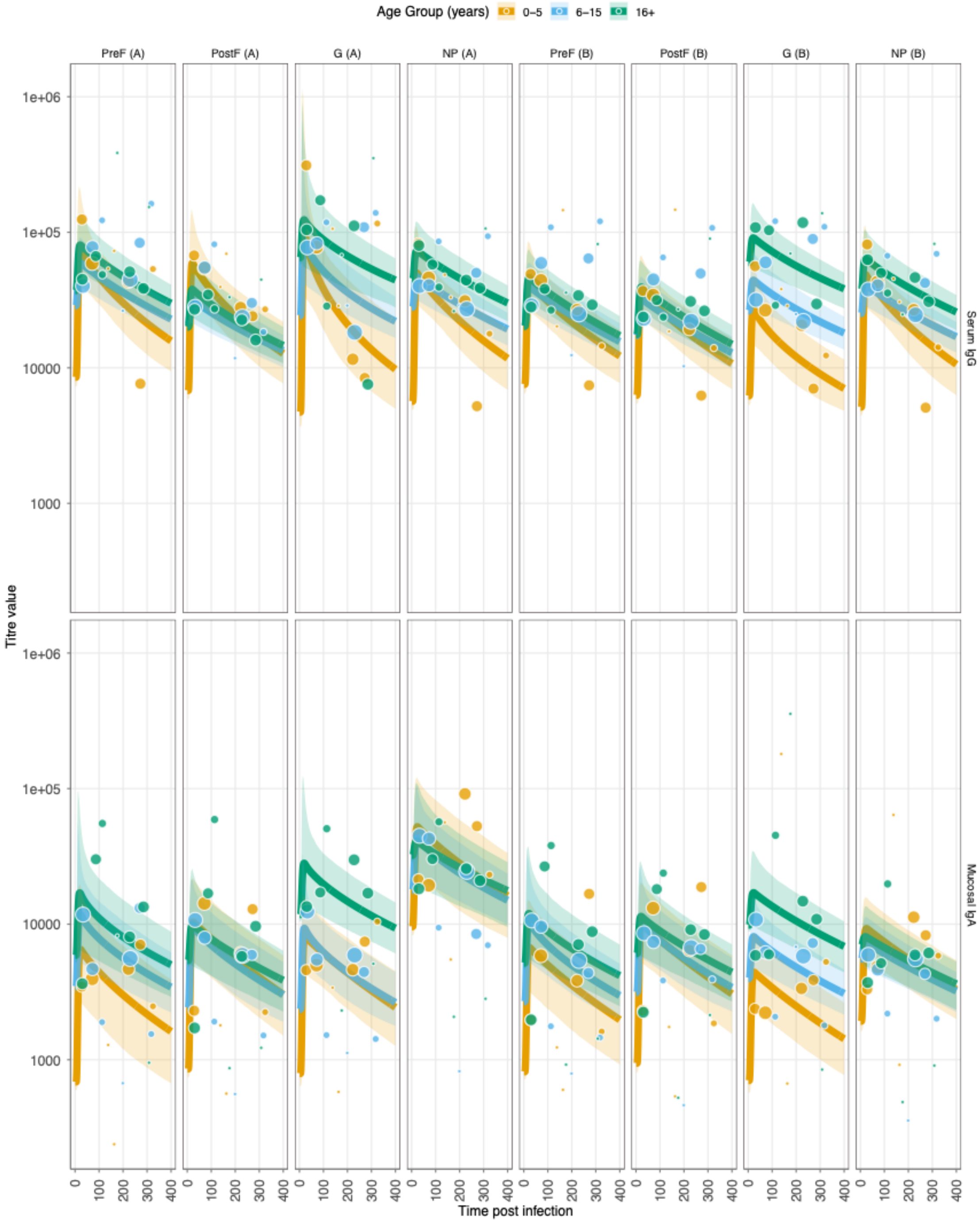

**Supplementary Results Figure 2.4. Age-stratified antibody kinetics fits and immune responses for RSV-A and RSV-B post and prior to infection. $\rho = 0.9$.** Layout as **Supplementary Figure 2.3.**

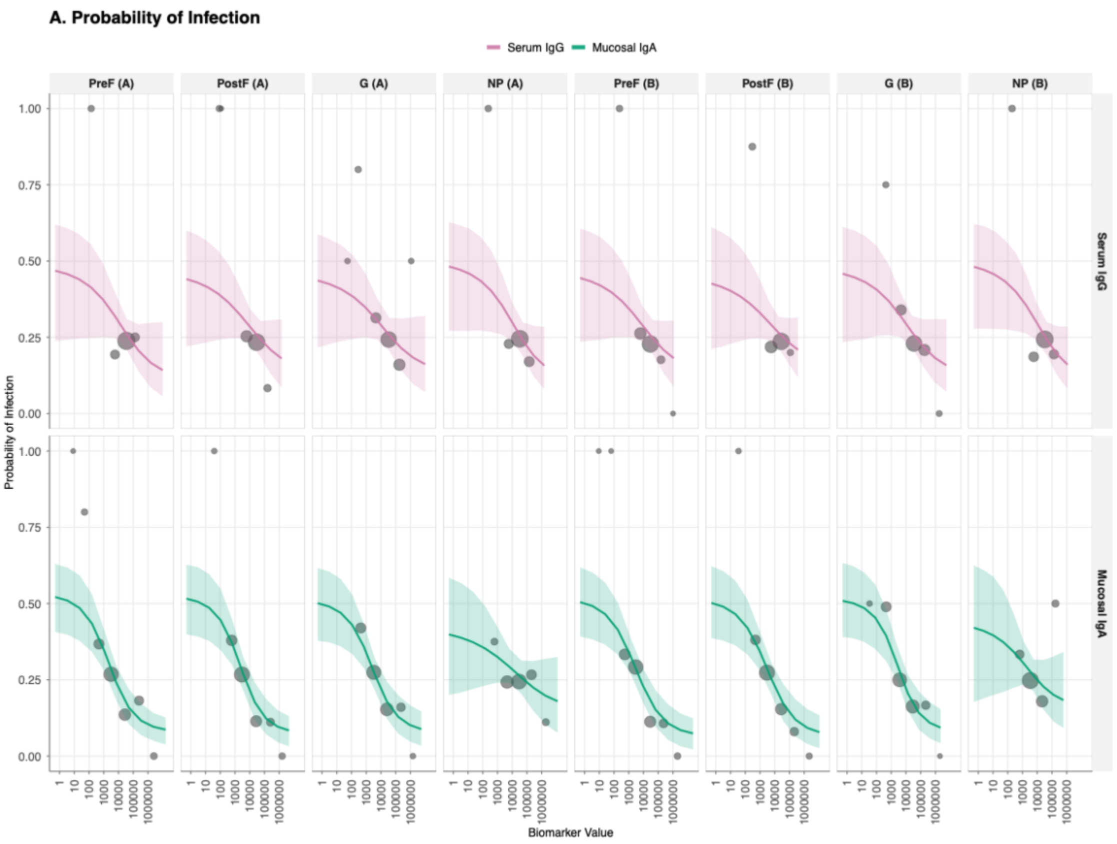


**Supplementary Results Figure 2.5. Correlate of risk (CoR) curves showing the relationship between antibody titre at infection and probability of protection from RSV infection for all 16 biomarker-antigen combinations. $\rho = 0.9$.** Serum IgG, top row; mucosal IgA, bottom row and columns are viral antigen target (PreF, PostF, G, and NP for both RSV-A and RSV-B strains). The solid lines represents the mean estimated probability of protection given exposure to infection as a function of antibody titre, with shaded ribbons indicating 95% credible intervals, and the points show the observational titre at infection, with the size correlating with the sample size for that bin.

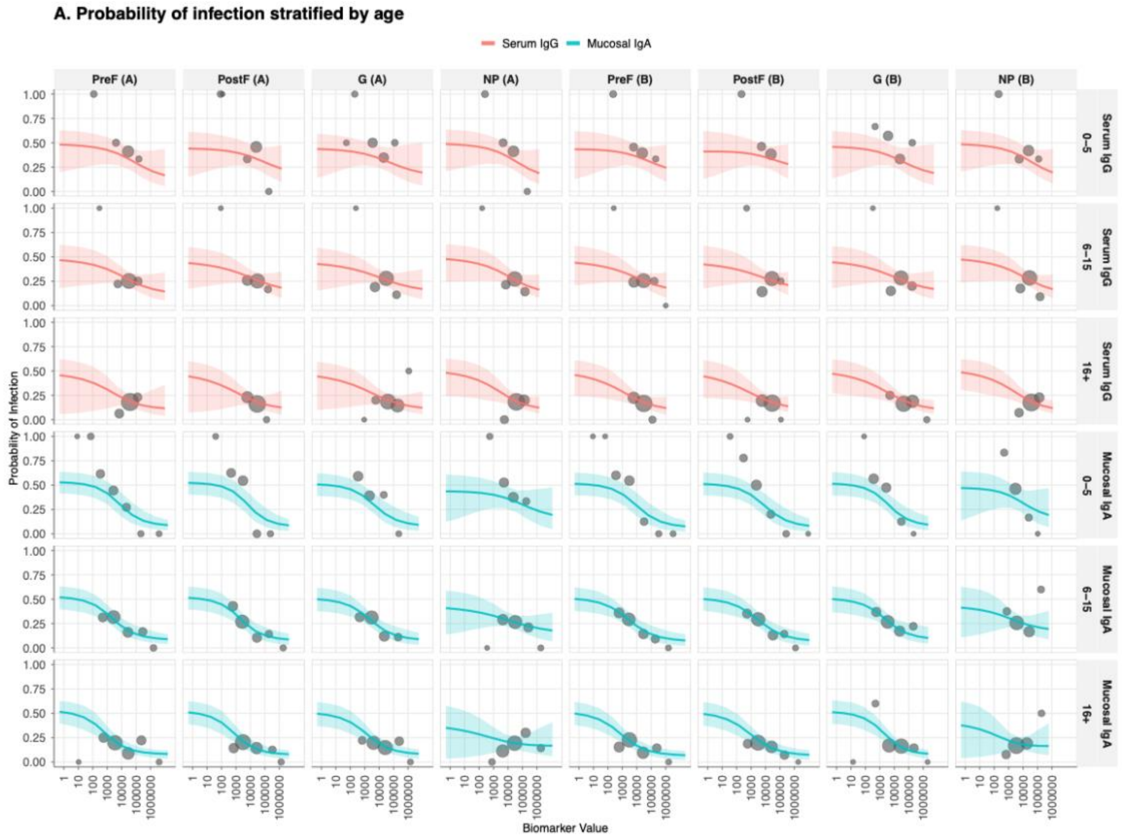


**Supplementary Results Figure 2.6. Age-stratified correlate of risk (CoR) curves showing the relationship between antibody titre at infection and probability of protection from RSV infection for all 16 biomarker-antigen combinations. $\rho = 0.9$.** Layout as **Supplementary Figure 1.5.**

### B. RESULTS AND INFERENCE

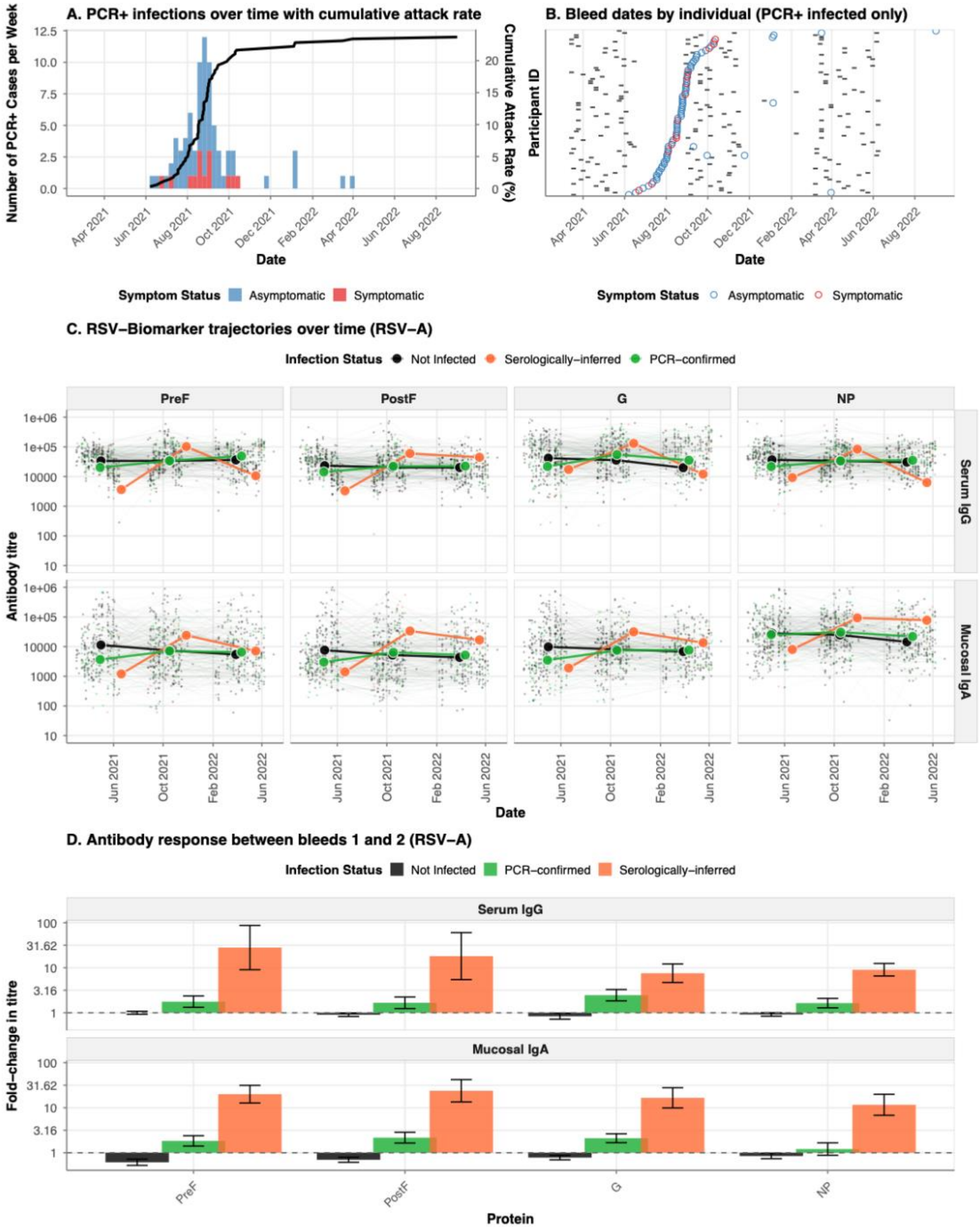


**Supplementary Results Figure 2.7. Epidemiology of RSV infections and antibody kinetics during the 2022-2023 epidemic season. $\rho = 0.1$ A.** Weekly incidence of PCR-confirmed RSV infections (stacked histogram) and cumulative attack rate (black line) among 343 household cohort participants from October 2022 to May 2023. Bars are colored by symptom status at detection: symptomatic (red, detected at unscheduled

visits) or asymptomatic (blue, detected at scheduled home or clinic visits). B. Timeline of serological sampling for PCR-confirmed infected individuals. Each horizontal line represents one participant (ordered by date of PCR-confirmed infection), with grey tiles indicating bleed dates and colored circles marking the timing of PCR-confirmed infection (colored by symptom status as in panel a). **C.** Longitudinal trajectories of serum IgG (top row) and mucosal IgA (bottom row) antibody responses against four RSV-A proteins (PreF, PostF, G, NP) over time. Individual participant trajectories are shown as thin lines with low opacity, colored by infection status: not infected (black), serologically-defined infections (orange), and PCR-confirmed infections (green). **D.** Mean fold-change in antibody titres between the first bleed (pre-epidemic baseline) and second bleed (post-epidemic) for serum IgG and mucosal IgA responses to RSV-A proteins. Bars represent mean fold-change (log10 scale) stratified by infection status, with error bars indicating standard error.

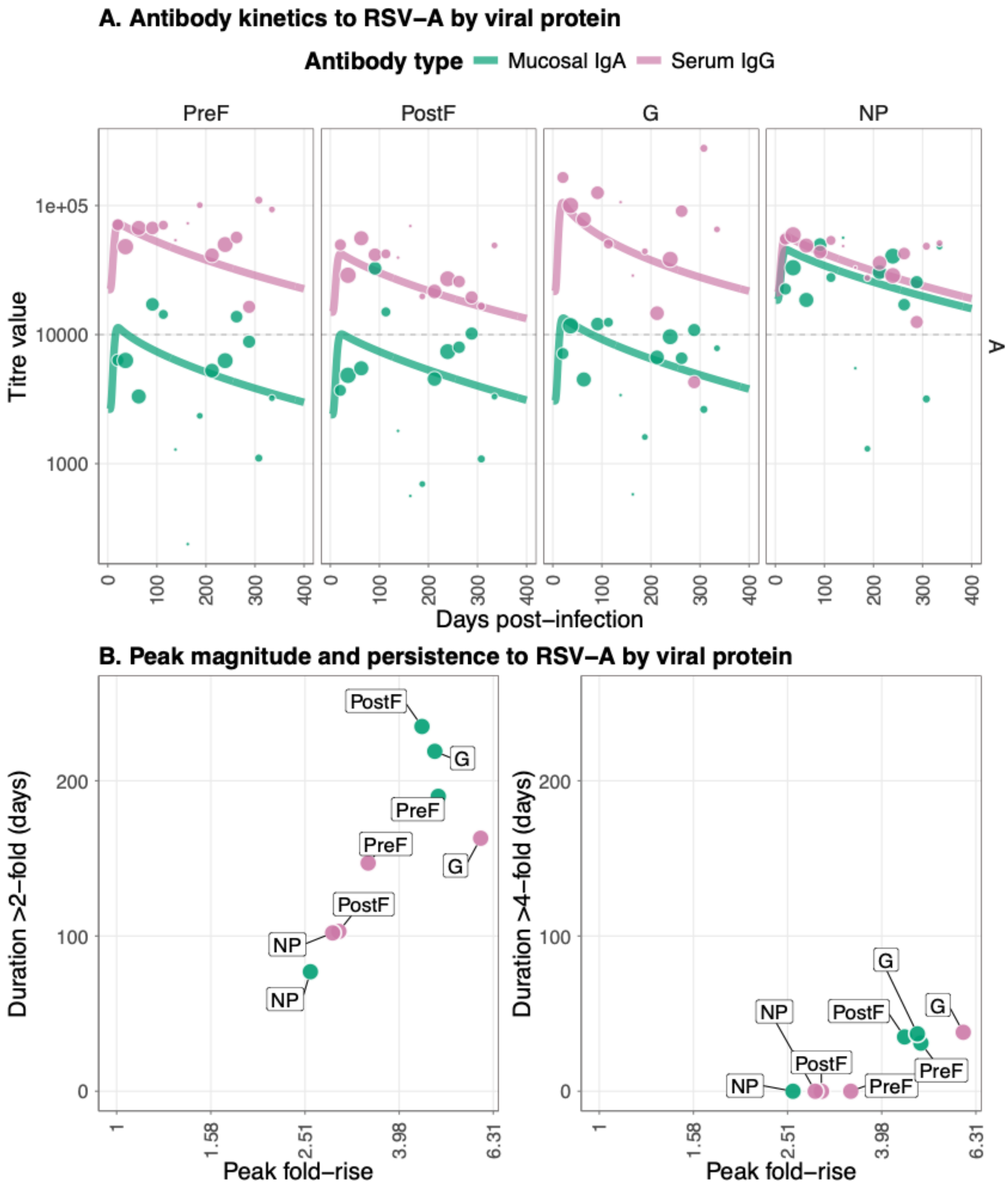


**Supplementary Results Figure 2.8. Antibody kinetics and immune responses for infections to RSV-A with $\rho = 0.9$** (A) Post-infection longitudinal antibody titers for four viral RSV-A proteins (PreF, PostF, G, and NP) across different antibody types; serum IgG and mucosal IgA. Lines show the median posterior predictive fit from the fitted Bayesian model, and the points show the observational titre data, with the size correlating with the sample size for that bin. (B) Peak antibody (x axis) and persistence measured as duration above a 2-fold (left panel) and 4-fold titre rise (right panel) in days. Data points show median posterior values for measurements of four viral RSV-A proteins (PreF, PostF, G, and NP) across different antibody types: serum IgG and mucosal IgA.

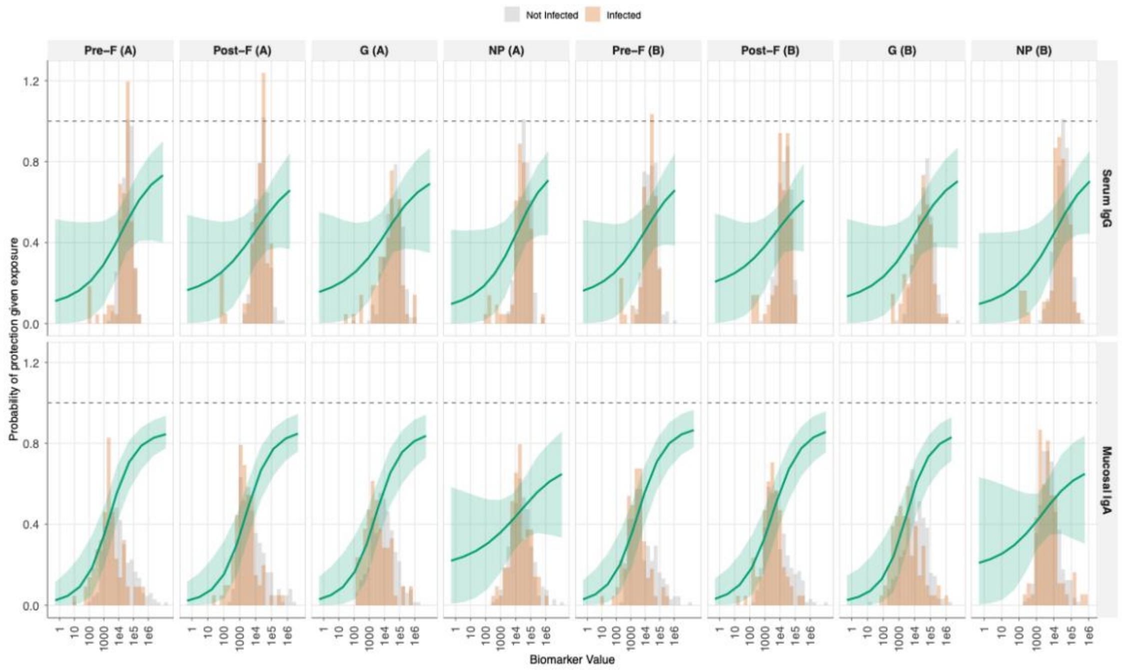


**Supplementary Results Figure 2.9. Correlate of protection (CoP) curves showing the relationship between antibody titre at infection and probability of protection from RSV infection for all 16 biomarker-antigen combinations. . $\rho = 0.9$** Serum IgG, top row; mucosal IgA, bottom row and columns are viral antigen target (PreF, PostF, G, and NP for both RSV-A and RSV-B strains). The solid green line represents the mean estimated probability of protection given exposure to infection as a function of antibody titre, with shaded ribbons indicating 95% credible intervals. Background histograms show the distribution of antibody titres at infection for infected individuals (orange) versus non-infected individuals (gray).

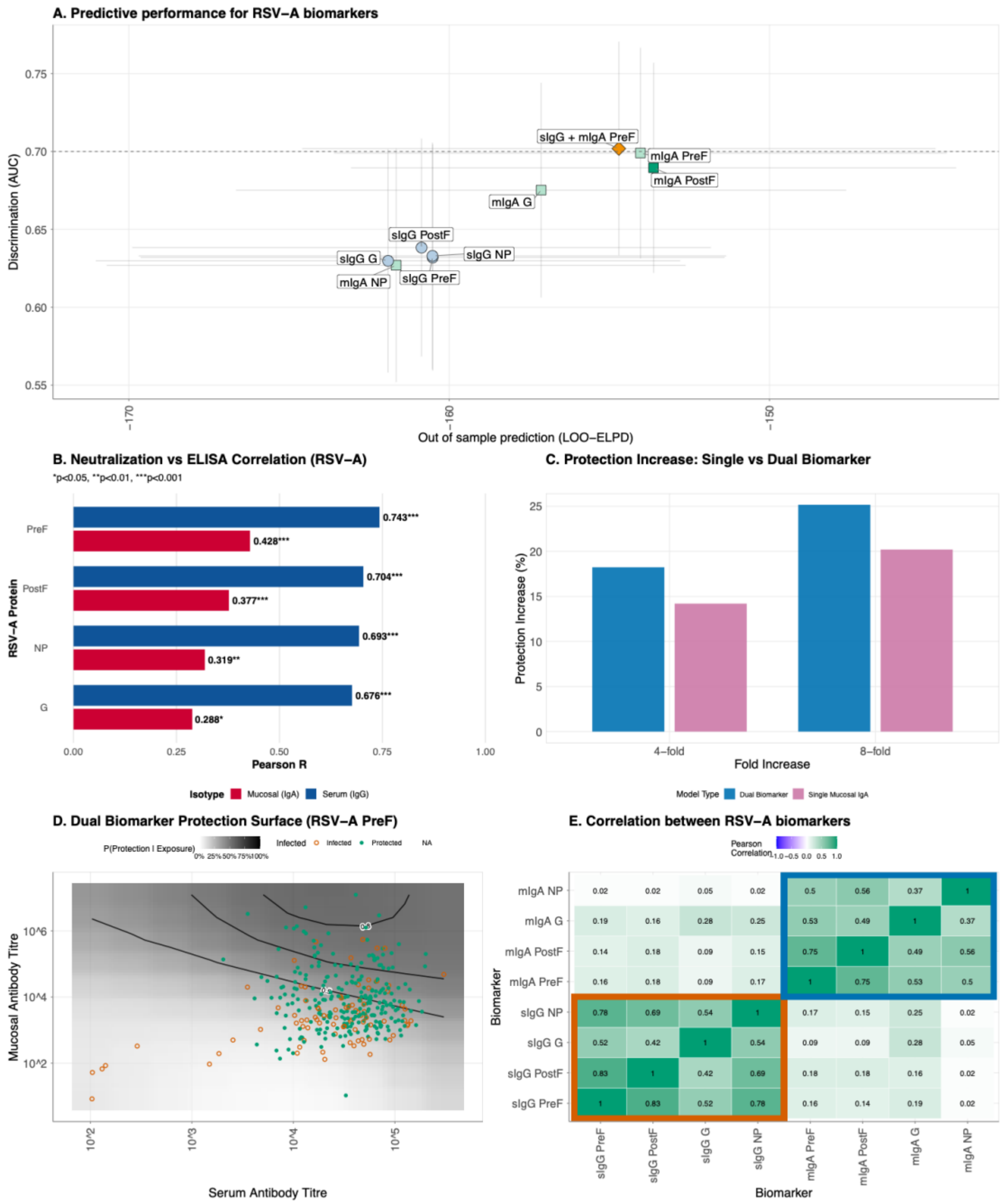


**Supplementary Results Figure 2.10. Comparison of single and dual biomarker models for predicting protection against RSV-A . $\rho = 0.9$ infection.** (A) Model performance comparison across single biomarker models and the dual biomarker model, defined by out-of-sample predictive accuracy (LOO-ELPD, x-axis) and discrimination ability (area under the ROC curve, AUC, y-axis). Circles indicate serum IgG models, squares represent mucosal IgA models, and the triangle denotes the dual biomarker model combining serum IgG and mucosal IgA to RSV-A PreF. The best-performing model within each biomarker class is highlighted with darker shading. Error bars show the standard error of LOO-ELPD (horizontal) and 95% confidence intervals for AUC (vertical). The dashed horizontal line

indicates an AUC of 0.7. (B) Summary of Pearson correlation coefficients for RSV-A protein antigens, comparing serum IgG (blue) and mucosal IgA (red) ELISA titres with neutralisation IC50 values. (C) Counterfactual analysis comparing the increase in protection from boosting pre-infection antibody titres by 4-fold or 8-fold using either a single mucosal IgA biomarker model (purple) or a dual biomarker model combining mucosal IgA and serum IgG (blue). (D) Two-dimensional protection surface for the dual biomarker model using serum IgG and mucosal IgA antibodies to RSV-A PreF. The continuous surface represents the predicted probability of protection given exposure as a function of both serum (x-axis) and mucosal (y-axis) antibody titres. Black contour lines delineate regions of constant protection probability at 50%, 70%, 80%, and 90%. Individual data points overlay the surface, with filled circles representing participants who were protected despite exposure and open circles representing those who became infected. Antibody titres are displayed on a log10 scale. (E) Pearson correlation coefficients between RSV-A variant biomarkers at the time of infection. Color intensity represents correlation strength, ranging from blue (weak correlation) through white (no correlation) to green (strong positive correlation).

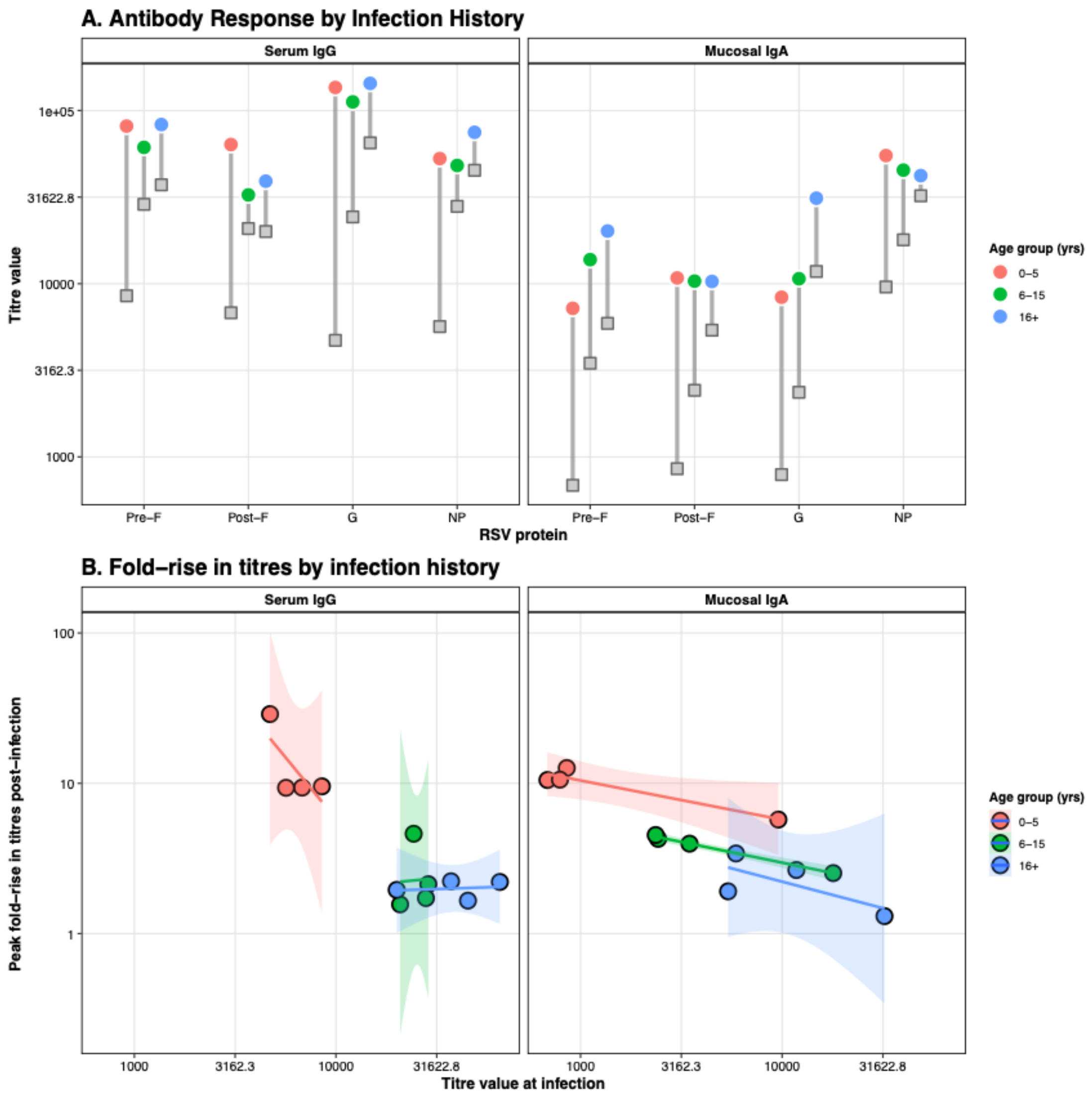


**Supplementary Results Figure 2.11. Age-dependent patterns in antibody magnitude and boosting following natural RSV infection. . $\rho = 0.9$** (A) Baseline and peak antibody titres stratified by age group for serum IgG (left panel) and mucosal IgA (right panel) against four RSV-A antigens (PreF, PostF, G, NP). Gray squares represent baseline titres at the time of infection, colored circles represent peak titres post-infection, and gray lines connecting them indicate the magnitude of antibody boosting. Point are colored according to age groups. (B) Relationship between baseline antibody titre at infection (x-axis) and fold-rise in titre post-infection (y-axis), illustrating the antibody ceiling effect where individuals with higher pre-existing titres exhibit smaller relative boosts. Linear regression lines (solid) with 95% confidence intervals (shaded regions) demonstrate the negative relationship.

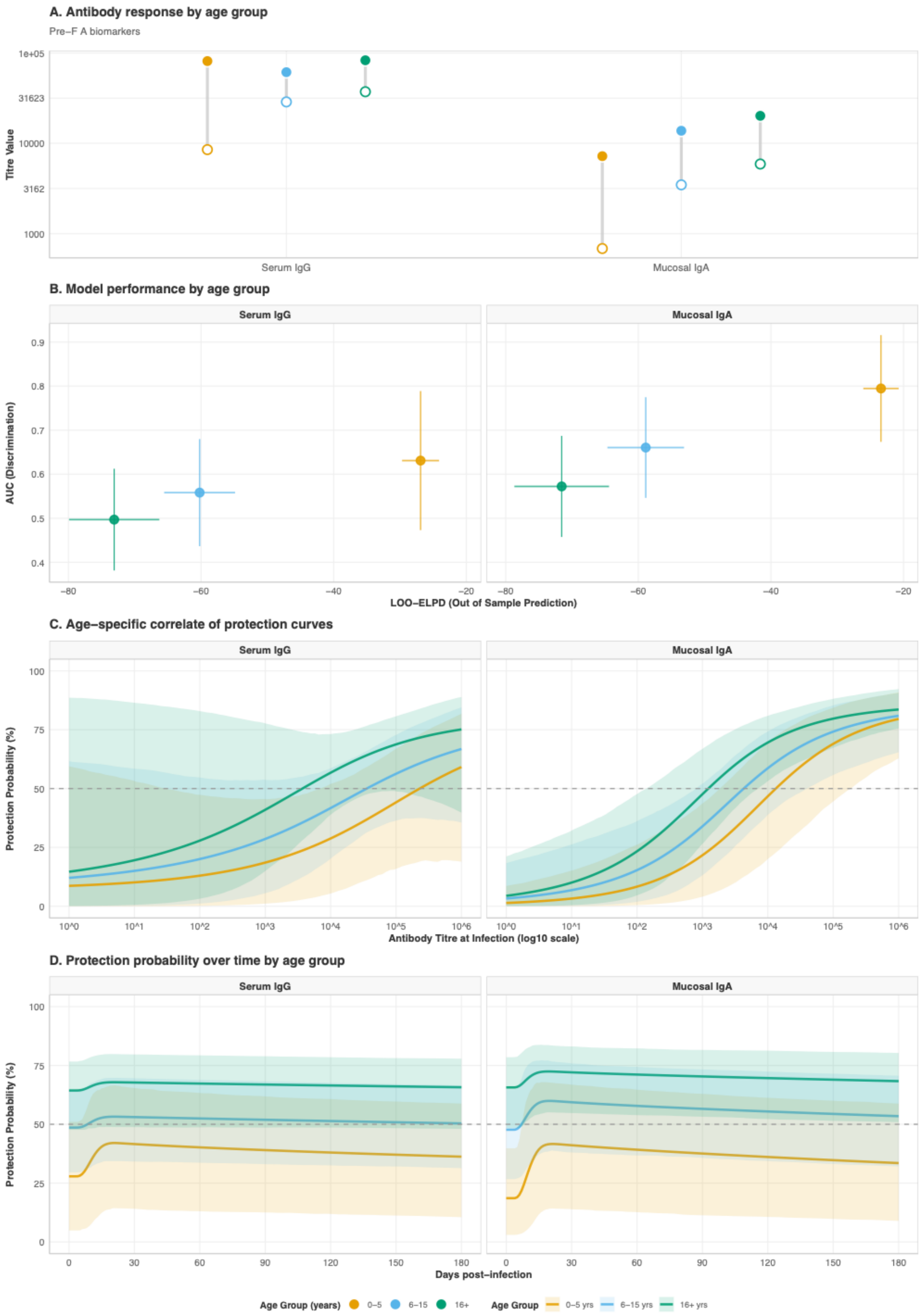


**Supplementary Results Figure 2.12. Age-stratified analysis of antibody responses and protective immunity for RSV-A PreF biomarkers. $\rho = 0.9$ (** A) Antibody response

magnitudes by age group, with baseline antibody titres at time of infection (hollow circles) and peak titres following infection (filled circles) for serum IgG and mucosal IgA antibodies to RSV-A PreF protein, stratified by age group (0-5 years in orange, 6-15 years in light blue, 16+ years in green). (B) Model performance by age group. The scatter plot compares out-of-sample predictive accuracy (LOO-ELPD, x-axis) and discrimination ability (AUC, y-axis) for correlate of protection models fitted separately within each age stratum.. Vertical error bars show 95% confidence intervals for AUC; horizontal error bars indicate standard error of LOO-ELPD. (C). Age-specific correlate of protection curves demonstrating the relationship between antibody titre at infection and probability of protection. (D) Protection probability trajectories by age group following infection. The lines show the predicted probability of protection against reinfection over time (0-180 days post-infection) for serum IgG (left panel) and mucosal IgA (right panel) antibodies to RSV-A PreF. Solid lines represent the median posterior prediction for each age group, with shaded ribbons indicating 95% credible intervals.

### 3. PCR + SERO MODEL $\rho = 0.8$

#### A. CONVERGENCE AND MODEL FITS

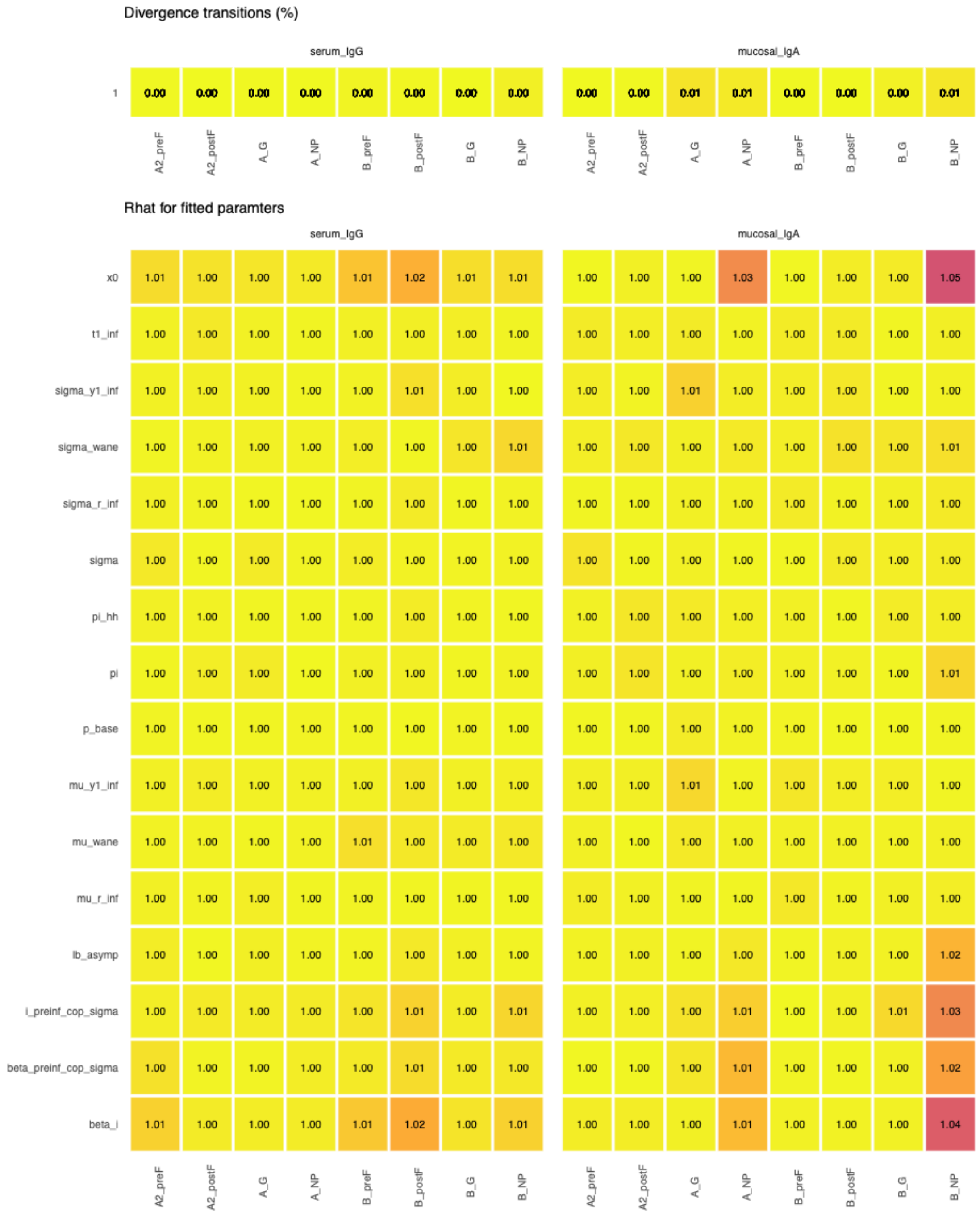


**Supplementary Results Figure 3.1. MCMC convergence diagnostics for 16 single biomarker antibody kinetics and correlates of protection models. $\rho = 0.8$.** Convergence diagnostics for Bayesian hierarchical models estimating antibody kinetics and correlates of protection. The top panel shows the percentage of divergent transitions

(out of 4,000 total iterations across 4 chains) for each biomarker-protein combination. All models achieved divergent transition rates < 2%, indicating successful exploration of the posterior distribution. Parameters shown include: mu_wane and sigma_wane (waning kinetics), mu_y1_inf and sigma_y1_inf (peak fold-rise), mu_r_inf and sigma_r_inf (boosting rate), t1_inf (time to peak), sigma (measurement error), lb_asymp (asymptotic lower bound), x0 (protection threshold), i_preinf_cop_sigma (pre-infection titre variability effect), beta_i (infection effect on susceptibility), beta_preinf_cop_sigma (pre-infection titre effect on protection), pi (within-household transmission probability), pi_hh (baseline attack rate), and p_base (baseline infection probability).

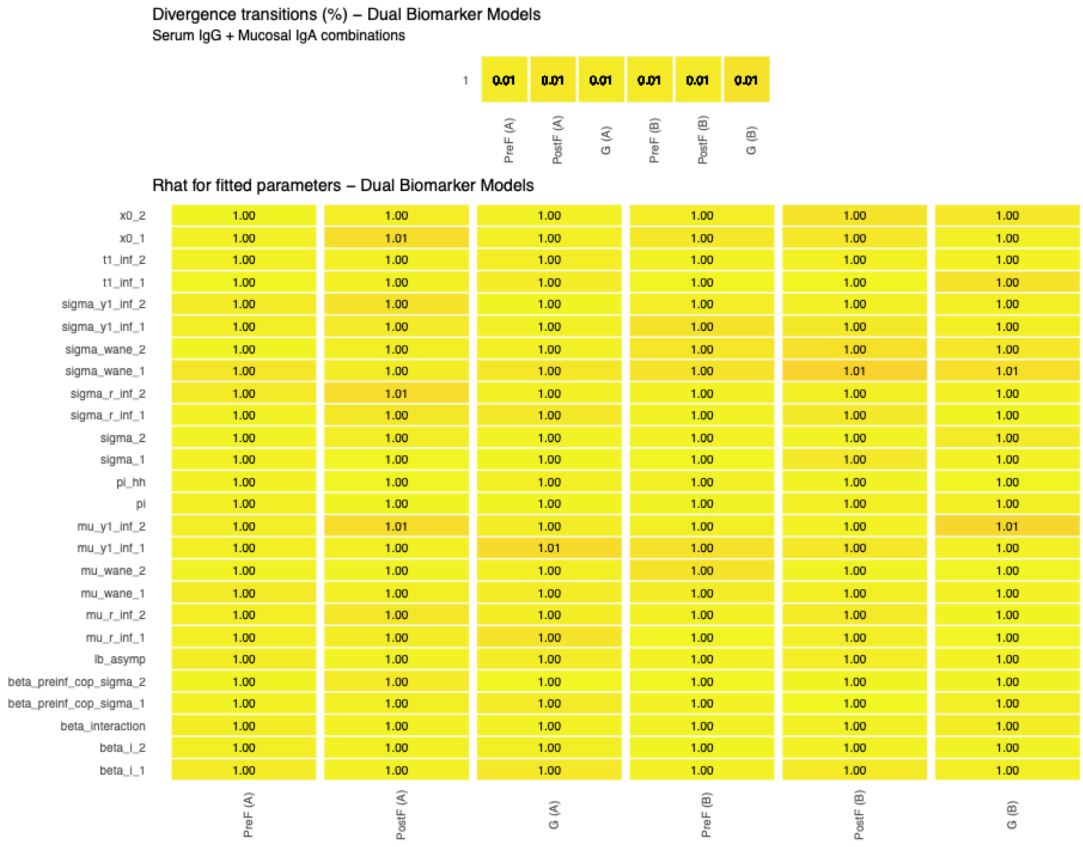


**Supplementary Results Figure 3.2. MCMC convergence diagnostics for 6 dual biomarker antibody kinetics and correlates of protection models. $\rho = 0.8$.** Convergence diagnostics for dual biomarker Bayesian hierarchical models simultaneously estimating antibody kinetics and correlates of protection for serum IgG and mucosal IgA targeting the same viral protein. The top panel shows the percentage of divergent transitions (out of 4,000 total iterations across 4 chains) for each dual biomarker model. All models achieved divergent transition rates < 1%, with most < 0.5%. Parameters shown include biomarker-specific parameters (with subscripts _1 and _2 denoting serum IgG and mucosal IgA, respectively): waning kinetics (mu_wane, sigma_wane), boosting kinetics (mu_y1_inf, sigma_y1_inf, mu_r_inf, sigma_r_inf, t1_inf), measurement error (sigma), protection thresholds (x0), and correlates of protection effects (beta_i,

beta_preinf_cop_sigma), as well as the interaction term (beta_interaction) capturing synergistic or antagonistic effects between the two biomarkers. Shared parameters include lb_asymp (asymptotic lower bound), pi (within-household transmission), and pi_hh (baseline attack rate).

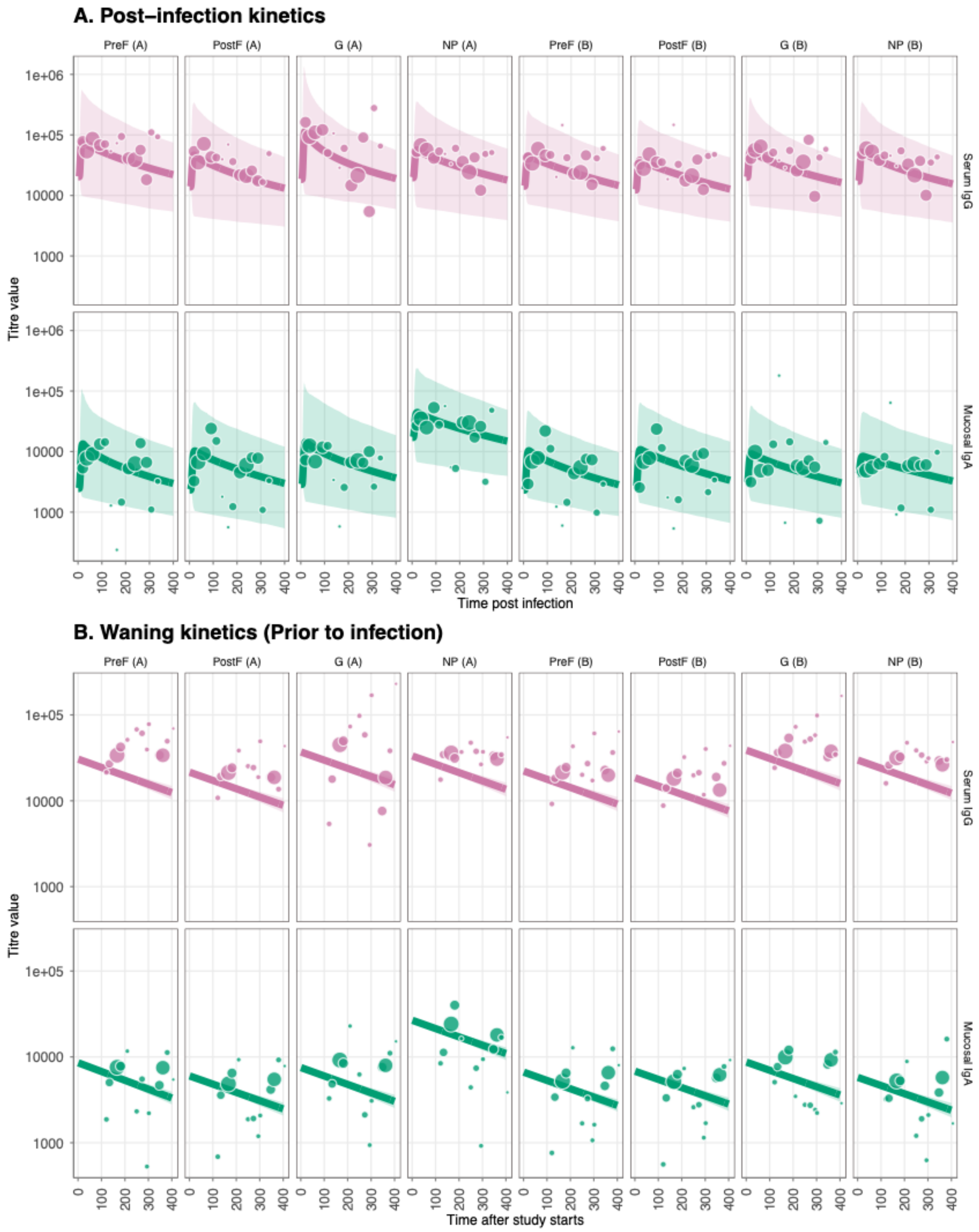


**Supplementary Results Figure 3.3. Antibody kinetics fits and immune responses for RSV-A and RSV-B post and prior to infection. $\rho = 0.8$.** (A) Post-infection longitudinal antibody titers for eight viral RSV-A/B proteins (PreF, PostF, G, and NP) across different

antibody types; serum IgG and mucosal IgA. Lines show the median posterior predictive fit from the fitted Bayesian model, and the points show the observational titre data, with the size correlating with the sample size for that bin. (B) Prior-infection antibody waning for eight viral RSV-A/B proteins (PreF, PostF, G, and NP) across different antibody types; serum IgG and mucosal IgA.

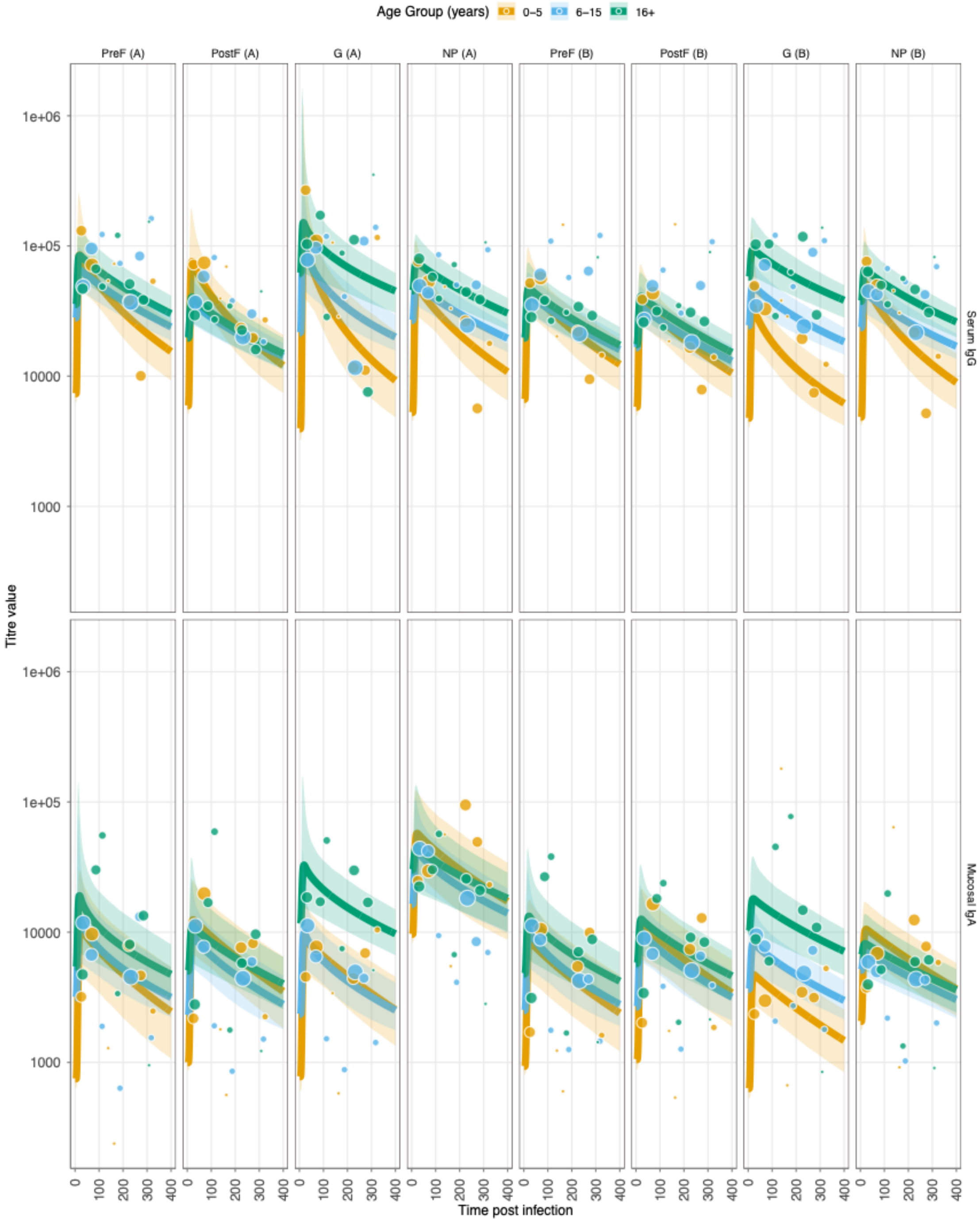

**Supplementary Results Figure 3.4. Age-stratified antibody kinetics fits and immune responses for RSV-A and RSV-B post and prior to infection. $\rho = 0.8$** Layout as **Supplementary Figure 3.3.**

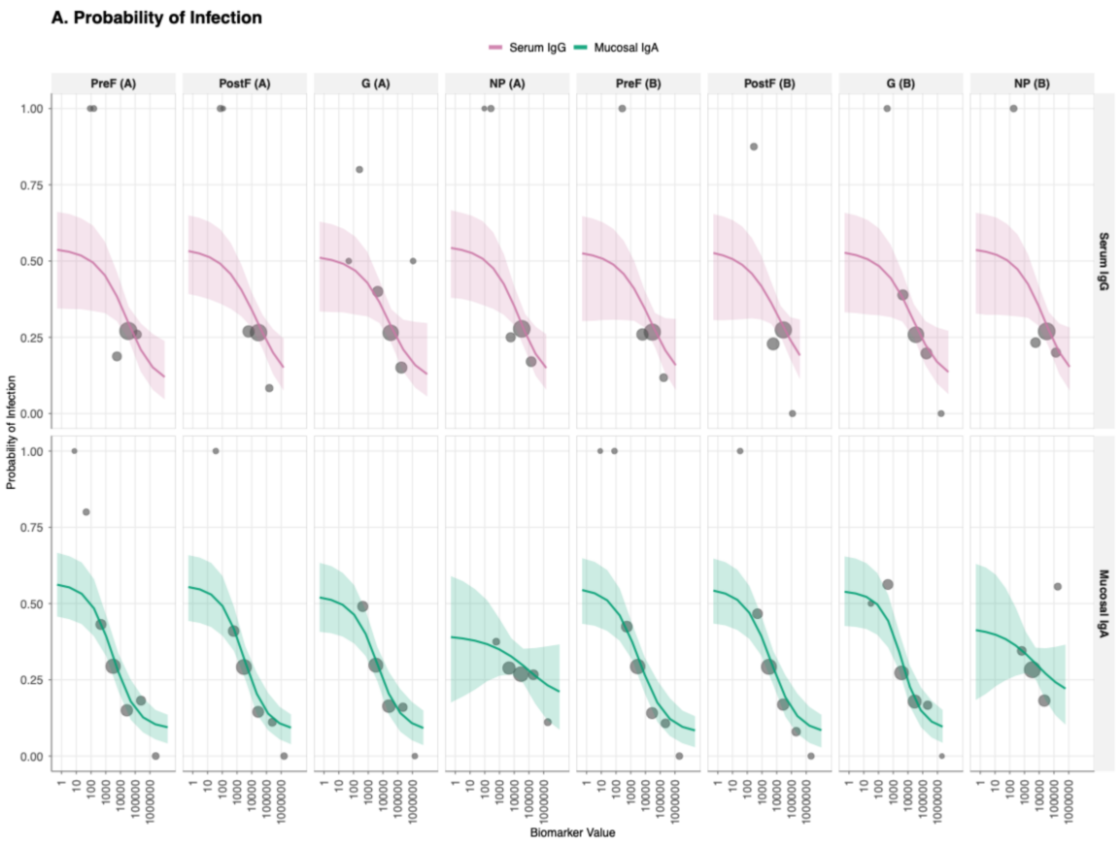


**Supplementary Results Figure 3.5. Correlate of risk (CoR) curves showing the relationship between antibody titre at infection and probability of protection from RSV infection for all 16 biomarker-antigen combinations. $\rho = 0.8$.** Serum IgG, top row; mucosal IgA, bottom row and columns are viral antigen target (PreF, PostF, G, and NP for both RSV-A and RSV-B strains). The solid lines represents the mean estimated probability of protection given exposure to infection as a function of antibody titre, with shaded ribbons indicating 95% credible intervals, and the points show the observational titre at infection, with the size correlating with the sample size for that bin.

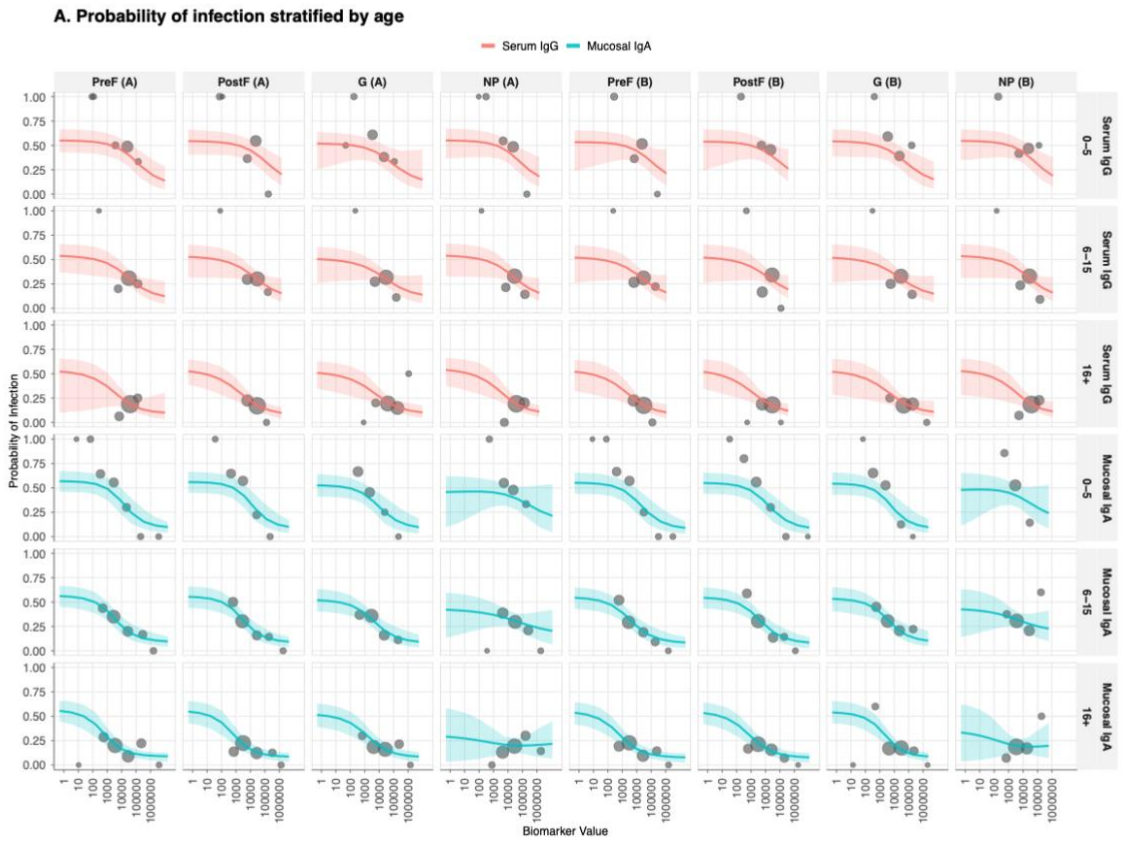


**Supplementary Results Figure 3.6. Age-stratified correlate of risk (CoR) curves showing the relationship between antibody titre at infection and probability of protection from RSV infection for all 16 biomarker-antigen combinations. $\rho = 0.8$.** Layout as **Supplementary Figure 3.5.**

#### B. RESULTS AND INFERENCE

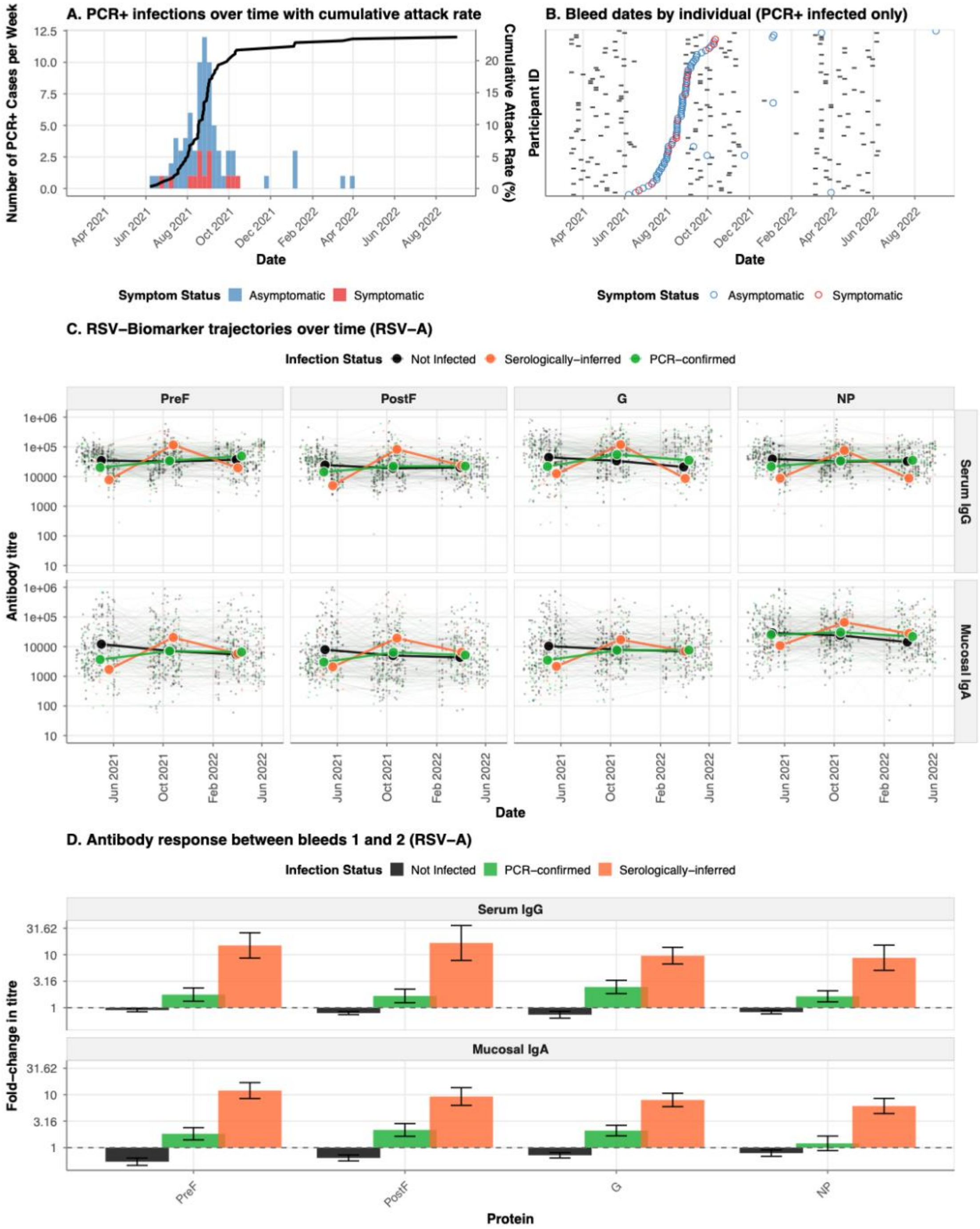


**Supplementary Results Figure 3.7. Epidemiology of RSV infections and antibody kinetics during the 2022-2023 epidemic season. $\rho = 0.8$ A.** Weekly incidence of PCR-confirmed RSV infections (stacked histogram) and cumulative attack rate (black line) among 343 household cohort participants from October 2022 to May 2023. Bars are colored by symptom status at detection: symptomatic (red, detected at unscheduled

visits) or asymptomatic (blue, detected at scheduled home or clinic visits). B. Timeline of serological sampling for PCR-confirmed infected individuals. Each horizontal line represents one participant (ordered by date of PCR-confirmed infection), with grey tiles indicating bleed dates and colored circles marking the timing of PCR-confirmed infection (colored by symptom status as in panel a). **C.** Longitudinal trajectories of serum IgG (top row) and mucosal IgA (bottom row) antibody responses against four RSV-A proteins (PreF, PostF, G, NP) over time. Individual participant trajectories are shown as thin lines with low opacity, colored by infection status: not infected (black), serologically-defined infections (orange), and PCR-confirmed infections (green). **D.** Mean fold-change in antibody titres between the first bleed (pre-epidemic baseline) and second bleed (post-epidemic) for serum IgG and mucosal IgA responses to RSV-A proteins. Bars represent mean fold-change (log10 scale) stratified by infection status, with error bars indicating standard error.

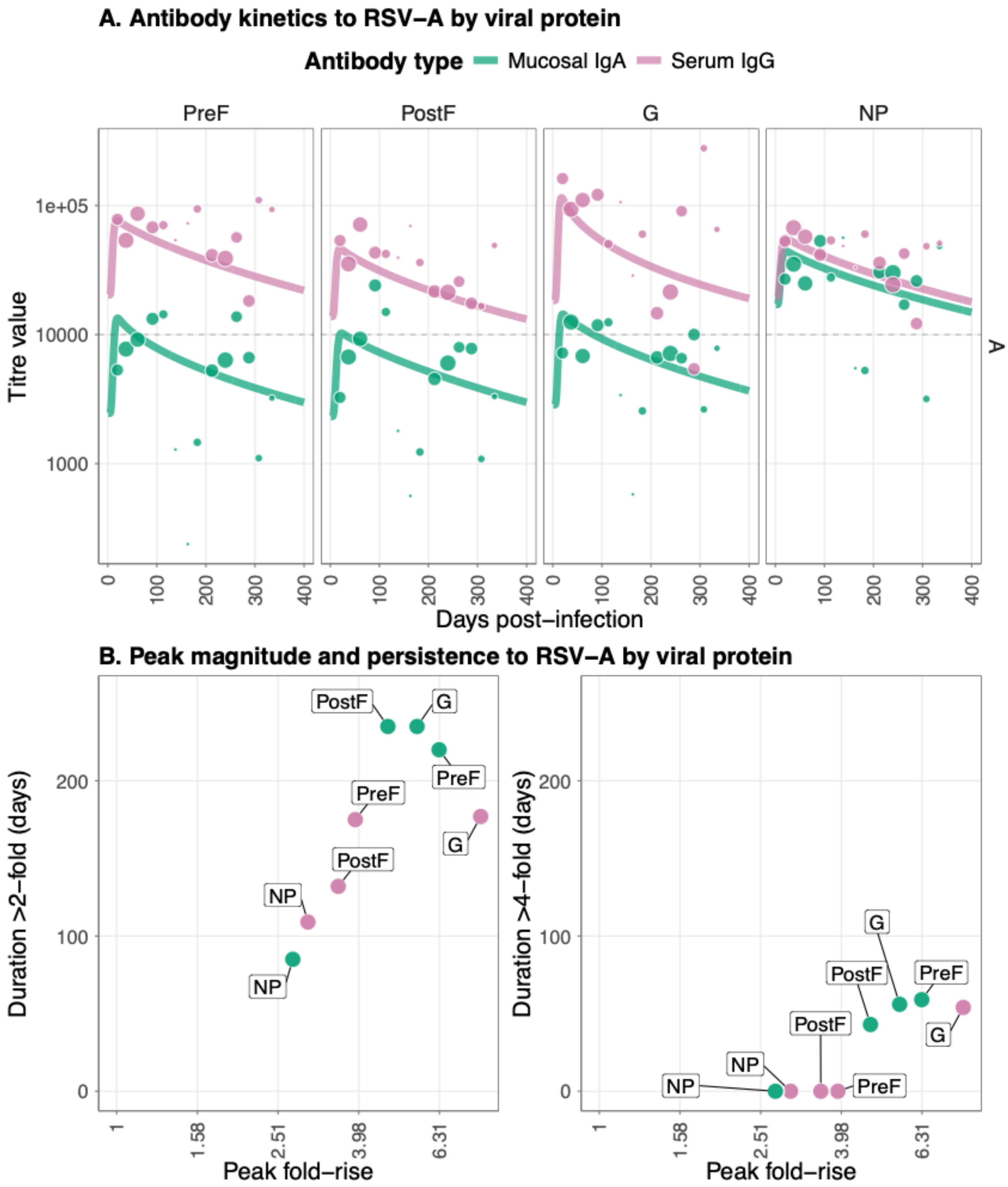


**Supplementary Results Figure 3.8. Antibody kinetics and immune responses for infections to RSV-A with $\rho = 0.8$** (A) Post-infection longitudinal antibody titers for four viral RSV-A proteins (PreF, PostF, G, and NP) across different antibody types; serum IgG and mucosal IgA. Lines show the median posterior predictive fit from the fitted Bayesian model, and the points show the observational titre data, with the size correlating with the sample size for that bin. (B) Peak antibody (x axis) and persistence measured as duration above a 2-fold (left panel) and 4-fold titre rise (right panel) in days. Data points show median posterior values for measurements of four viral RSV-A proteins PreF, PostF, G, and NP across different antibody types: serum IgG and mucosal IgA.

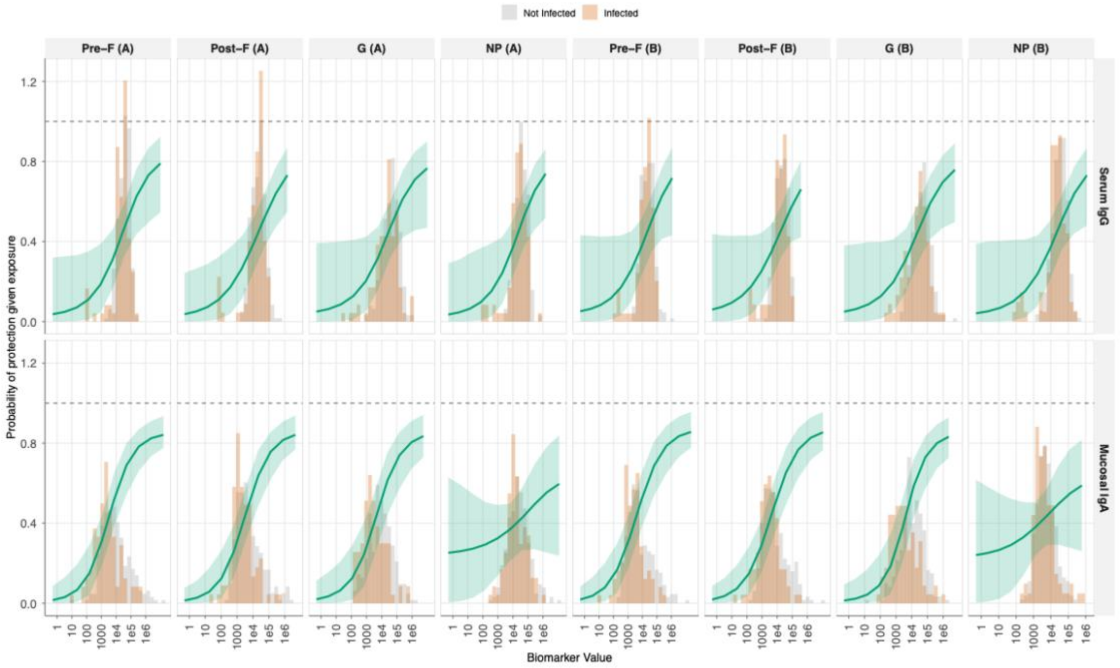


**Supplementary Results Figure 3.9. Correlate of protection (CoP) curves showing the relationship between antibody titre at infection and probability of protection from RSV infection for all 16 biomarker-antigen combinations. .** $\boldsymbol{\rho = 0.8}$ Serum IgG, top row; mucosal IgA, bottom row and columns are viral antigen target (PreF, PostF, G, and NP for both RSV-A and RSV-B strains). The solid green line represents the mean estimated probability of protection given exposure to infection as a function of antibody titre, with shaded ribbons indicating 95% credible intervals. Background histograms show the distribution of antibody titres at infection for infected individuals (orange) versus non-infected individuals (gray).

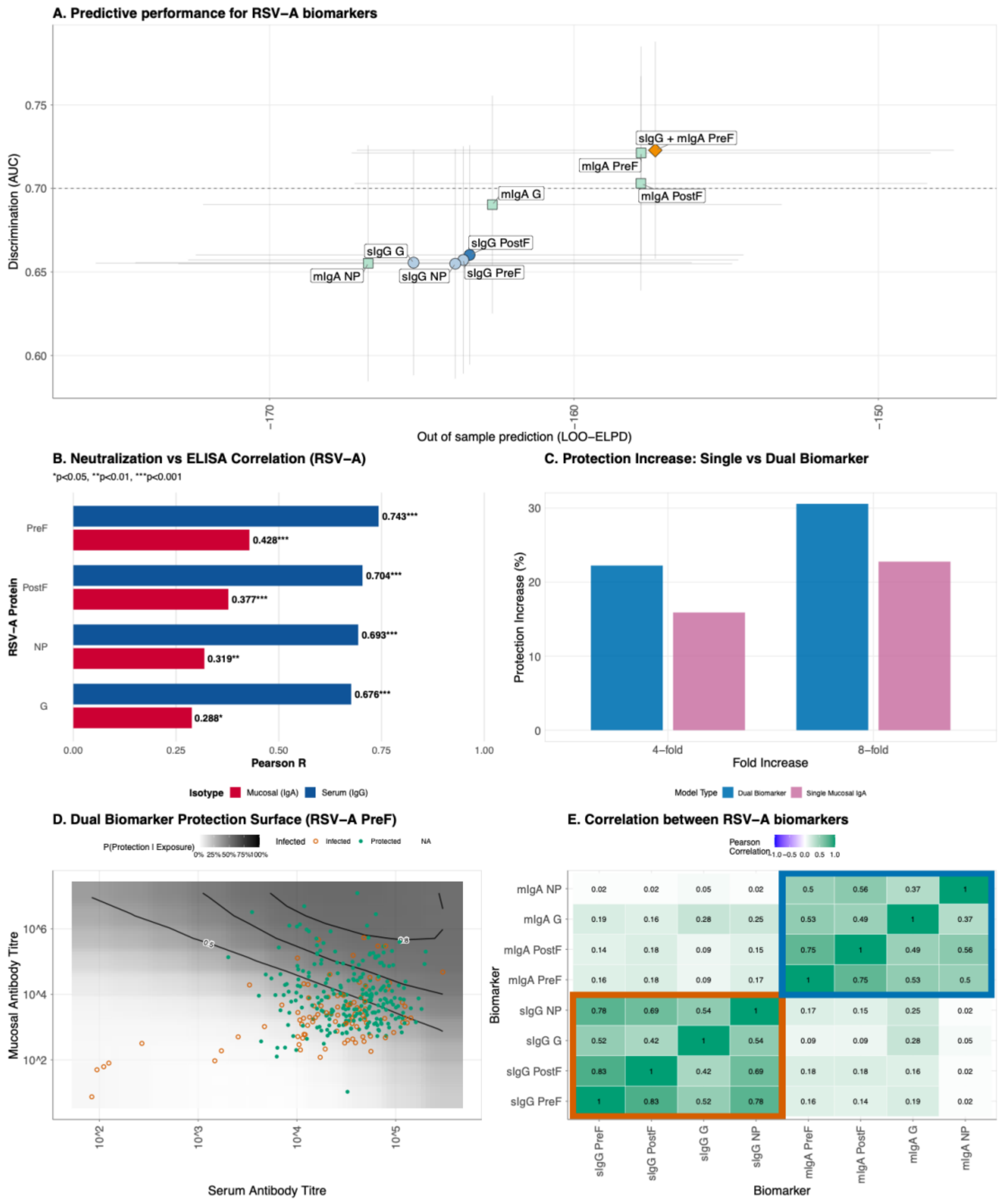


**Supplementary Results Figure 3.10. Comparison of single and dual biomarker models for predicting protection against RSV-A . $\rho = 0.8$ infection.** (A) Model performance comparison across single biomarker models and the dual biomarker model, defined by out-of-sample predictive accuracy (LOO-ELPD, x-axis) and discrimination ability (area under the ROC curve, AUC, y-axis). Circles indicate serum IgG models, squares represent mucosal IgA models, and the triangle denotes the dual biomarker model combining serum IgG and mucosal IgA to RSV-A PreF. The best-performing model within each biomarker class is highlighted with darker shading. Error bars show the standard error of LOO-ELPD (horizontal) and 95% confidence intervals for AUC (vertical). The dashed horizontal line

indicates an AUC of 0.7. (B) Summary of Pearson correlation coefficients for RSV-A protein antigens, comparing serum IgG (blue) and mucosal IgA (red) ELISA titres with neutralisation IC50 values. (C) Counterfactual analysis comparing the increase in protection from boosting pre-infection antibody titres by 4-fold or 8-fold using either a single mucosal IgA biomarker model (purple) or a dual biomarker model combining mucosal IgA and serum IgG (blue). (D) Two-dimensional protection surface for the dual biomarker model using serum IgG and mucosal IgA antibodies to RSV-A PreF. The continuous surface represents the predicted probability of protection given exposure as a function of both serum (x-axis) and mucosal (y-axis) antibody titres. Black contour lines delineate regions of constant protection probability at 50%, 70%, 80%, and 90%. Individual data points overlay the surface, with filled circles representing participants who were protected despite exposure and open circles representing those who became infected. Antibody titres are displayed on a log10 scale. (E) Pearson correlation coefficients between RSV-A variant biomarkers at the time of infection. Color intensity represents correlation strength, ranging from blue (weak correlation) through white (no correlation) to green (strong positive correlation).

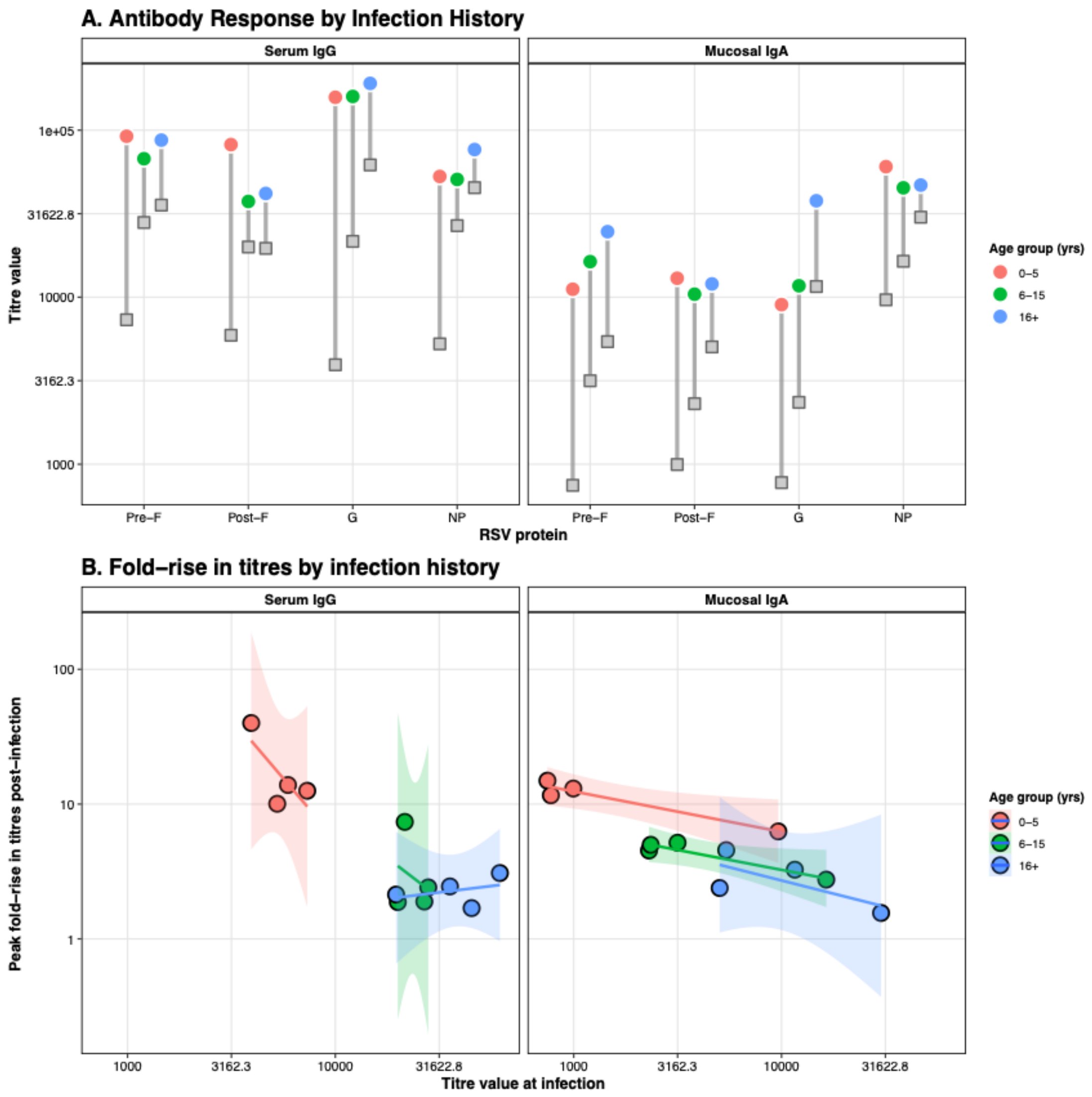


**Supplementary Results Figure 3.11. Age-dependent patterns in antibody magnitude and boosting following natural RSV infection. . $\rho = 0.8$** (A) Baseline and peak antibody titres stratified by age group for serum IgG (left panel) and mucosal IgA (right panel) against four RSV-A antigens (PreF, PostF, G, NP). Gray squares represent baseline titres at the time of infection, colored circles represent peak titres post-infection, and gray lines connecting them indicate the magnitude of antibody boosting. Point are colored according to age groups. (B) Relationship between baseline antibody titre at infection (x-axis) and fold-rise in titre post-infection (y-axis), illustrating the antibody ceiling effect where individuals with higher pre-existing titres exhibit smaller relative boosts. Linear regression lines (solid) with 95% confidence intervals (shaded regions) demonstrate the negative relationship.

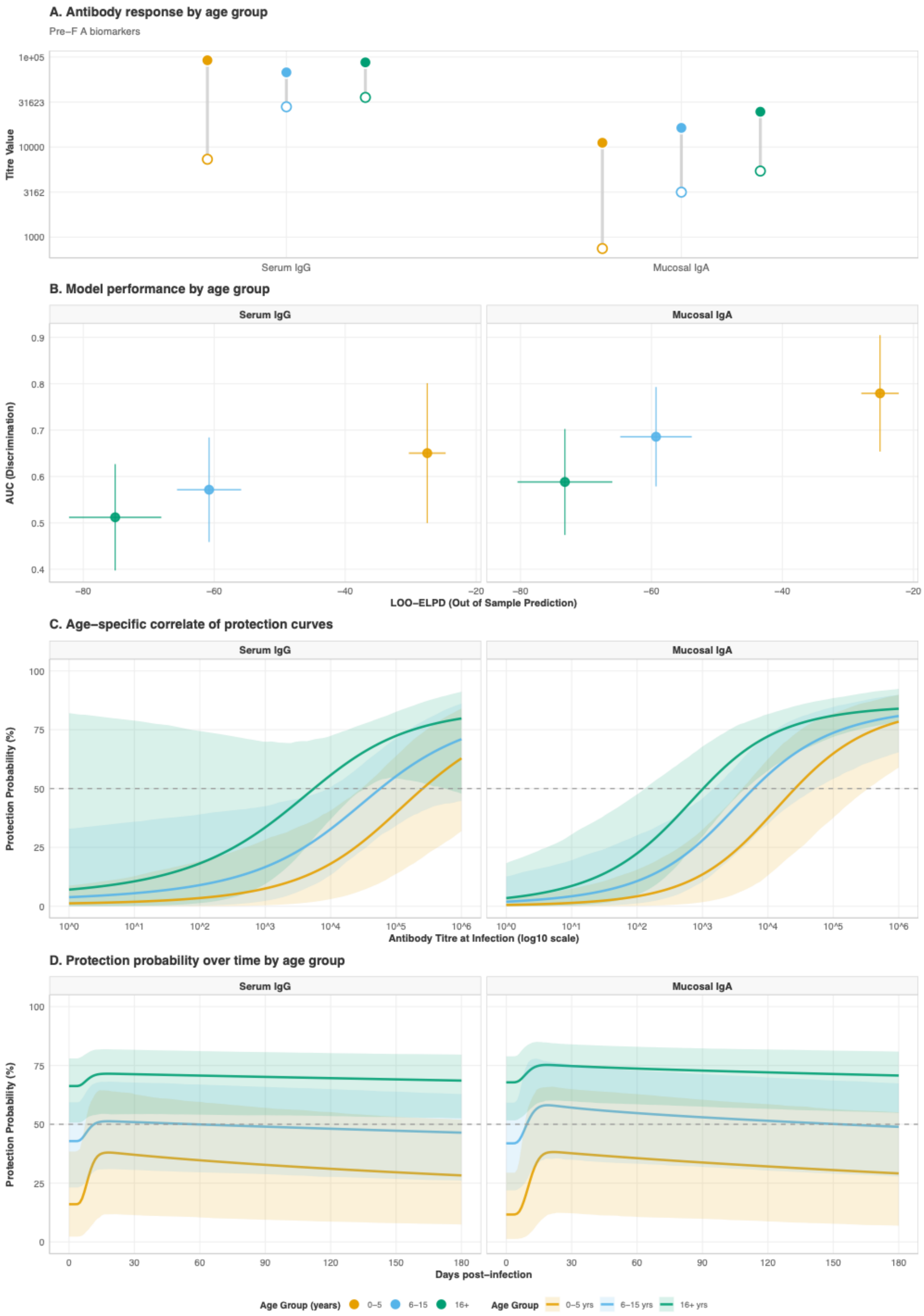


**Supplementary Results Figure 3.12. Age-stratified analysis of antibody responses and protective immunity for RSV-A PreF biomarkers. $\rho = 0.8$ (** A) Antibody response

magnitudes by age group, with baseline antibody titres at time of infection (hollow circles) and peak titres following infection (filled circles) for serum IgG and mucosal IgA antibodies to RSV-A PreF protein, stratified by age group (0-5 years in orange, 6-15 years in light blue, 16+ years in green). (B) Model performance by age group. The scatter plot compares out-of-sample predictive accuracy (LOO-ELPD, x-axis) and discrimination ability (AUC, y-axis) for correlate of protection models fitted separately within each age stratum.. Vertical error bars show 95% confidence intervals for AUC; horizontal error bars indicate standard error of LOO-ELPD. (C). Age-specific correlate of protection curves demonstrating the relationship between antibody titre at infection and probability of protection. (D) Protection probability trajectories by age group following infection. The lines show the predicted probability of protection against reinfection over time (0-180 days post-infection) for serum IgG (left panel) and mucosal IgA (right panel) antibodies to RSV-A PreF. Solid lines represent the median posterior prediction for each age group, with shaded ribbons indicating 95% credible intervals.

---

### 4. **PCR + SERO MODEL $\rho = 0.7$**

#### A. CONVERGENCE AND MODEL FITS

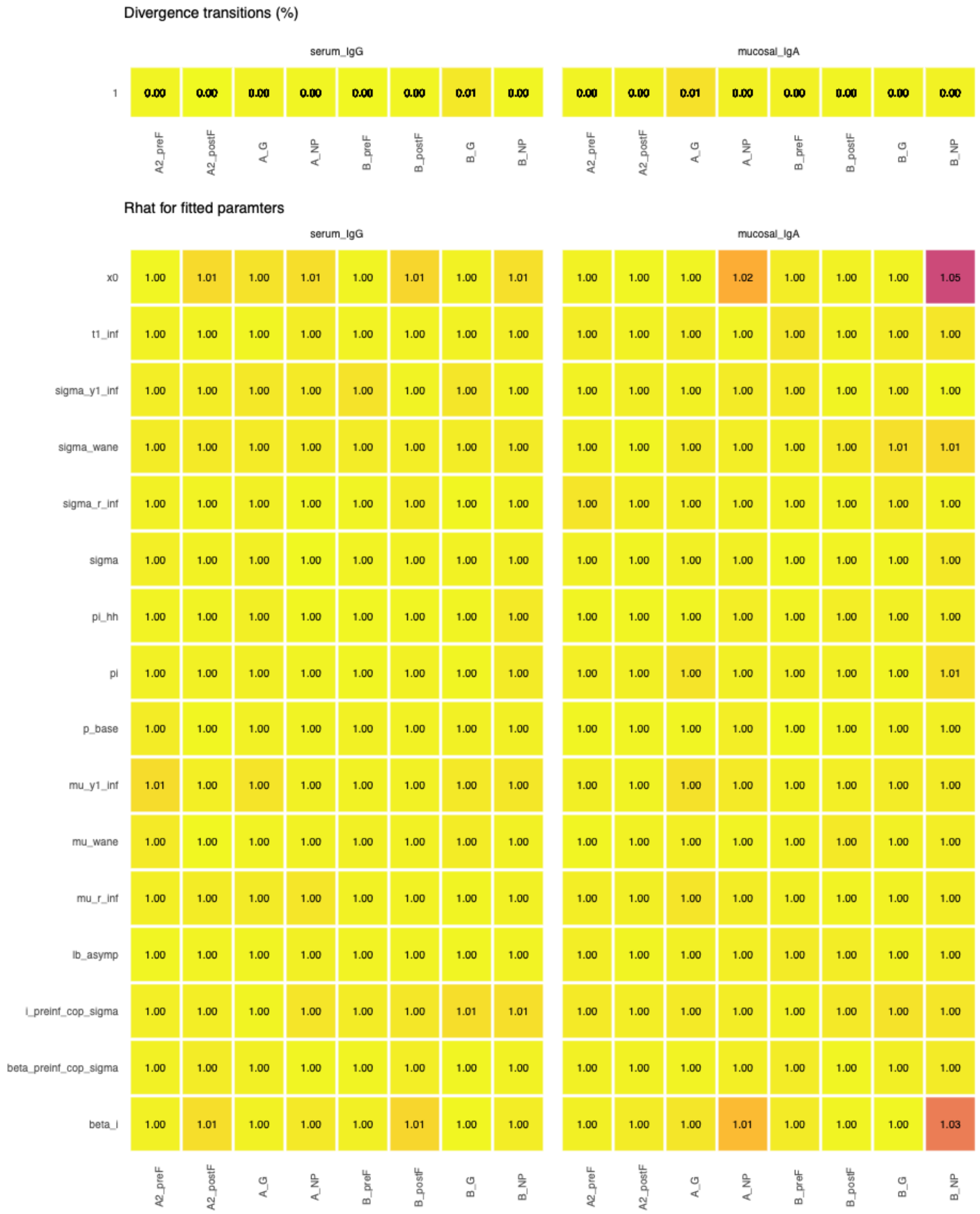


**Supplementary Results Figure 4.1. MCMC convergence diagnostics for 16 single biomarker antibody kinetics and correlates of protection models. $\rho = 0.7$.** Convergence diagnostics for Bayesian hierarchical models estimating antibody kinetics and correlates of protection. The top panel shows the percentage of divergent transitions

(out of 4,000 total iterations across 4 chains) for each biomarker-protein combination. All models achieved divergent transition rates < 2%, indicating successful exploration of the posterior distribution. Parameters shown include: mu_wane and sigma_wane (waning kinetics), mu_y1_inf and sigma_y1_inf (peak fold-rise), mu_r_inf and sigma_r_inf (boosting rate), t1_inf (time to peak), sigma (measurement error), lb_asymp (asymptotic lower bound), x0 (protection threshold), i_preinf_cop_sigma (pre-infection titre variability effect), beta_i (infection effect on susceptibility), beta_preinf_cop_sigma (pre-infection titre effect on protection), pi (within-household transmission probability), pi_hh (baseline attack rate), and p_base (baseline infection probability).

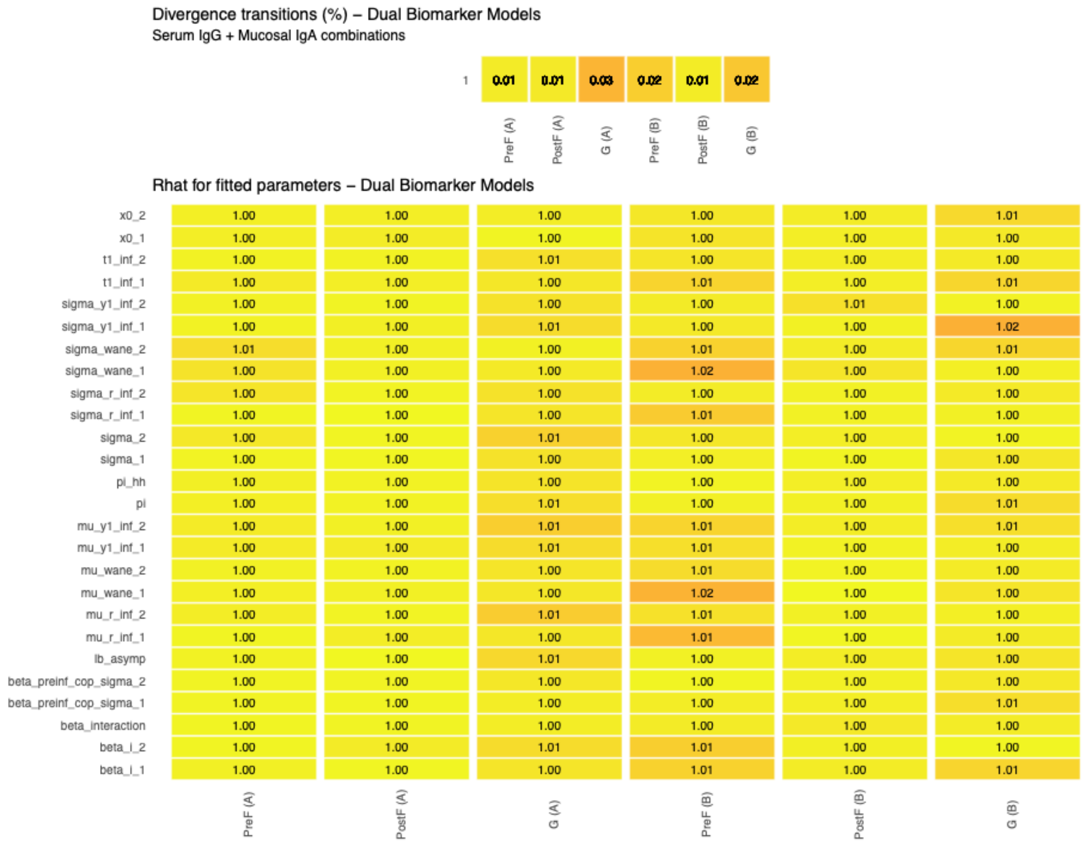


**Supplementary Results Figure 4.2. MCMC convergence diagnostics for 6 dual biomarker antibody kinetics and correlates of protection models. $\rho = 0.7$.** Convergence diagnostics for dual biomarker Bayesian hierarchical models simultaneously estimating antibody kinetics and correlates of protection for serum IgG and mucosal IgA targeting the same viral protein. The top panel shows the percentage of divergent transitions (out of 4,000 total iterations across 4 chains) for each dual biomarker model. All models achieved divergent transition rates < 1%, with most < 0.5%. Parameters shown include biomarker-specific parameters (with subscripts _1 and _2 denoting serum IgG and mucosal IgA, respectively): waning kinetics (mu_wane, sigma_wane), boosting kinetics (mu_y1_inf, sigma_y1_inf, mu_r_inf, sigma_r_inf, t1_inf), measurement error (sigma), protection thresholds (x0), and correlates of protection effects (beta_i,

beta_preinf_cop_sigma), as well as the interaction term (beta_interaction) capturing synergistic or antagonistic effects between the two biomarkers. Shared parameters include lb_asymp (asymptotic lower bound), pi (within-household transmission), and pi_hh (baseline attack rate).

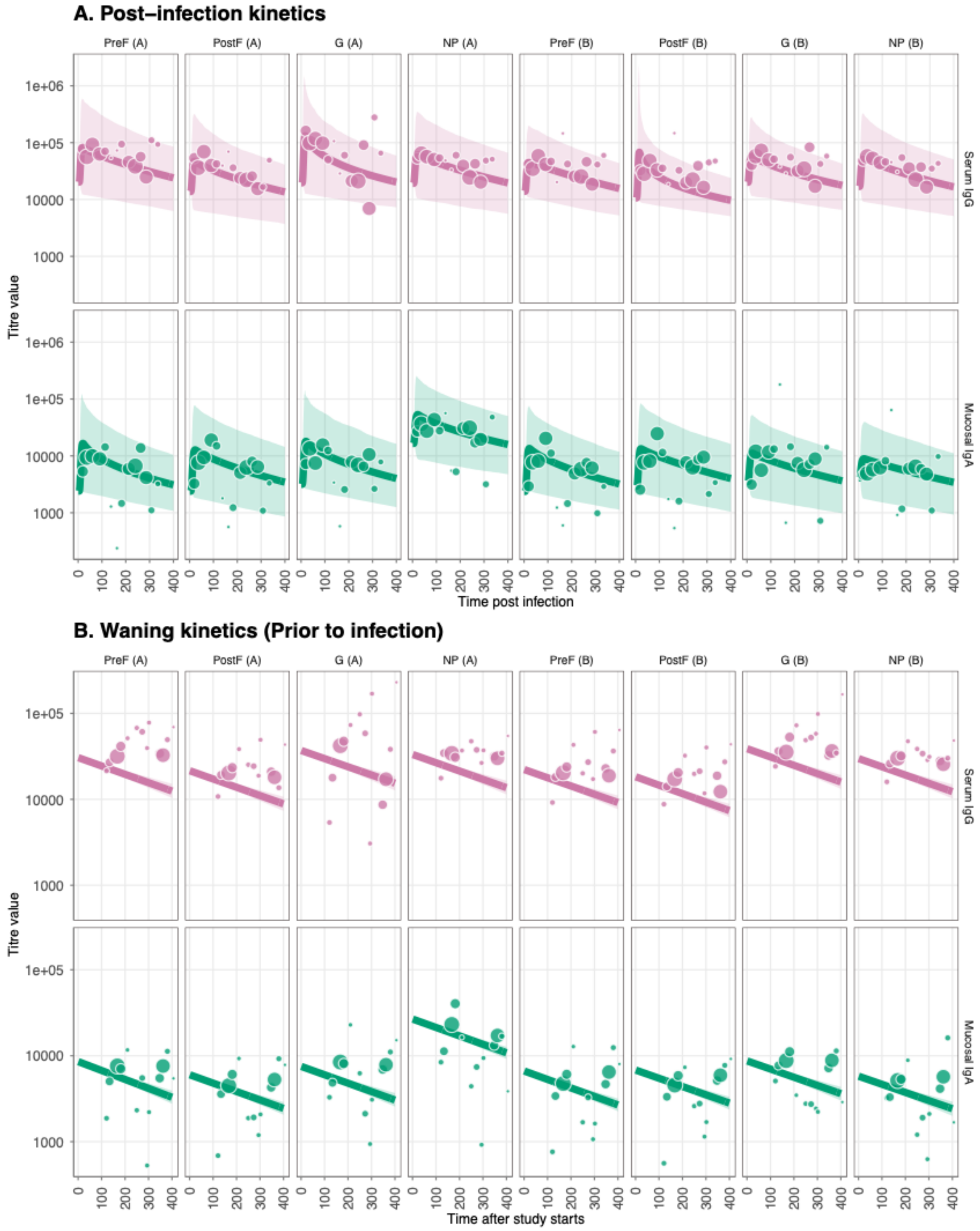


**Supplementary Results Figure 4.3. Antibody kinetics fits and immune responses for RSV-A and RSV-B post and prior to infection. $\rho = 0.7$.** (A) Post-infection longitudinal antibody titers for eight viral RSV-A/B proteins (PreF, PostF, G, and NP) across different

antibody types; serum IgG and mucosal IgA. Lines show the median posterior predictive fit from the fitted Bayesian model, and the points show the observational titre data, with the size correlating with the sample size for that bin. (B) Prior-infection antibody waning for eight viral RSV-A/B proteins (PreF, PostF, G, and NP) across different antibody types; serum IgG and mucosal IgA.

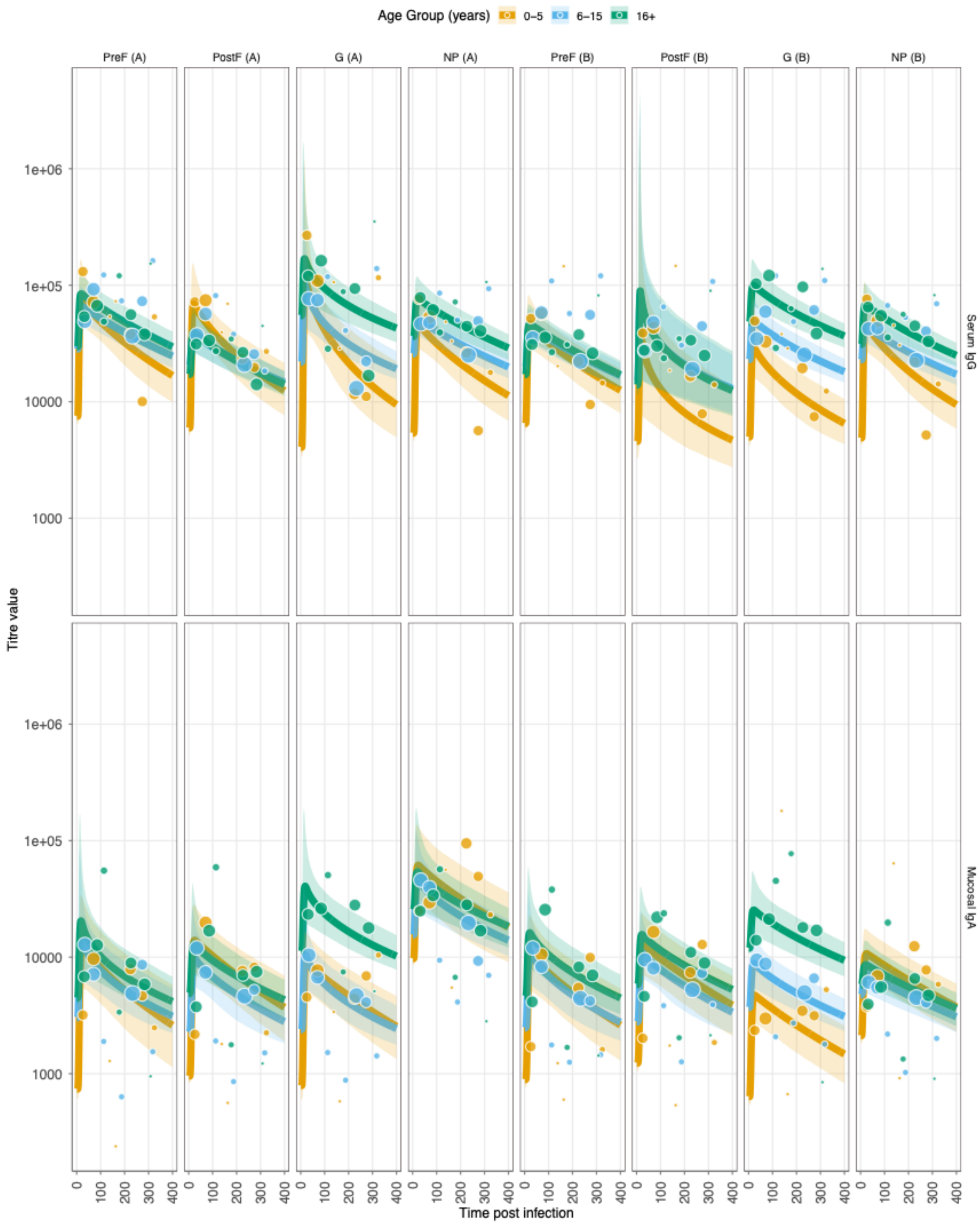

**Supplementary Results Figure 4.4. Age-stratified antibody kinetics fits and immune responses for RSV-A and RSV-B post and prior to infection. $\rho = 0.7$.** Layout as **Supplementary Figure 4.3.**

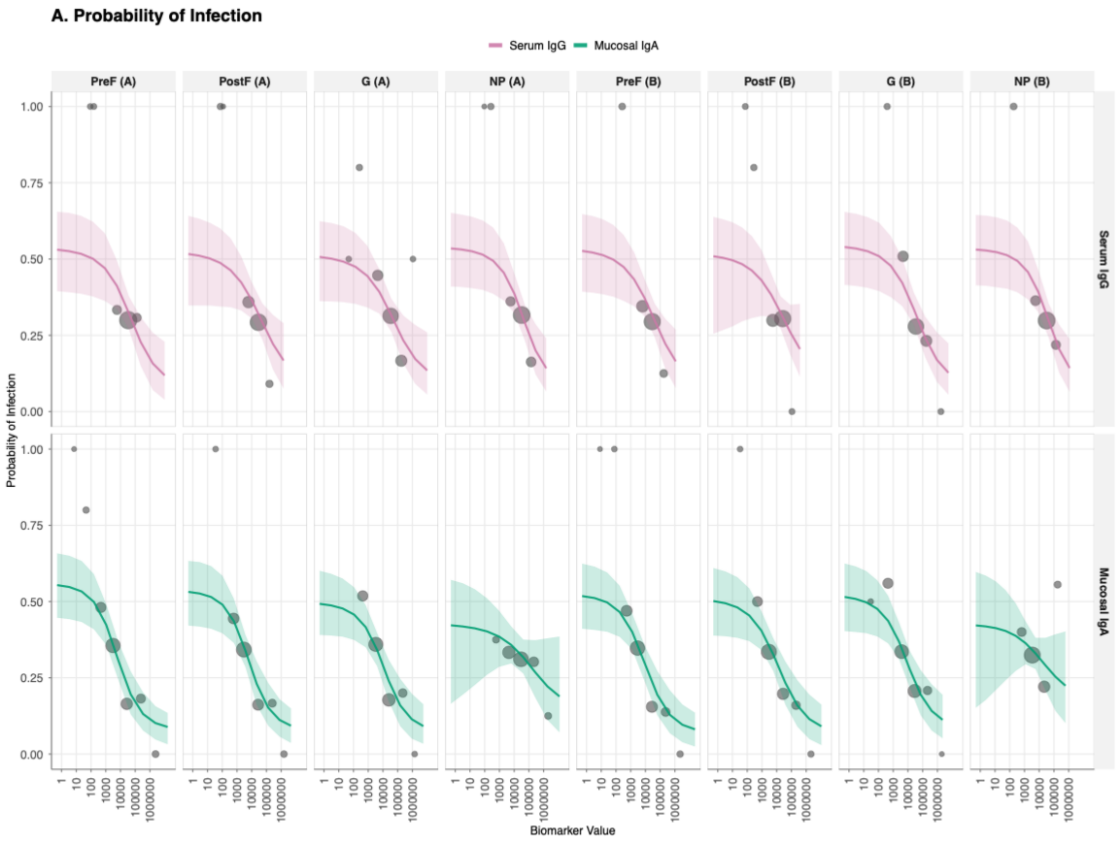


**Supplementary Results Figure 4.5. Correlate of risk (CoR) curves showing the relationship between antibody titre at infection and probability of protection from RSV infection for all 16 biomarker-antigen combinations. $\rho = 0.7$.** Serum IgG, top row; mucosal IgA, bottom row and columns are viral antigen target (PreF, PostF, G, and NP for both RSV-A and RSV-B strains). The solid lines represents the mean estimated probability of protection given exposure to infection as a function of antibody titre, with shaded ribbons indicating 95% credible intervals, and the points show the observational titre at infection, with the size correlating with the sample size for that bin.

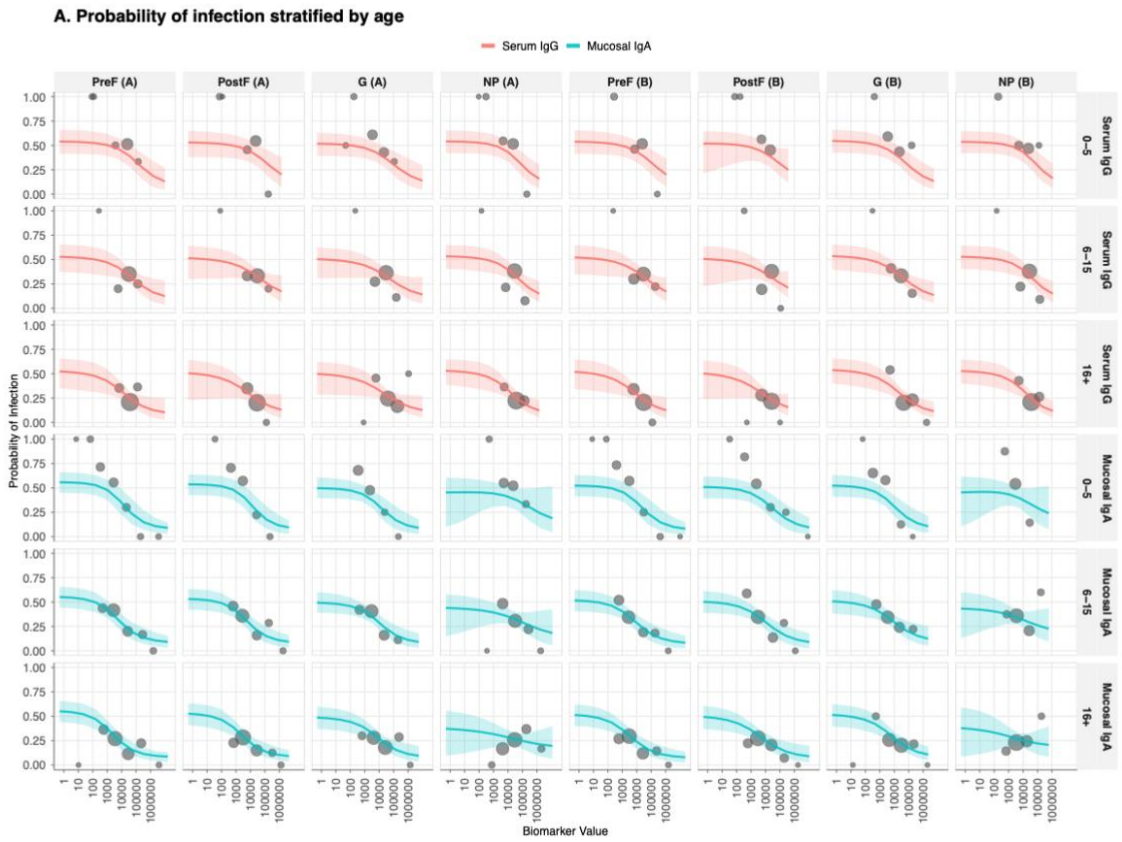


**Supplementary Results Figure 4.6. Age-stratified correlate of risk (CoR) curves showing the relationship between antibody titre at infection and probability of protection from RSV infection for all 16 biomarker-antigen combinations. $\rho = 0.7$.** Layout as **Supplementary Figure 4.5.**

#### B. RESULTS AND INFERENCE

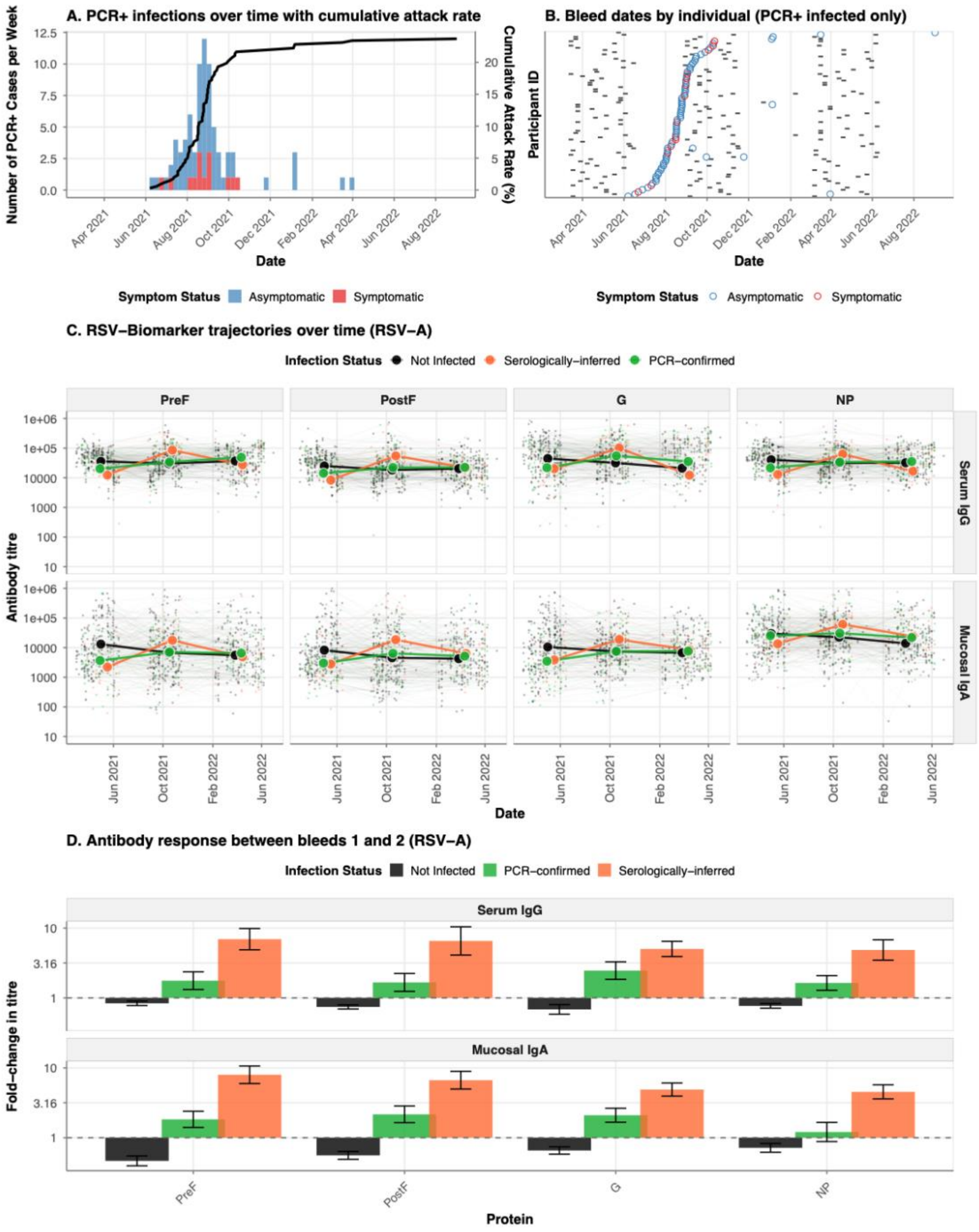


**Supplementary Results Figure 4.7. Epidemiology of RSV infections and antibody kinetics during the 2022-2023 epidemic season. $\rho = 0.7$ A.** Weekly incidence of PCR-confirmed RSV infections (stacked histogram) and cumulative attack rate (black line) among 343 household cohort participants from October 2022 to May 2023. Bars are colored by symptom status at detection: symptomatic (red, detected at unscheduled

visits) or asymptomatic (blue, detected at scheduled home or clinic visits). B. Timeline of serological sampling for PCR-confirmed infected individuals. Each horizontal line represents one participant (ordered by date of PCR-confirmed infection), with grey tiles indicating bleed dates and colored circles marking the timing of PCR-confirmed infection (colored by symptom status as in panel a). **C.** Longitudinal trajectories of serum IgG (top row) and mucosal IgA (bottom row) antibody responses against four RSV-A proteins (PreF, PostF, G, NP) over time. Individual participant trajectories are shown as thin lines with low opacity, colored by infection status: not infected (black), serologically-defined infections (orange), and PCR-confirmed infections (green). **D.** Mean fold-change in antibody titres between the first bleed (pre-epidemic baseline) and second bleed (post-epidemic) for serum IgG and mucosal IgA responses to RSV-A proteins. Bars represent mean fold-change (log10 scale) stratified by infection status, with error bars indicating standard error.

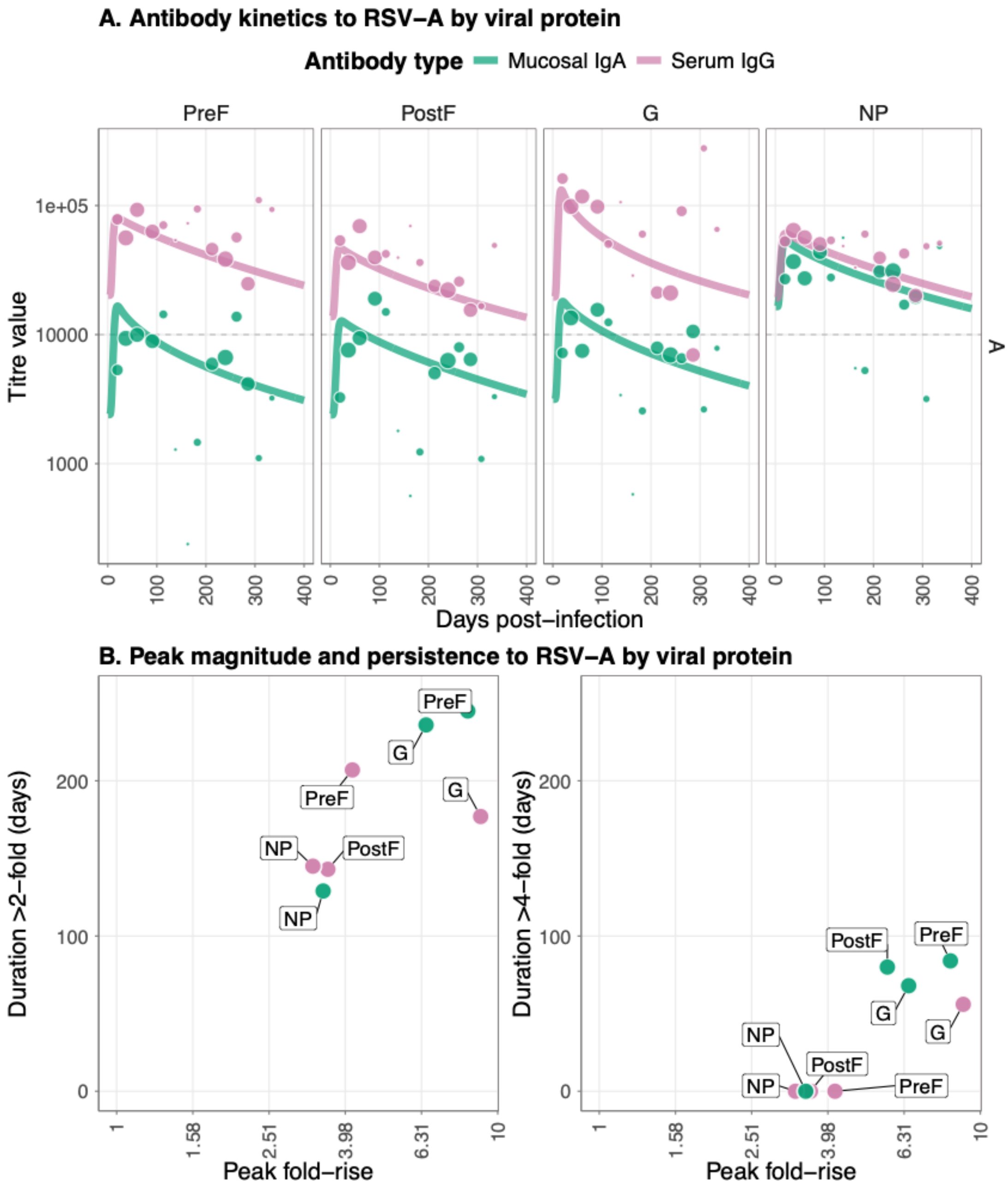


**Supplementary Results Figure 4.8. Antibody kinetics and immune responses for infections to RSV-A with $\rho = 0.7$** (A) Post-infection longitudinal antibody titers for four viral RSV-A proteins (PreF, PostF, G, and NP) across different antibody types; serum IgG and mucosal IgA. Lines show the median posterior predictive fit from the fitted Bayesian model, and the points show the observational titre data, with the size correlating with the sample size for that bin. (B) Peak antibody (x axis) and persistence measured as duration above a 2-fold (left panel) and 4-fold titre rise (right panel) in days. Data points show median posterior values for measurements of four viral RSV-A proteins (PreF, PostF, G, and NP) across different antibody types: serum IgG and mucosal IgA.

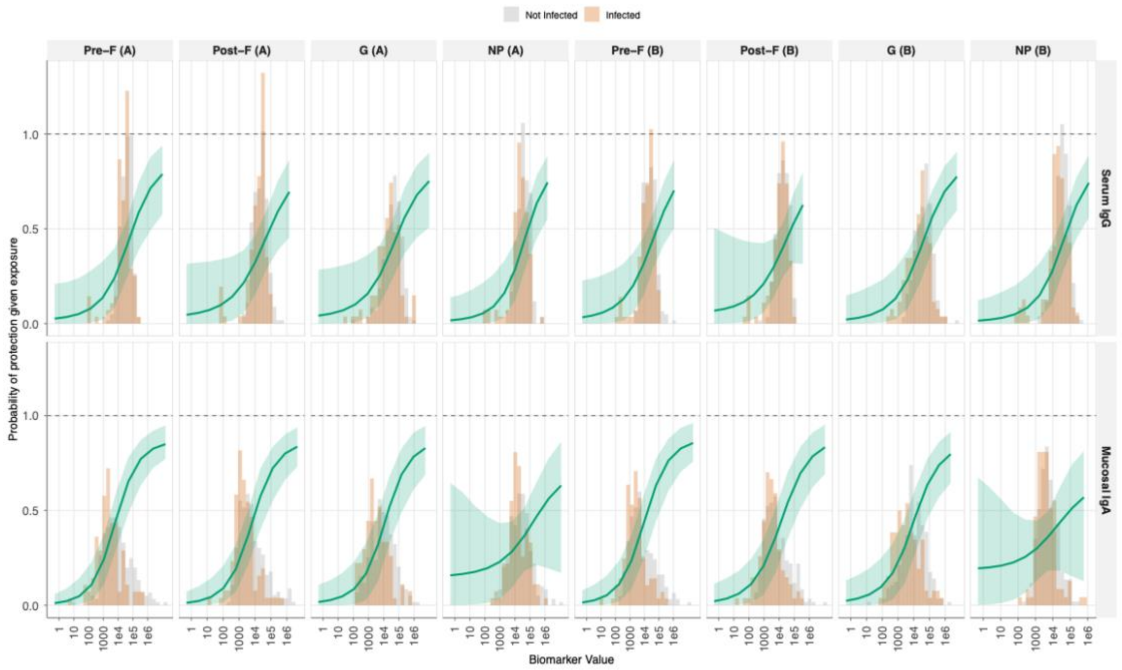


**Supplementary Results Figure 4.9. Correlate of protection (CoP) curves showing the relationship between antibody titre at infection and probability of protection from RSV infection for all 16 biomarker-antigen combinations. . $\boldsymbol{\rho = 0.7}$** Serum IgG, top row; mucosal IgA, bottom row and columns are viral antigen target (PreF, PostF, G, and NP for both RSV-A and RSV-B strains). The solid green line represents the mean estimated probability of protection given exposure to infection as a function of antibody titre, with shaded ribbons indicating 95% credible intervals. Background histograms show the distribution of antibody titres at infection for infected individuals (orange) versus non-infected individuals (gray).

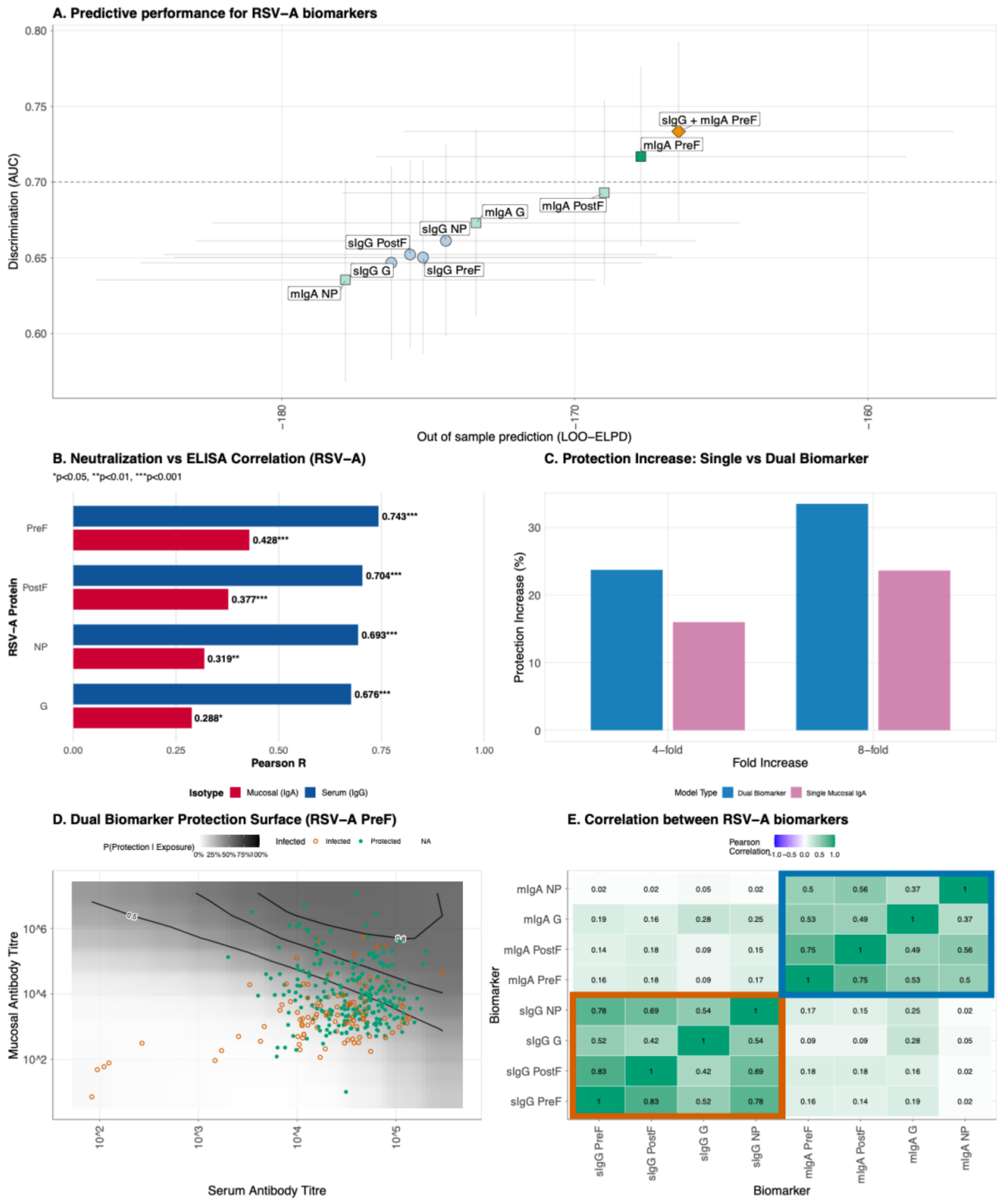


**Supplementary Results Figure 4.10. Comparison of single and dual biomarker models for predicting protection against RSV-A . $\rho = 0.7$ infection.** (A) Model performance comparison across single biomarker models and the dual biomarker model, defined by out-of-sample predictive accuracy (LOO-ELPD, x-axis) and discrimination ability (area under the ROC curve, AUC, y-axis). Circles indicate serum IgG models, squares represent mucosal IgA models, and the triangle denotes the dual biomarker model combining serum IgG and mucosal IgA to RSV-A PreF. The best-performing model within each biomarker class is highlighted with darker shading. Error bars show the standard error of LOO-ELPD (horizontal) and 95% confidence intervals for AUC (vertical). The dashed horizontal line

indicates an AUC of 0.7. (B) Summary of Pearson correlation coefficients for RSV-A protein antigens, comparing serum IgG (blue) and mucosal IgA (red) ELISA titres with neutralisation IC50 values. (C) Counterfactual analysis comparing the increase in protection from boosting pre-infection antibody titres by 4-fold or 8-fold using either a single mucosal IgA biomarker model (purple) or a dual biomarker model combining mucosal IgA and serum IgG (blue). (D) Two-dimensional protection surface for the dual biomarker model using serum IgG and mucosal IgA antibodies to RSV-A PreF. The continuous surface represents the predicted probability of protection given exposure as a function of both serum (x-axis) and mucosal (y-axis) antibody titres. Black contour lines delineate regions of constant protection probability at 50%, 70%, 80%, and 90%. Individual data points overlay the surface, with filled circles representing participants who were protected despite exposure and open circles representing those who became infected. Antibody titres are displayed on a log10 scale. (E) Pearson correlation coefficients between RSV-A variant biomarkers at the time of infection. Color intensity represents correlation strength, ranging from blue (weak correlation) through white (no correlation) to green (strong positive correlation).

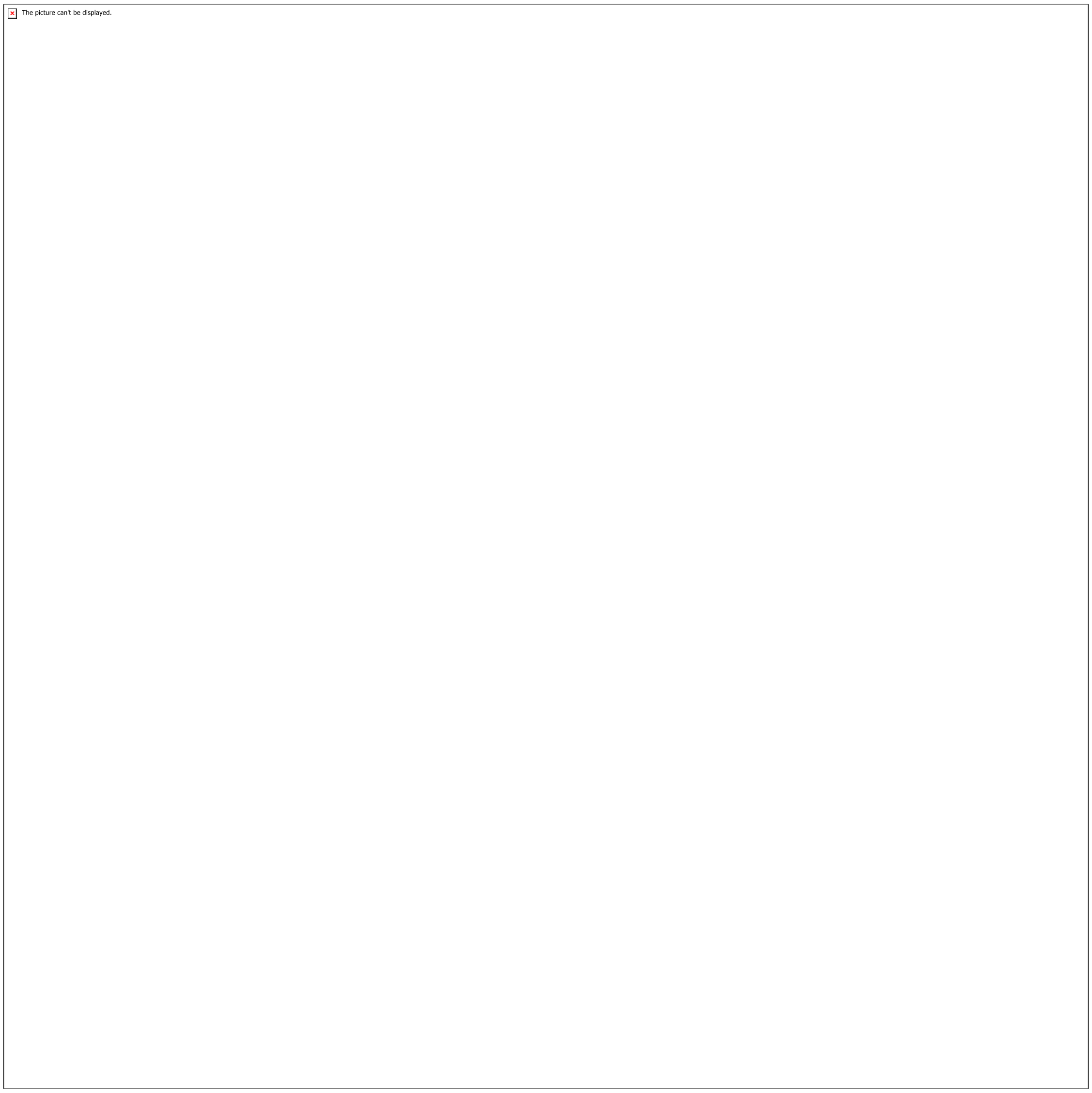


**Supplementary Results Figure 4.11. Age-dependent patterns in antibody magnitude and boosting following natural RSV infection. . $\boldsymbol{\rho = 0.7}$** (A) Baseline and peak antibody titres stratified by age group for serum IgG (left panel) and mucosal IgA (right panel) against four RSV-A antigens (PreF, PostF, G, NP). Gray squares represent baseline titres at the time of infection, colored circles represent peak titres post-infection, and gray lines connecting them indicate the magnitude of antibody boosting. Point are colored according to age groups. (B) Relationship between baseline antibody titre at infection (x-axis) and fold-rise in titre post-infection (y-axis), illustrating the antibody ceiling effect where individuals with higher pre-existing titres exhibit smaller relative boosts. Linear regression lines (solid) with 95% confidence intervals (shaded regions) demonstrate the negative relationship.

The picture can't be displayed.

**Supplementary Results Figure 4.12. Age-stratified analysis of antibody responses and protective immunity for RSV-A PreF biomarkers.** $\boldsymbol{\rho = 0.7}$ **(** A) Antibody response magnitudes by age group, with baseline antibody titres at time of infection (hollow circles) and peak titres following infection (filled circles) for serum IgG and mucosal IgA antibodies to RSV-A PreF protein, stratified by age group (0-5 years in orange, 6-15 years in light blue, 16+ years in green). (B) Model performance by age group. The scatter plot compares out-of-sample predictive accuracy (LOO-ELPD, x-axis) and discrimination ability (AUC, y-axis) for correlate of protection models fitted separately within each age stratum.. Vertical error bars show 95% confidence intervals for AUC; horizontal error bars indicate standard error of LOO-ELPD. (C). Age-specific correlate of protection curves demonstrating the relationship between antibody titre at infection and probability of protection. (D) Protection probability trajectories by age group following infection. The lines show the predicted probability of protection against reinfection over time (0-180 days post-infection) for serum IgG (left panel) and mucosal IgA (right panel) antibodies to RSV-A PreF. Solid lines represent the median posterior prediction for each age group, with shaded ribbons indicating 95% credible intervals
